# The evolving worldwide dynamic state of the COVID-19 outbreak

**DOI:** 10.1101/2020.11.26.20239434

**Authors:** Jose M. G. Vilar, Leonor Saiz

**Affiliations:** Biofisika Institute (CSIC, UPV/EHU), University of the Basque Country (UPV/EHU), P.O. Box 644, 48080 Bilbao, Spain; IKERBASQUE, Basque Foundation for Science, 48011 Bilbao, Spain; Department of Biomedical Engineering, University of California, 451 E. Health Sciences Drive, Davis, CA 95616, USA

## Abstract

The dynamic characterization of the COVID-19 outbreak is critical to implement effective actions for its control and eradication but the information available at a global scale is not sufficiently reliable to be used directly. Here, we integrate multiple data sources through dynamical constraints to quantify its temporal evolution and controllability around the world and within the United States. Overall, the numbers of actively infectious individuals have remained high beyond targeted controllability, with worldwide estimates of 10.24 million on November 24, 2020, totaling in 266.1 million cumulative infections growing at a rate of 11.12 million new infections per week. The actively infectious population reached a local maximum of 7.33 million on July 16, 2020 and remained virtually stagnant at a global scale, with growth rates for most countries around zero that compensated each other, until reverting to net growth on September 22, 2020. We validated the approach, contrasting with prevalence data and the effects of nonpharmaceutical interventions, and we identified general patterns of recession, stabilization, and resurgence. The diversity of dynamic behaviors of the outbreak across countries is paralleled by those of states and territories in the United States, converging to remarkably similar global states in both cases. Our results offer precise insights into the dynamics of the outbreak and an efficient avenue for the estimation of the prevalence rates over time.

## INTRODUCTION

The global spread of the COVID-19 outbreak has had a major global impact. As of November 24, 2020, there have been 1.67 million reported deaths and 72.4 million confirmed cases [1]. Massive travel restrictions, imposed quarantines, lockdowns, and other nonpharmaceutical interventions (NPIs) around the world have been able to slow down the progression of the outbreak, but their gradual lifting has already led to its deadly resurgence in multiple areas [2]. Currently, it is still unclear how best to proceed to balance the economic, societal, ethical, and health trade-offs present. The most widespread quantification, based on the basic reproduction number, is insufficient to address the different trade-offs [3]. It indicates whether the outbreak is growing up or dying out, but it misses crucial information, including the timescales of the processes, the latent potential for a resurgence of the outbreak, and the possibility of actively controlling and tracing the infectious population.

Obtaining a detailed dynamic characterization of the outbreak, however, has proved to be particularly challenging and has been achieved only over small controlled populations. This characterization involved testing for the causative virus SARS-CoV-2, identifying contacts and the infection time, and following up the clinical evolution [4-7]. The available epidemiological information has not been sufficiently reliable to be used directly and the detailed characterization is not feasible at a global scale. Methods based on compartmental models [8] or Bayesian approaches [9] have relied on location-specific information that is not readily available at a global scale.

Here, we uncovered optimal dynamical constraints to integrate the available detailed clinical information with demographic data and epidemiological curves, and we applied them to the quantification of the key determinants of the temporal evolution and controllability of the COVID-19 outbreak, including the number of infectious individuals, the growth rate of the infectious population, and the size of the infected population. We considered explicitly countries in the world and states and territories within the United States (US) with at least 30 COVID-19 reported deaths, which covers 97.4% of the world and 99.9% of the US population.

## RESULTS

### Optimal dynamical constraints

The approach considers the dynamics of the infectious population, *n*_*I*_(*t*), at time *t* described through the expression

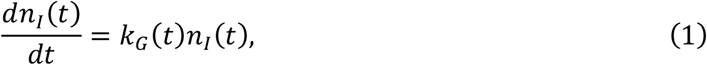

which establishes the definition of the (per capita) growth rate *k*_*G*_(*t*). This expression is completely general and independent of the underlying dynamics of the infection.

The underlying infection dynamics dictates the relationship of *k*_*G*_(*t*) and *n*_*I*_(*t*) with the different epidemiological quantities. Based on an infection-age structured mathematical description [10, 11], we have developed an approach to uncover the optimal dynamic constraints for these relationships in terms of delays and scaling factors (Supplementary Information).

We find that *n*_*I*_(*t*) is optimally related to the rate of increase of the expected cumulative deaths, *n*_*D*_, at a later time *t* + *τ*_*D*_ according to

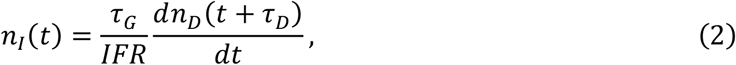

where *IFR* is the infection fatality rate and *τ*_*G*_ is the average generation time. This equation explicitly takes into account that, on average, an infectious individual within *n*_*I*_(*t*) has been infected at time 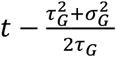 and potentially dies with probability *IFR* at a time *τ*_*I*_ + *τ*_*OD*_ after infection. Here, 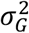 is the variance of the generation time, and *τ*_*I*_ and *τ*_*OD*_ are the incubation and symptom onset-to-death average times, respectively. The values of these characteristic times have been estimated in days as *τ*_*G*_ = 6.5, *σ*_*G*_ = 4.2, *τ*_*I*_ = 6.4, and *τ*_*OD*_ = 17.8 from precise follow up of specific groups of patients [5, 12, 13], which leads to 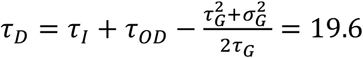 (Supplementary Information).

A general assumption of the approach is that the *IFR* for each age group remains the same for all locations [5] and that the overall *IFR* for each location is obtained as the average over its specific population’s age distribution.

The growth rate is obtained directly from Eqs. (1) and (2) as

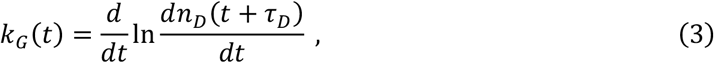

which is related to the time-varying reproduction number, *R*_*t*_, through the Euler–Lotka equation (Supplementary Information).

The expected cumulative number of infected individuals at a time *t, n*_*T*_(*t*), follows from

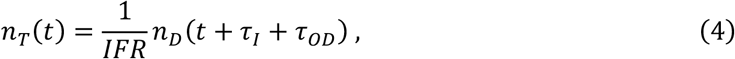

which is obtained also from the dynamical constraints (Supplementary Information).

To compare with prevalence studies, we used the dynamical constraints (Supplementary Information) to obtain the relationship of the expected number of seropositive individuals, *n*_*SP*_(*t*), with the infected population,

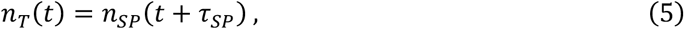

where *τ*_*SP*_ is the average seroconversion time after infection. We also obtained the relationship of the expected number of positive reverse transcription polymerase chain reaction (RT-PCR) testing individuals, *n*_*TP*_(*t*), with the infectious population,

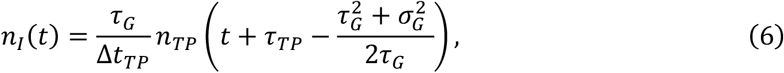

where *τ*_*TP*_ is the average time for testing positive after infection and Δ*t*_*TP*_ is the average number of days of positive testing. The values of these additional characteristic times have been estimated in days as *τ*_*SP*_ = 13.4, *τ*_*TP*_ = 14.4, and Δ*t*_*TP*_ = 20 from clinical studies [13-16] (Supplementary information).

### Implementation

Eqs. (1) – (4) completely characterize the dynamics of the outbreak from the expected cumulative deaths, the age structure of the population, and the general clinical parameters of the infection. We used a workflow (Fig. 1A) that incorporates explicitly the death counts compiled by the Johns Hopkins University Center for Systems Science and Engineering [1], the age structure reported by the United Nations for countries [17] and the US Census for states and territories [18], previously estimated aged-stratified *IFR* [5], and other previously estimated clinical parameters [5, 12-16]. The expected number of daily deaths was inferred using density estimation from the death curves after preprocessing to minimize reporting artifacts (Supplementary Information). Eqs. (5) and (6) were used to set the appropriate scale and delay to compare with seroprevalence studies.

**Fig 1.**
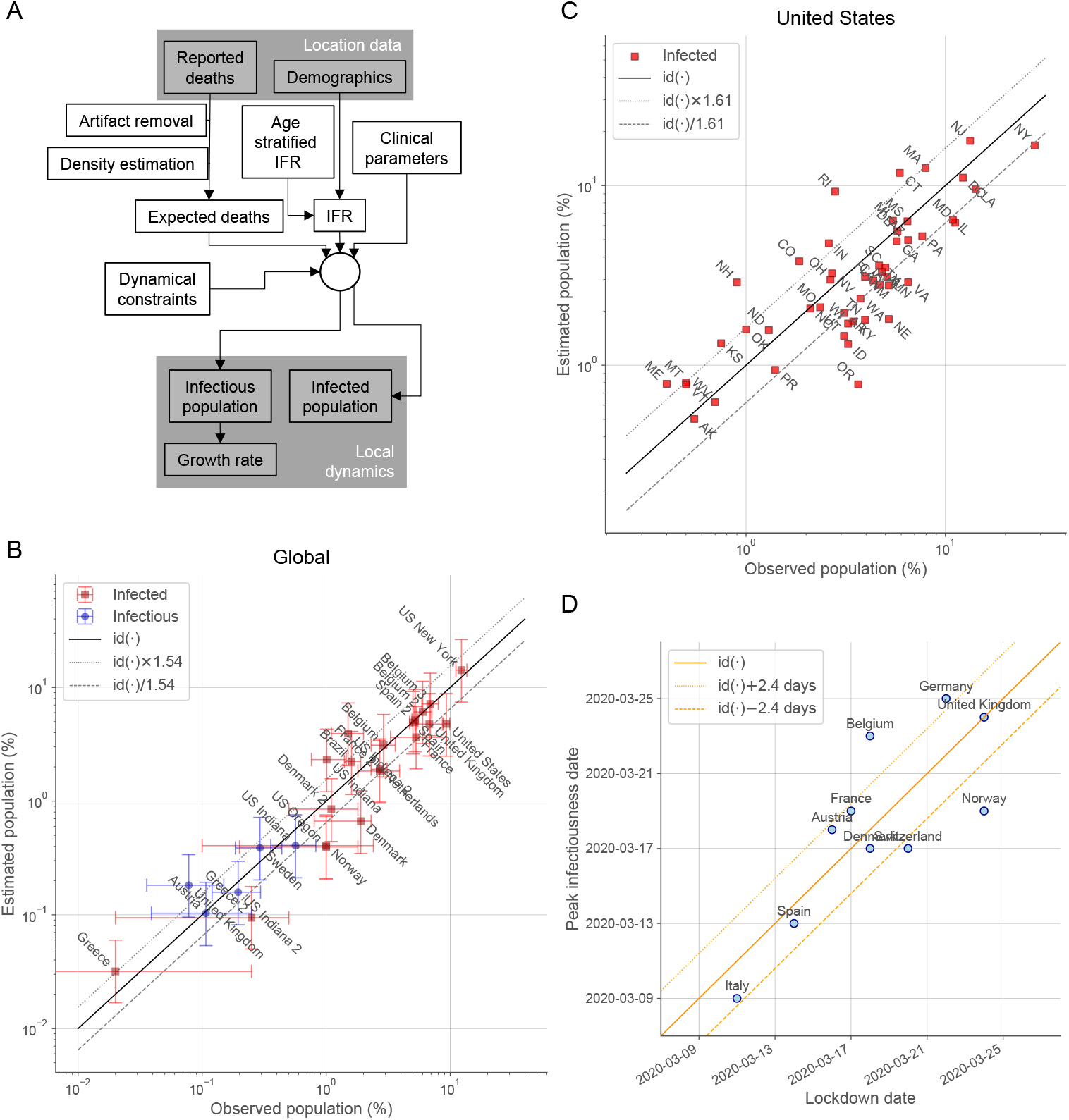
The approach consistently estimates prevalence and the timing of NPIs. The approach (**A**) has been validated with prevalence data of the infectious (PCR-RT testing) and infected (antibody testing) populations at a global scale (**B**) and for states within the US (**C**). The continuous black lines (**B, C**) represent the perfect prediction (identity function denoted by id(·)) with the parallel dotted/dashed lines indicating the fold accuracy. Global data were obtained from sources described in Supplementary Table S1. Several locations have estimates for multiple date ranges. State data correspond to two studies with specimen samples taken primarily on the first two weeks of July [19] and of August, 2020 [20]. The inferred timings of the peak infectiousness are plotted against the dates of the major country-wide lockdowns in Europe [9] (**D**). The continuous orange line represents perfect concordance, and the parallel dotted/dashed lines indicate the mean absolute error.

### Validation against prevalence data and NPIs

To validate the approach, we contrasted the estimated infectious and infected populations with the results from available antibody seroprevalence and RT-PCR testing studies for countries in the world and locations in the US with at least 30 reported deaths (Fig. 1B). The observed and estimated values agree with each other within the 95% credibility intervals (CrI), with a 1.54-fold accuracy over almost a 1,000-fold variation and a correlation coefficient (*ρ*) on a logarithmic scale of *ρ* = 0.94.

We combined into a separate analysis (Fig. 1C) the data of two additional comprehensive antibody seroprevalence studies within the US, which included 49 [19] and 38 [20] states with non-zero prevalence values. For the states present in the two studies, we considered their average values. The estimated values agree with the observed prevalence with 1.62-fold accuracy and *ρ* = 0.81. The agreement for the combined data is better than for the data of each of the studies independently (1.65-fold accuracy and *ρ* = 0.74 for one study [19] and 1.74-fold accuracy and *ρ* = 0.77 for the other study [20]) and better than the agreement of both studies with each other (*ρ* = 0.55 for the 38 states common to both studies), indicating that the estimations of the approach fall within the observed variability of the prevalence studies. Indeed, the approach is effectively unbiased collectively, with the (geometric) average of the estimations being just a factor 1.12 (globally) and 1.14 (US) larger than that of the observations.

Overall, the validation of the approach shows that it can reproduce the available prevalence studies over a factor 615.0 between minimum and maximum values for countries and states with 72.0% (globally) and 79.6% (US) of the estimates within a factor 2 of the observed values, and with 100.0% (globally) and 93.9% (US) within a factor 3. There are no countries and only three states with values outside the factor 3 boundary. In the cases of Rhode Island and New Hampshire, the estimated infectious population is larger than the ones reported by one study [20], which is consistent with the observed age-specific seroprevalence biased to older populations. In the case of Oregon, the estimated infectious population is smaller than the ones reported [19, 20], which is consistent with the observed age-specific seroprevalence biased to younger populations [20]. In the case of Oregon, in addition, only 338 of the 1123 excess deaths during the outbreak before the collection of the specimens for analysis were attributed to COVID-19 [21].

We also validated the ability of the approach to capture the effects of NPIs (Fig. 1D). The inferred timings of the peak infectiousness (maximum infectious population) are concordant with the dates of the major early country-wide lockdowns in Europe [9], with an overall average deviation of 0.0 days between lockdown and peak dates and a mean absolute deviation of 2.4 days. This concordance also indicates that there is only a small variability in the average timing between infection and death among those countries.

### Outbreak progression motifs

We analyzed explicitly the time evolution of the growth rate, the infectious population, the cumulative number of infections, and the relationship between growth rate and infectious population for all countries and states with at least 30 deaths (Supplementary Fig. S1 and Table 1).

**Table 1.** Key estimates of the outbreak.

|  | **World** | **United States** |
| --- | --- | --- |
| **Highest local maximum of the infectious population** | 7.33 million on July 16, 2020 | 1.41 million on March 28, 2020 |
| **Current infectious population** | 10.24 million on November 24, 2020 | 1.37 million on November 24, 2020 |
| **Infected population** | 266.1 million as of November 24, 2020 | 31.8 million as of November 24, 2020 |
| **Underreporting factor** | 3.7 (only 72.4 million cases reported) | 2.5 (only 12.6 million cases reported) |
| **New infections per week** | 11.12 million | 1.50 million |
| **Growth rate: 10% quantile** | -0.014/days<br>(48.5-day half-life) | 0.005/days<br>(138.2-day doubling time) |
| **Growth rate: median** | 0.002/days | 0.017/days |
| **Growth rate: 90% quantile** | 0.022/days<br>(30.8-day doubling time) | 0.024/days<br>(28.7-day doubling time) |
| **Population considered (locations with at least 30 deaths)** | 97.4% | 99.9% |
| **Number of locations considered** | 153 | 53 |
| **Number of locations with more than 10% of the population infected** | 20 | 18 |
| **Number of locations with more than 20% of the population infected** | 1 | 1 |
| **Number of locations with less than 7 actively infectious individuals per 100,000 population** | 31 | 0 |
| **Number of locations that reached 1,000 actively infectious individuals per 100,000 population** | 11 | 11 |
| **Correlation coefficient between infectious/infected population estimates and observations** | 0.94 | 0.81 |
| **Infectious/infected population estimates within a factor 2 of the observations** | 72.0% | 79.6% |
| **Infectious/infected population estimates within a factor 3 of the observations** | 100.0% | 93.9% |

The characterization of the dynamics in terms of the growth rate and infectious population (Supplementary Fig. S1) shows that there are prototypical types of behavior, or motifs (highlighted with representative examples in Fig. 2). Locations either contained (Figs. 2A-2C) or amplified (Figs. 2E-2F) the outbreak in its initial stages. In the case of initial containment, the behavior branched subsequently into contained sporadic resurgences (Fig. 2A), uncontrolled resurgence of the infectious population over the initial maximum infectiousness (Fig. 2B), and sustained decrease of the outbreak (Fig. 2C). The specific behavior depended on the success of the measures implemented, e.g. targeted control and moderate lockdowns, and their subsequent relaxation [22]. In the case of initial amplification, the dynamics proceeded in diverse ways, including an increasing infectious population converging to zero growth (Fig. 2D), a fast-evolving infectious population switching from positive to negative growth (Fig. 2E), and fast convergence to subsequent sustained residual growth (Fig. 2F). In general, the locations that reached a substantial negative growth rate are those that implemented long-term strict lockdowns, whereas zero or small positive growth rates correspond to intermediate measures with partial restrictions [22]. In many cases, lifting restrictions has led to fast switching from negative to positive sustained growth, as illustrated by United Kingdom, and Italy (Fig. 2E), indicating the inability of these locations to contain the outbreak even at low values of the infectious population.

**Fig 2.**
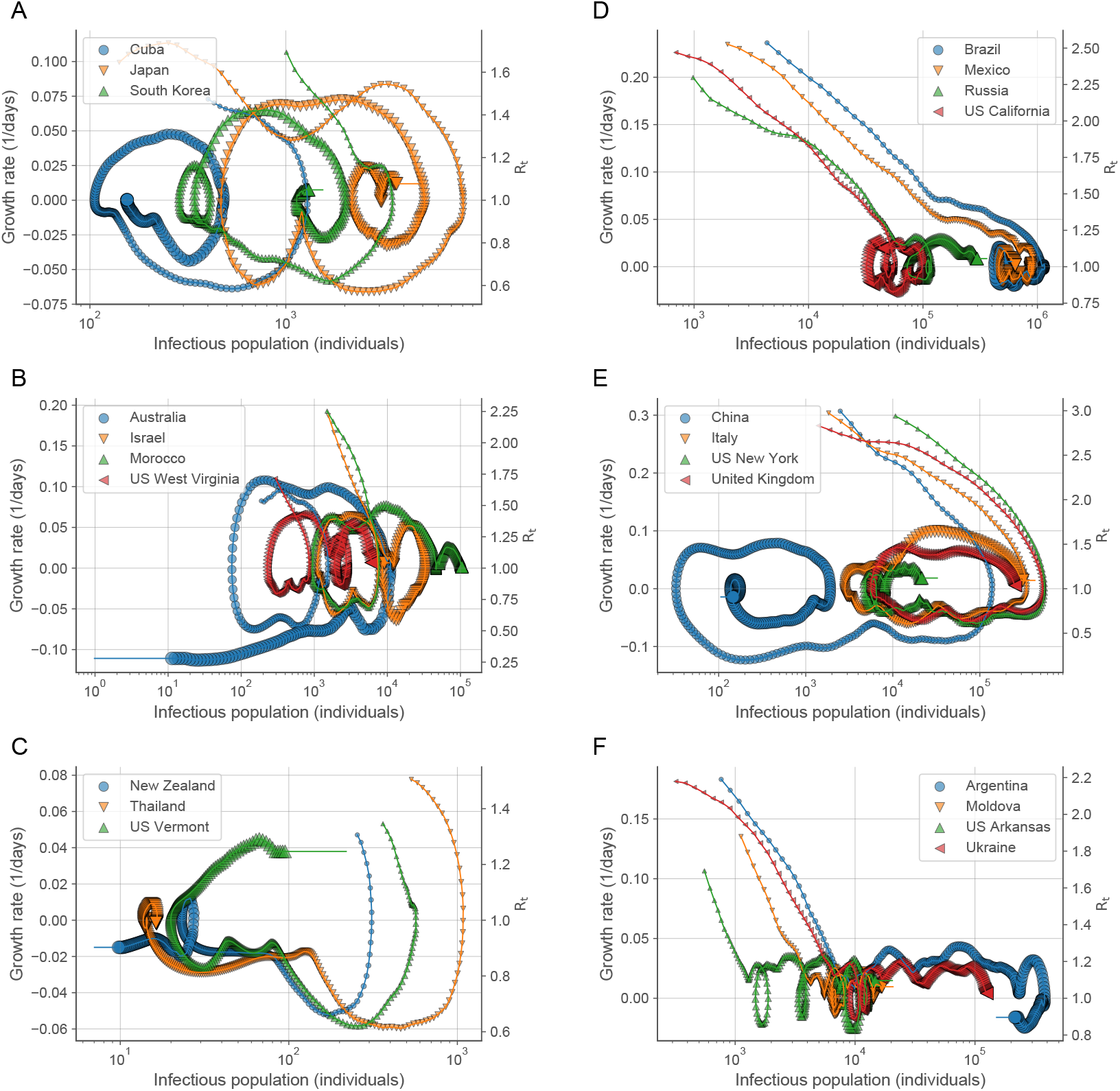
Trajectories in the growth rate-infectious population space follow prototypical types of behavior. Each day is indicated by a symbol increasing in size with time. The largest symbol corresponds to the last day of the estimation (November 2, 2020). The colored lines at the end indicate the extrapolation to current time (November 24, 2020). The right axes indicate the growth rate in the scale of the reproduction number as *R*_*t*_ = 1 + *k*_*G*_(*t*)*τ*_*G*_ (Supplementary Information). Common types of behavior include initial containment (**A, B, C**) with subsequent minor increases (**A**), amplified resurgence (**B**), and a sustained regression (**C**) of the outbreak; and initial amplification (**D, E, F**) with slow convergence to zero growth (**D**), convergence to negative growth (**E**), and fast convergence to initial sustained residual positive growth (**F**). The prefix “US “has been added to the name of the locations in the US. Confidence intervals for the growth rates and infectious population are provided in Supplementary Fig. S1.

### Global dynamics

At a global scale, the outbreak is characterized by two early local maxima of the overall worldwide infectious population on January 25, 2020 and March 25, 2020, coincidental with the regression of the outbreak in China, initially, and in Europe, afterward, reaching the highest local maximum of 7.33 million active infections (Figs. 3A and 3B) on July 16, 2020 with a subsequent increasing trend since September 22, 2020. In the US, there has been an overall maximum of the infectious population on March 28, 2020 with 1.41 million active infections and a local maximum on July 15, 2020 (Figs. 3D and 3E). The estimated infected population is 266.1 million, growing at a rate of 11.12 million new infections per week, for the World (Fig. 3C) and 31.8 million, growing at a rate of 1.50 million new infections per week, for the US (Fig. 3F), which is a factor 3.7 for countries in the World and 2.5 for locations in the US higher than the corresponding reported cases [1].

**Fig 3.**
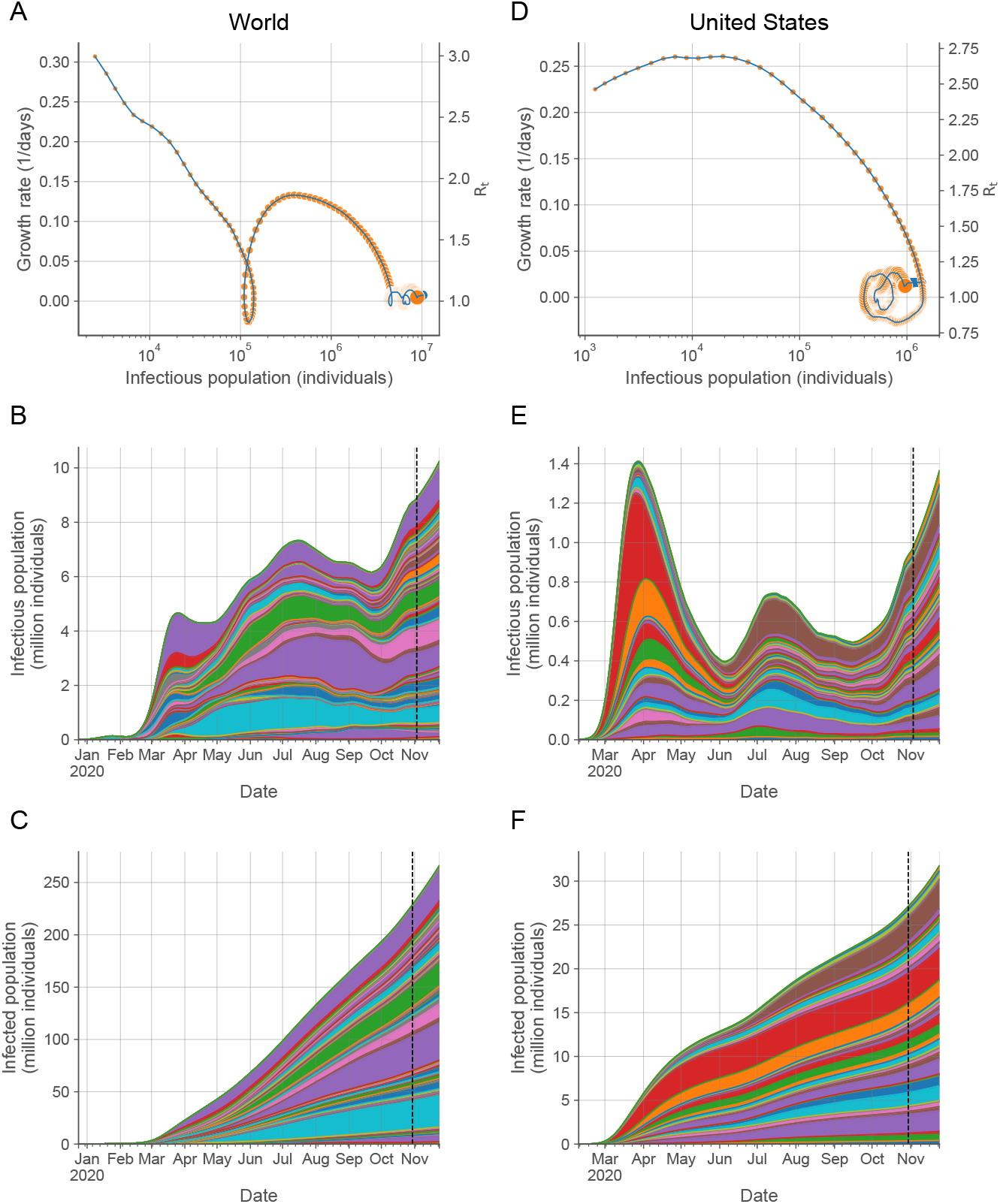
Characterization of the temporal evolution of the COVID-19 outbreak. Trajectory in the growth rate-infectious population space (**A, D**) and infectious (**B, E**) and infected (**C, F**) populations over time of the aggregate values for countries in the world (**A, B, C**) and locations in the US (**D, E, F**). In the trajectories (**A, D**), each day is indicated by a symbol increasing in size with time, ending with the largest symbol on November 2, 2020. The blue line at the end indicates the extrapolation to current time (November 24, 2020). The right axes indicate the growth rate in the scale of the reproduction number as *R*_*t*_ = 1 + *k*_*G*_(*t*)*τ*_*G*_ (Supplementary Information). Each colored region in the area plots represents the contribution of a country (**B, C**) or a state (**E, F**) to the overall infectious and infected populations. Countries and states are arranged in alphabetical order from bottom to top. The vertical dashed lines (**B, C, E, F**) indicate the transition from inference to extrapolation. Individual data for all the countries and states are provided in Supplementary Fig. S1.

### State of the outbreak across locations and its controllability

At a local scale, the per capita infectious population of countries and states has only exceptionally crossed the 1% value (11 out of 153 countries and 11 out of 53 states) (Fig. 4). The infected populations have surpassed the 10% of the total population only for 20 countries and 18 states (Fig. 4), with just one country and one state reaching the 20% value. These results confirm that relying on herd immunity is not a realistic option. Controlling the outbreak by actively tracing and isolating newly and potentially infected individuals has been successfully implemented in South Korea, with the use of large resources and with occasional outbreaks that required short-term extended human-interaction restrictions, and almost successfully in Japan, with voluntary business closures and other restrictions [22]. These countries have always remained below 7 actively infectious cases per 100,000 individuals (0.007% of their population), with average values of 2.2 (South Korea) and 2.5 (Japan) since March 1, 2020. None of the states and 31 of the countries analyzed have had a per capita number of infectious individuals below the average value of South Korea since May 1, 2020, which indicates that 61.1% of the World (79.5% excluding China) and 99.9% of the US human population reside in countries or states that have not allowed targeted controllability of the outbreak with the resources used by South Korea. This inability to control the outbreak as soon as restrictions are lifted, even at low values of the infectious population but above the South Korea average value, is illustrated by United Kingdom and Italy (Fig. 2E).

**Figure 4.**
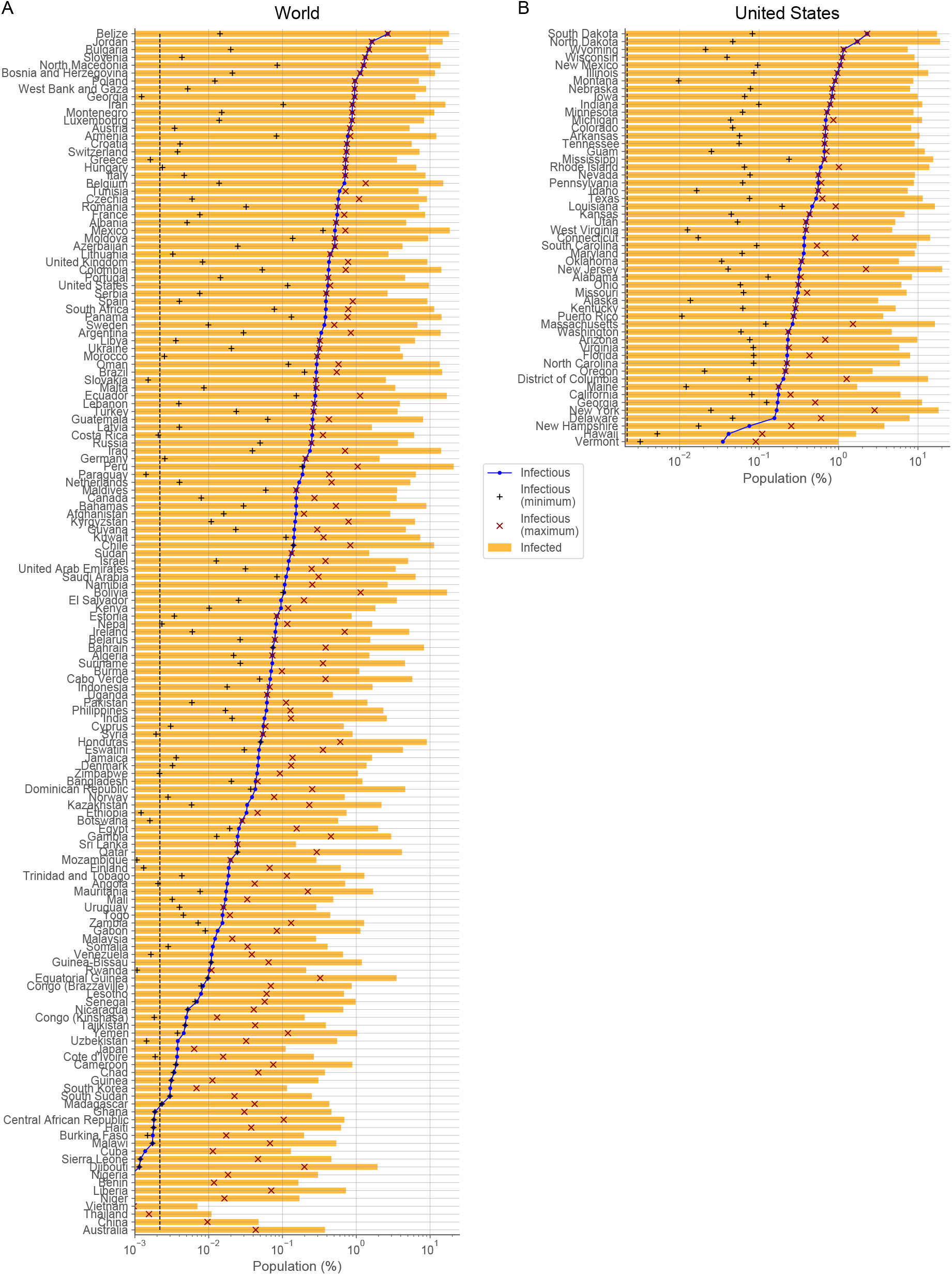
Per capita state of the outbreak. The percentage of infectious and infected populations on the last day estimated (November 24, 2020) and of the minimum and maximum infectious population reached are shown for countries in the world (**A**) and locations in the US (**B**). The minimum was computed for values after May 1, 2020. Countries and US locations have been sorted according to their per capita infectious population. The vertical dashed lines (**A, B**) indicate the time-averaged infectious population of South Korea since March 1, 2020.

### From local to global dynamics

The collective properties of the individual local dynamics, as quantified by the distribution of growth rates across countries and states over time (Fig. 5), shows a progressive double stabilization of the outbreak before a subsequent growth period starting in late September 2020. At a local scale, growth rates for most countries, whether positive or negative, decreased in absolute value, leading to a slowdown of the dynamics. At a global scale, positive and negative values have converged towards statistically compensating each other, decreasing further the overall net growth of the infectious population. Specifically, the latest estimates of the growth rates on November 2, 2020 for countries in the world have a median value of 0.002/days, with 80% of the countries within the narrow range of values from −0.014/days to 0.022/days, which implies a slow local dynamics with half-lives and doubling times of 48.5 and 30.8 days, respectively. For locations in the US, the median value of the growth rates is 0.017/days, with 80% of locations ranging from 0.005/days (138.2 days doubling time) to 0.024/days (28.7 days doubling time).

**Fig 5.**
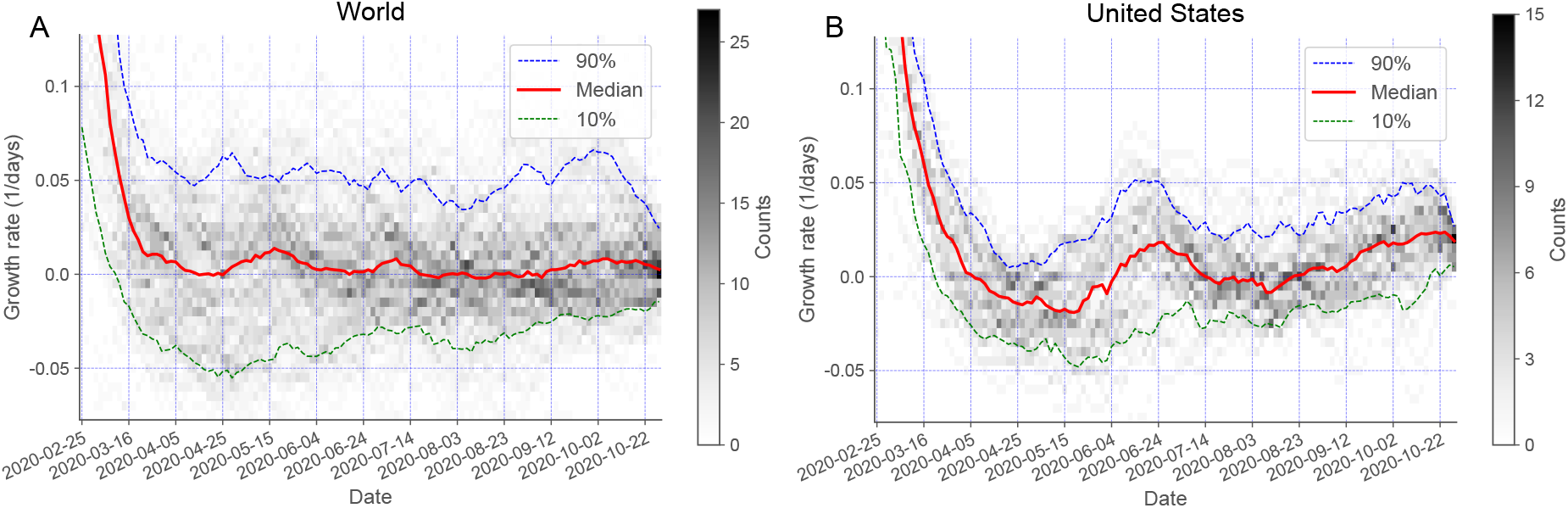
The temporal evolution of the distribution of growth rates shows a double stabilization of the outbreak. The temporal evolution of the median (red line), first decile (green line), and tenth decile (blue line) of the growth rates are plotted over the gray-coded histogram for countries in the world (**A**) and states and territories of the US (**B**). The median converges towards zero values and the deciles move closer to each other in a fluctuating manner for countries in the World and locations in the US.

## DISCUSSION

The dynamical constraints we have obtained through a detailed infection-age mathematical description of the outbreak allowed us to find the optimal time delays and scaling factors to connect the evolution of the reported death counts over time with those of the infectious and infected populations. Overall, integrating these constraints through a workflow with essential preprocessing steps showed that the approach can consistently infer the precise timing of NPIs and estimate prevalence data across countries in the world and locations in the US. A prominent feature of the approach is its ability to provide reliable results even for low death counts, which overcomes the major limitations of choosing between unreliable infection case data (highly dependent on testing rates) or noisy death counts as input to the inference problem [9].

The approach assumes a general age-stratified *IFR*. In general, these quantities are expected to depend potentially on specific features of the population and the medical care facilities available. The available studies show a minimal variability among different countries and other locations that reported on prevalence [23]. It also assumes an age-uniform exposure (attack rate), which is consistent with data for other respiratory diseases [5] and holds to a large extent when there is information available for COVID-19 [19, 24, 25]. We have also assumed a constant generation interval typical of non-confinement locations, which has been observed to shorten in some cases by NPIs [26]. Prevalence studies can also depend on the diminishing antibody levels after infection [14, 27], collecting and processing specimens for analysis [28], and potential biases towards specific population groups [19]. In addition, there might be a degree of under-reporting of COVID-19 deaths, as suggested by excess mortality not attributable to other causes than COVID-19 [21]. Our results show that all these potential deviations on the assumptions, on the data, and on prevalence studies collectively have only a restricted impact on the approach, with 72.0% (globally) and 79.6% (US) of the estimates within a factor 2 of the observed values and 100.0% (globally) and 93.9% (US), within a factor 3. This accuracy of the estimations is highly remarkable in the context of the observed prevalence spread over a factor 615.0 between minimum and maximum values, ranging from 0.02% to 12.30%.

The analysis in terms of the growth rate–infectious population trajectories has revealed universal types of behavior of the outbreak for countries around the world and locations within the US. This information can be used to anticipate the response to enacting, modifying, or lifting NPIs. The most marked example is the response to strict lockdowns across countries in Europe (e.g. United Kingdom, Italy, Belgium, Spain, France, Germany, Austria, Netherlands, and Switzerland) and Northeast states in the US (e.g. New York, New Jersey, Massachusetts, Pennsylvania, and District of Columbia). They followed the same type of behavior (fast decrease of the growth rate from high to sustained negative values) until major restrictions were lifted in the European countries [22], turning the growth rate of their infectious populations into highly sustained positive values. Our results show that those European countries had small actively infectious populations but not as small as required for targeted controllability. They also show that most Northeast states in the US are in a similar resurgence path but at much earlier stages, with many of their NPIs still in place and with markedly smaller growth rates, which makes their reaching as deadly a resurgence as in Europe still avoidable.

At a global scale, the outbreak has reached a net growth rate fluctuating near zero values but with a high infectious population. A similar state has also been present in the US. This type of fluctuating states, with long stagnant overall infectious population periods and median growth rate close to zero, is expected of bounded unsynchronized fluctuating populations [29], such as those from uncoordinated locations aiming at just preventing an unrestricted expansion of the outbreak rather than at its eradication. This widespread feature is present for both countries in the world and locations within the US. Considering the NPIs implemented [22], our results show that there have been locations with interventions to move the growth rate towards zero values and that there have been locations switching on and off severe measures to decrease temporarily the active infectious population. Despite not growing substantially since reaching its highest local maximum of 7.33 million active infections on July 16, 2020, the high value of the global infectious population attained is currently leading to 11.12 million new infections per week that replace the same ballpark number of individuals that stop being infectious. This high turnover makes the control of any potential resurgence extremely costly.

At a local scale, our results show a highly variable temporal evolution of the infectious populations, both over time for each location and across locations. Having an up-to-date estimate of the infectiousness of populations would allow policymakers to better implement travel planning among locations. The approach has proven to accurately track the effects of local NPIs. We also expect it to play a fundamental role in evaluating the progress of vaccination efforts, especially considering the challenges present, such as waning immunity levels and pathogen evolution [30].

## Data Availability

Details for accessing the data sources are provided in the manuscript.

https://github.com/Covid19Dynamics/trajectories

## ACKNOWLEDGMENTS

J.M.G.V. acknowledges support from Ministerio de Ciencia e Innovación under grant PGC2018-101282-B-I00 (MCI/AEI/FEDER, UE). L.S. acknowledges support from the University of California, Davis.

## SUPPLEMENTARY INFORMATION

Methods; Supplementary Table S1; Supplementary Fig. S1.

## DATA AVAILABILITY

Datasets of the trajectories of the dynamic state of the COVID-19 outbreak for all the countries in the world and states and territories in the United States analyzed are available at https://github.com/Covid19Dynamics/trajectories.

## SUPPLEMENTARY INFORMATION

### MATHEMATICAL DESCRIPTION

#### Infection-age structured dynamics

We consider the infection-age structured dynamics of the number of individuals *u*_*I*_(*t, τ*) at time *t* who were infected at time *t* − *τ* given by

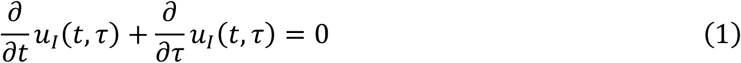

with boundary condition

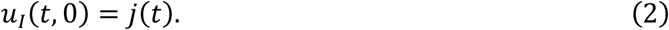

Here, *τ* is the time elapsed after infection, referred to as infection age, and 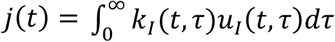 is the incidence, with *k*_*I*_(*t, τ*) being the rate of secondary transmissions per single primary case [10, 11].

The solution is obtained through the method of characteristics as

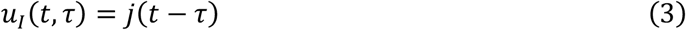

for *t* ≥ *τ* and *u*_*I*_(*t, τ*) = 0 for *t* < *τ*. The resulting renewal equation, 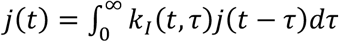, is used as the basis for the definitions of the reproduction number 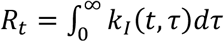 and the probability density of the generation time 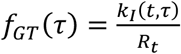.

The infectious population is given by

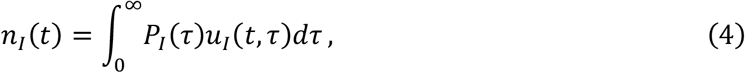

which considers that an individual remains potentially infectious after a time *τ* from infection with probability

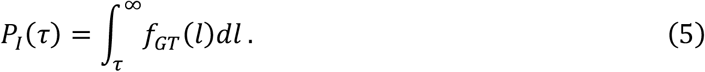

Therefore, in terms of the incidence, we have

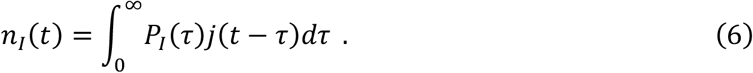

Additionally, we consider the expected cumulative number of infections, *n*_*T*_(*t*), expressed in terms of the overall accumulated incidence as

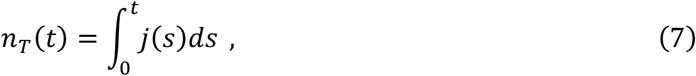

and the dynamics of the expected cumulative deaths, *n*_*D*_(*t*),

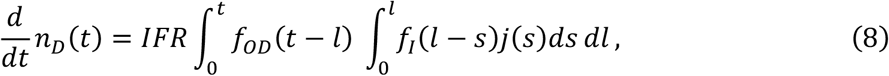

which takes into account that deaths occur with probability given by the infection fatality rate, *IFR*, at times after infection given by the convolution of the probability density functions of the incubation, *f*_*I*_, and symptom onset-to-death, *f*_*OD*_, times.

Similarly, the variation of the expected number of seropositive individuals at a time *t, n*_*SP*_(*t*), is expressed as

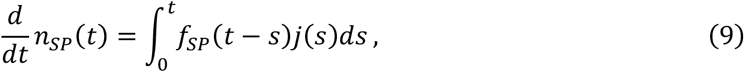

where *f*_*SP*_ is the probability density function of the seroconversion time after infection, and the expected number of individuals with positive RT-PCR testing *n*_*PT*_(*t*), as

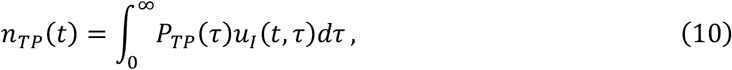

where *P*_*TP*_(*τ*) is the probability that an infected individual would test positive at a time *τ* after infection.

#### Dynamical constraints

To obtain a closed set of equations from the expressions involving an integral 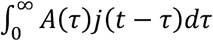 of a function *A* with the incidence *j*, we perform a series expansion of the incidence around the infection-age time *τ*_*A*_,

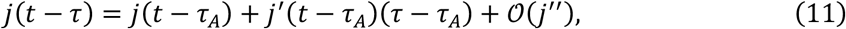

with the value of *τ*_*A*_ chosen as

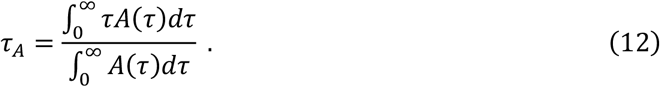

The specific value of *τ*_*A*_ leads directly to a first-order approximation,

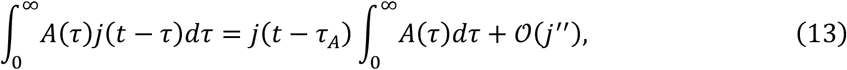

because 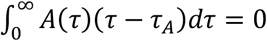.

Using this approach, we obtain

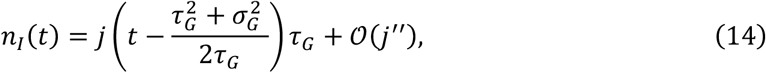

where *τ*_*G*_ and 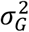 are the average and variance of the generation time, respectively, and

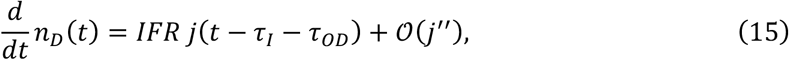

where *τ*_*I*_ and *τ*_*OD*_ are the incubation and symptom onset-to-death average times, respectively, which leads to

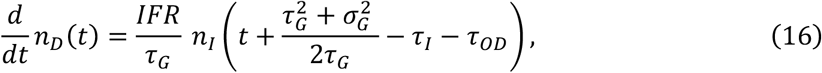

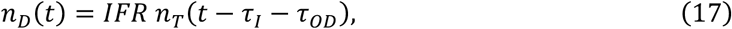

up to *𝒪*(*j*″). These expressions are used to estimate the infectious population *n*_*I*_(*t*) from the daily deaths, 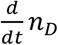, at time 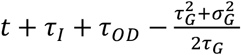 and the cumulative infected population *n*_*T*_(*t*) from the cumulative deaths, *n*_*D*_, at time *t* + *τ*_*I*_ + *τ*_*OD*_, leading to

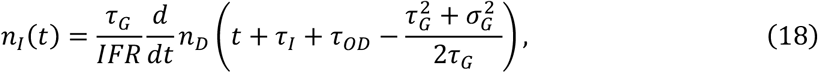

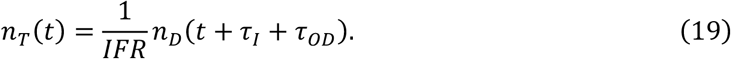

Similarly, we obtain

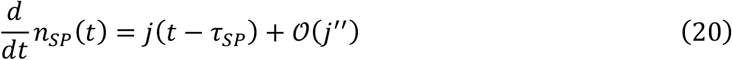

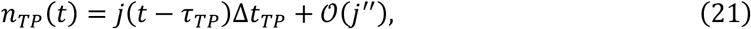

where *τ*_*SP*_ is the average seroconversion time after infection and Δ*t*_*TP*_ is the average number of days an individual tests positive, which up to *𝒪*(*j*″) leads to

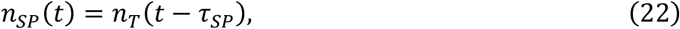

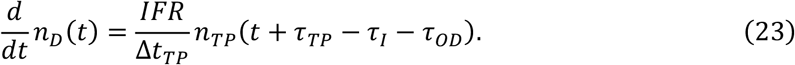

Combining Eqs. (22) and (23) with Eqs. (18) and (19) leads to

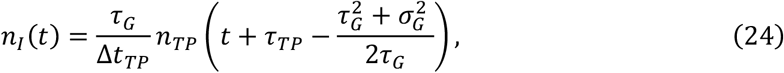

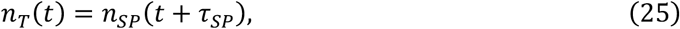

which is used to validate the values of the estimated infectious population *n*_*I*_(*t*) from RT-PCR testing results, *n*_*TP*_, at time 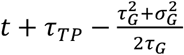 and the cumulative infected population *n*_*T*_(*t*) from seropositivity testing, *n*_*SP*_, at time *t* + *τ*_*SP*_.

### APPROACH WORKFLOW

#### Expected deaths

The raw cumulative death counts over time, *n*_*W*_(*t*), are obtained from the Johns Hopkins University Center for Systems Science and Engineering [1] for countries and for US locations.

The daily death counts Δ*n*_*W*_(*t*) = *n*_*W*_(*t*) – *n*_*W*_(*t* − 1) are considered to contain reporting artifacts if they are negative or if they are unrealistically large. This last condition is defined explicitly as larger than 4 times its previous 14-day average value plus 10 deaths, 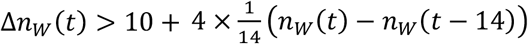, from a non-sparse reporting schedule with at least 2 consecutive non-zero values before and after the time *t*, 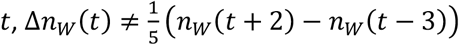.

Reporting artifacts identified at time *t* are considered to be the result of previous miscounting. The excess or lack of deaths are imputed proportionally to previous death counts. Explicitly, death counts are updated as

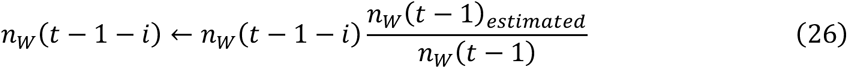

with 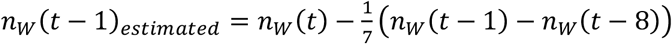 for all *i* ≥ 0. In this way, Δ*n*_*W*_(*t*) is assigned its previous seven-day average value.

The expected daily deaths, Δ*n*_*D*_(*t*), are obtained through a density estimation multiscale functional, *f*_*de*_, as Δ*n*_*D*_(*t*) = *f*_*de*_(Δ*n*_*W*_(*t*)), which leads to the estimation of the expected cumulative deaths at time *t* as 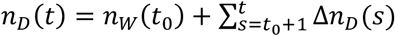. Specifically,

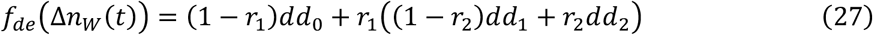

With

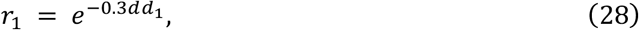

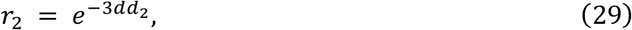

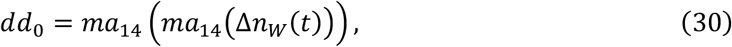

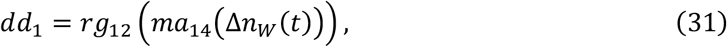

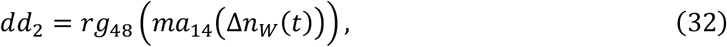

where *ma*_14_(·) is a centered moving average with window size of 14 days and *rg*_*σ*_ (·) is a centered rolling average through a Gaussian window with standard deviation *σ*.

#### Reporting delays

We consider an average delay of two days between reporting a death and its occurrence. This value is obtained by comparing the daily death counts reported for Spain [1] and their actual values [31] from February 15 to March 31, 2020. The values of the root-mean-squared deviation between reported and actual deaths shifted by 0, 1, 2, 3, and 4 days are 77.9, 58.4, 38.5, 58.7, and 88.6 deaths respectively.

#### Infection fatality rate (IFR)

The infection fatality rate is computed assuming homogeneous attack rate as

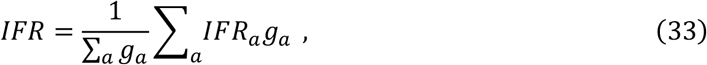

where *IFR*_*a*_ is the previously estimated *IFR* for the age group *a* [5] and *g*_*a*_ is the population in the age group *a* as reported by the United Nations for countries [17] and the US Census for states [18].

#### Clinical parameters

We obtained the values of the average *τ*_*G*_ and standard deviation *σ*_*G*_ of the generation time from Ref. [12], of the averages of the incubation *τ*_*I*_ and symptom onset-to-death *τ*_*OD*_ times from Refs. [5, 13], and of the average number of days Δ*t*_*TP*_ of positive testing by an infected individual from Refs. [14, 16]. The average time at which an individual tested positive after infection *τ*_*TP*_ was computed as *τ*_*TP*_ = *τ*_*I*_ − 2 + Δ*t*_*TP*_/2, where we have assumed that on average an individual started to test positive 2 days before symptom onset. The average seroconversion time after infection *τ*_*SP*_ was estimated as *τ*_*I*_ plus the 7 days of 50% seroconversion after symptom onset reported in Ref. [15].

#### Dynamical constraints implementation with discrete time

We implemented the dynamical constraints to compute the infectious and infected population as outlined in the main text and as detailed in the previous section of this document, using days as time units. Time delays were rounded to days to assign daily values.

The first derivative of the cumulative number of deaths is computed as

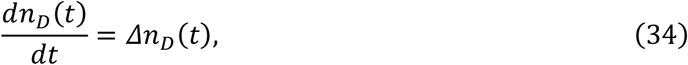

with Δ*n*_*D*_(*t*) = *n*_*D*_(*t*) − *n*_*D*_(*t* − 1).

The growth rate was computed explicitly from the discrete time series as the centered 7-day difference

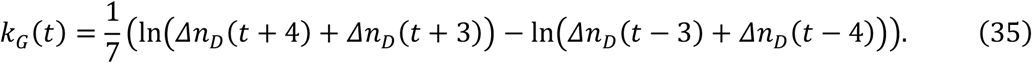

The 7-day value was chosen to mitigate reporting artifacts.

#### Confidence and credibility intervals

Confidence intervals associated with death counts were computed using bootstrapping with 10,000 realizations [32]. These confidence intervals were combined with the credibility intervals of the *IFR* in infectious and infected populations assuming independence and additivity on a logarithmic scale.

#### Fold accuracy

The fold accuracy, *F*_*A*_, is explicitly computed as

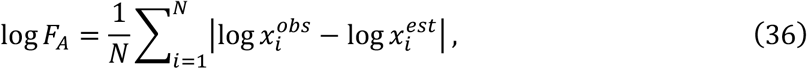

where |·| is the absolute value function, 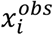 is the *i*^*th*^ observation, 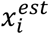 is its corresponding estimation, and *N* is the total number of observations.

#### Inference and extrapolation

Because of the delay between infections and deaths, inference for the values of the growth rate and infectious populations ends on November 2, 2020 and for the values of the infected populations ends on October 29, 2020. Extrapolation to the current time (November 24, 2020) is carried out assuming the last growth rate computed.

### Reproduction number

The quantities *R*_*t*_ and *k*_*G*_(*t*) are related to each other through the Euler–Lotka equation, 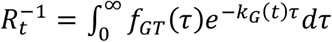, which considers 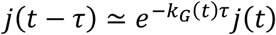 in the renewal equation 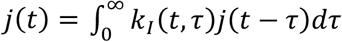. Generation times can generally be described through a gamma distribution 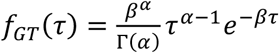 with 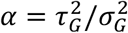 and 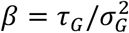, which leads to *R*_*t*_ = (1 + *k*_*G*_(*t*)/*β*)^*α*^ for *k*_*G*_(*t*) > −*β* and *R*_*t*_ = 0 for *k*_*G*_(*t*) ≤ −*β*. In the case of the exponentially distributed limit (*α* ≃ 1) or small values of *k*_*G*_(*t*)/*β*, it simplifies to *R*_*t*_ = 1 + *k*_*G*_(*t*)*τ*_*G*_ for *k*_*G*_(*t*) > −1/*τ*_*G*_ and *R*_*t*_ = 0 for *k*_*G*_(*t*) ≤ −1/*τ*_*G*_.

References are supplied in the main text of the manuscript.

## SUPPLEMENTARY TABLE

**Table S1.**
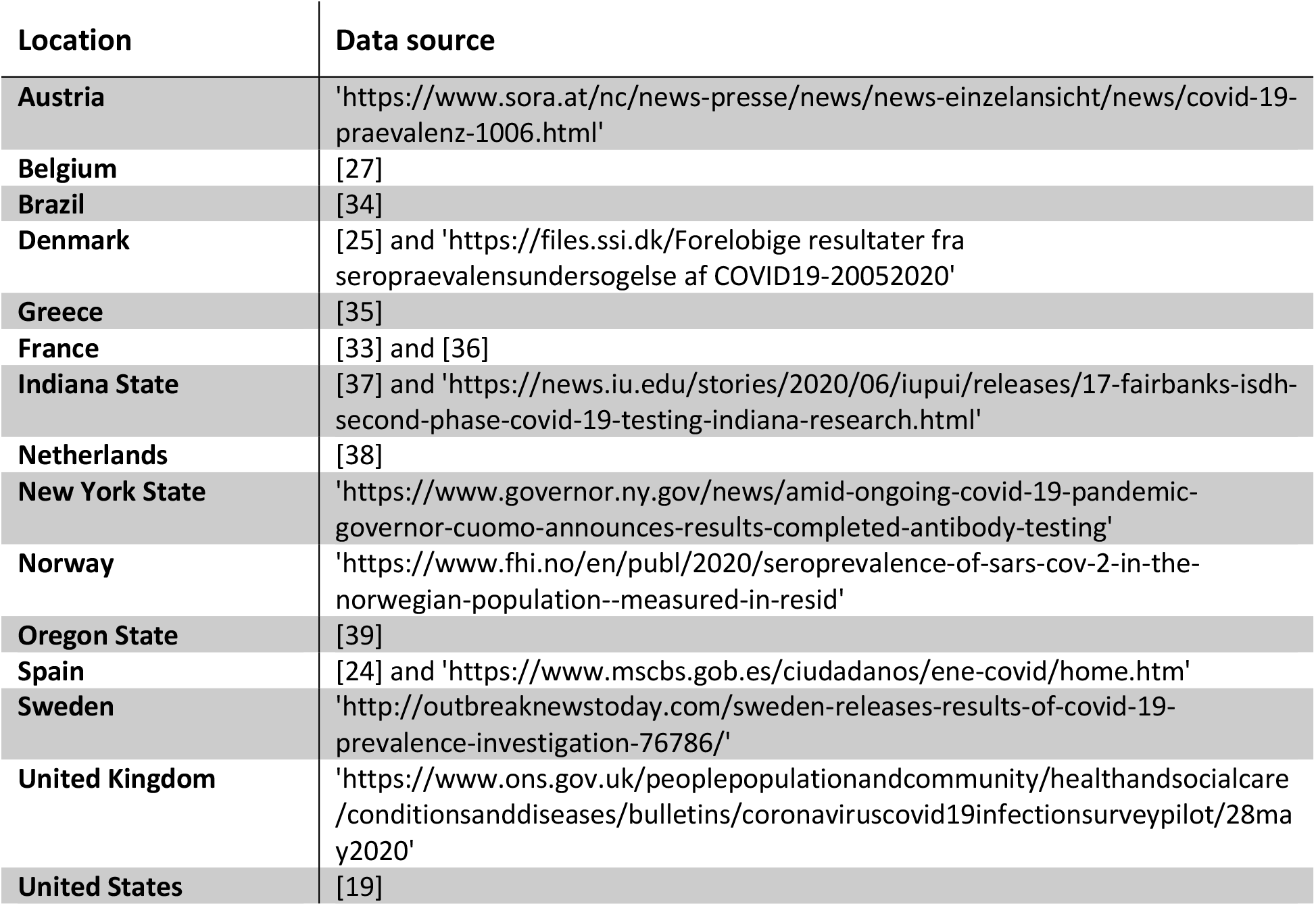
Sources for the prevalence data at a global scale. The sources include press releases, online preprints, and publications. Seroprevalence data for Belgium and Spain does not include late estimates that show clear effects of diminishing antibody levels after infection. Prevalence data for England was used as a proxy for the United Kingdom. One of the data points for France [33] is a theoretical estimate based on detailed data.

## SUPPLEMENTARY FIGURE

**Fig S1.**
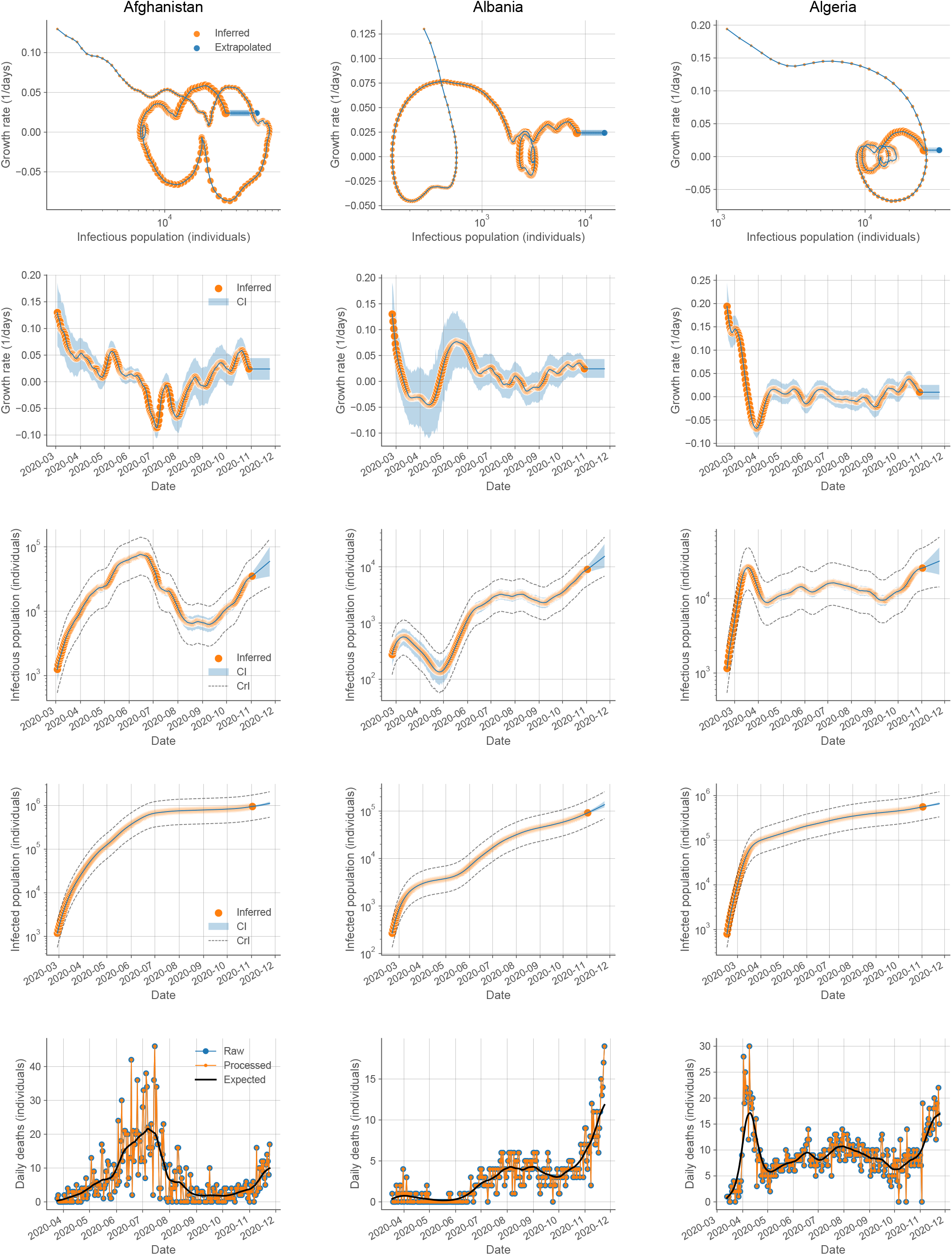
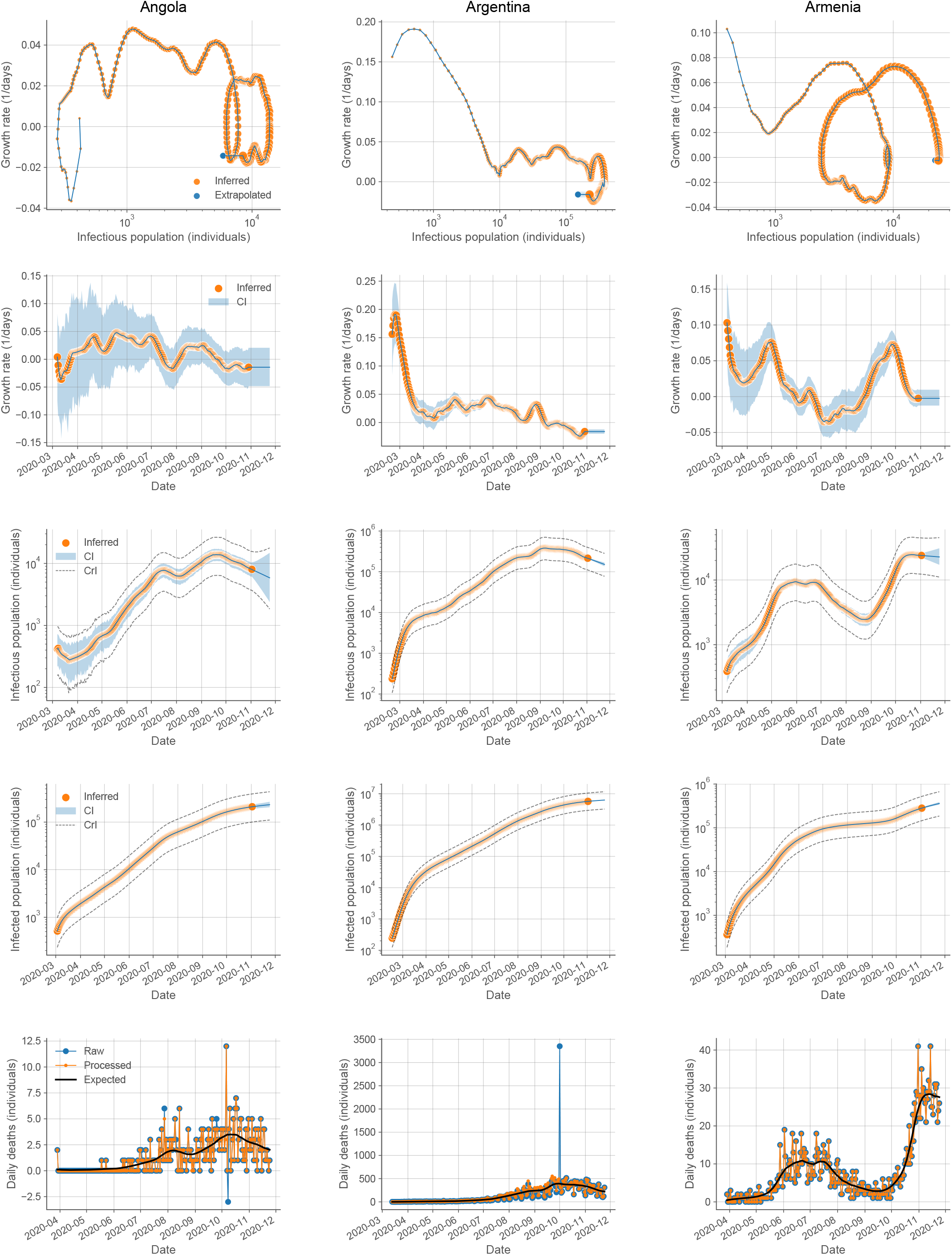
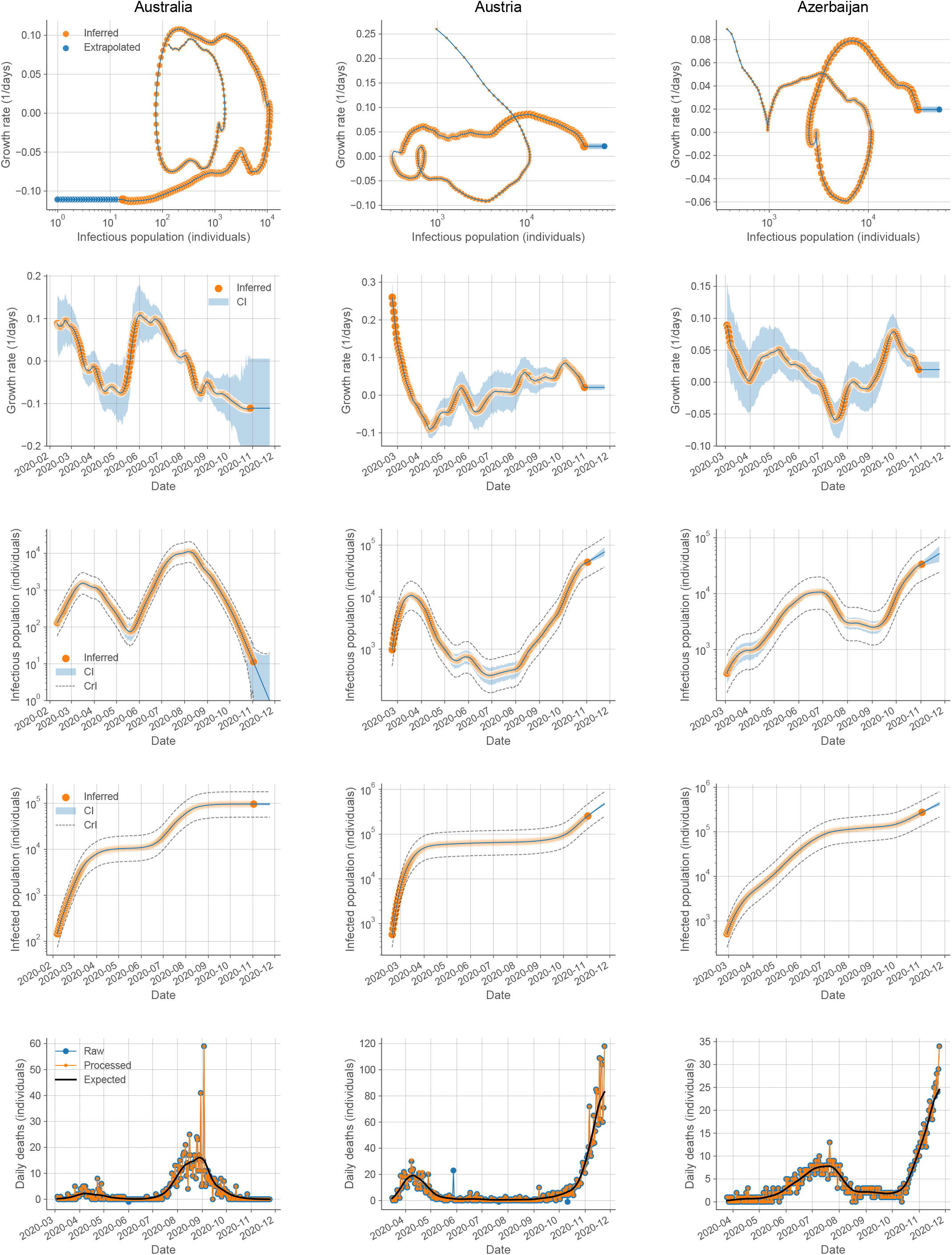
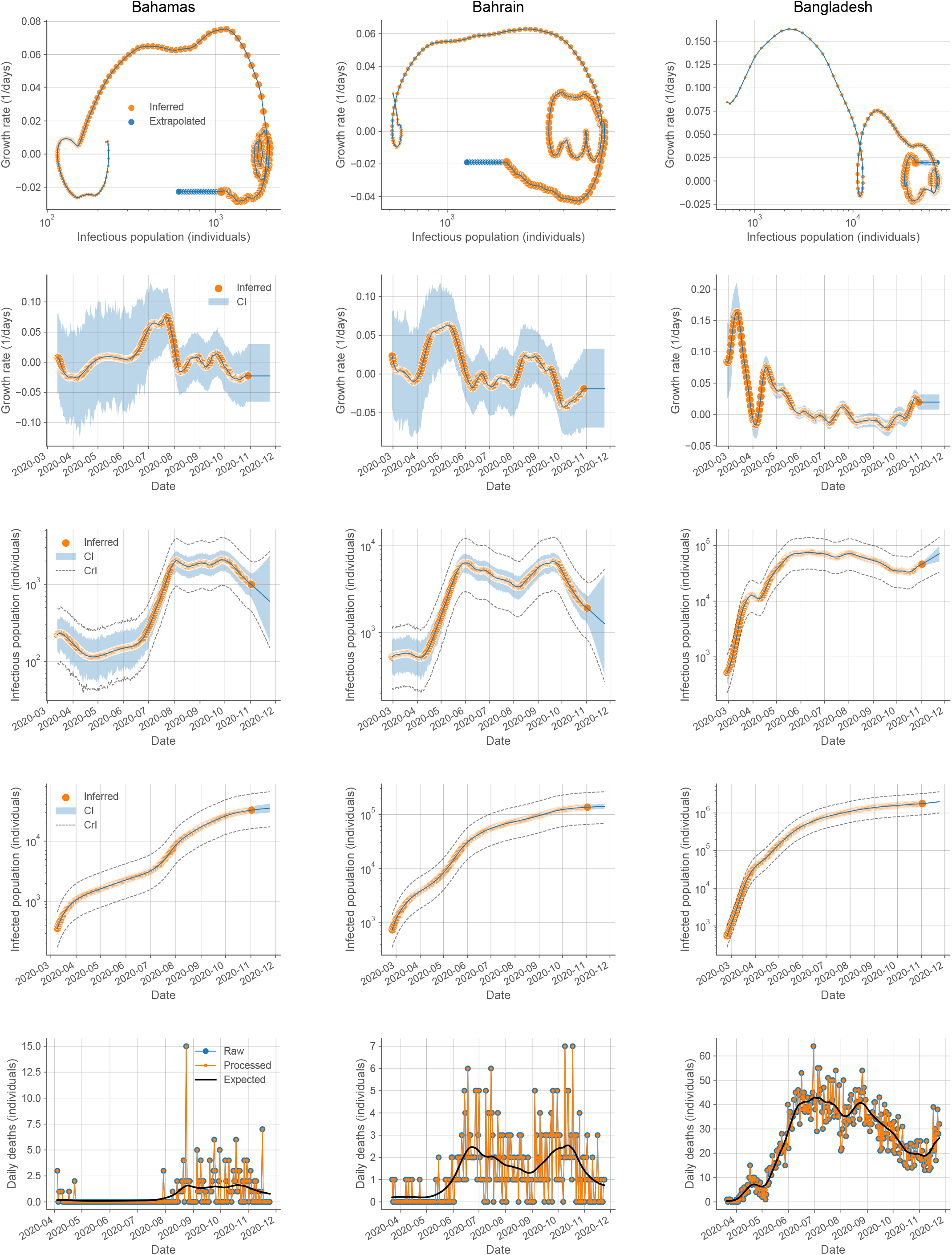
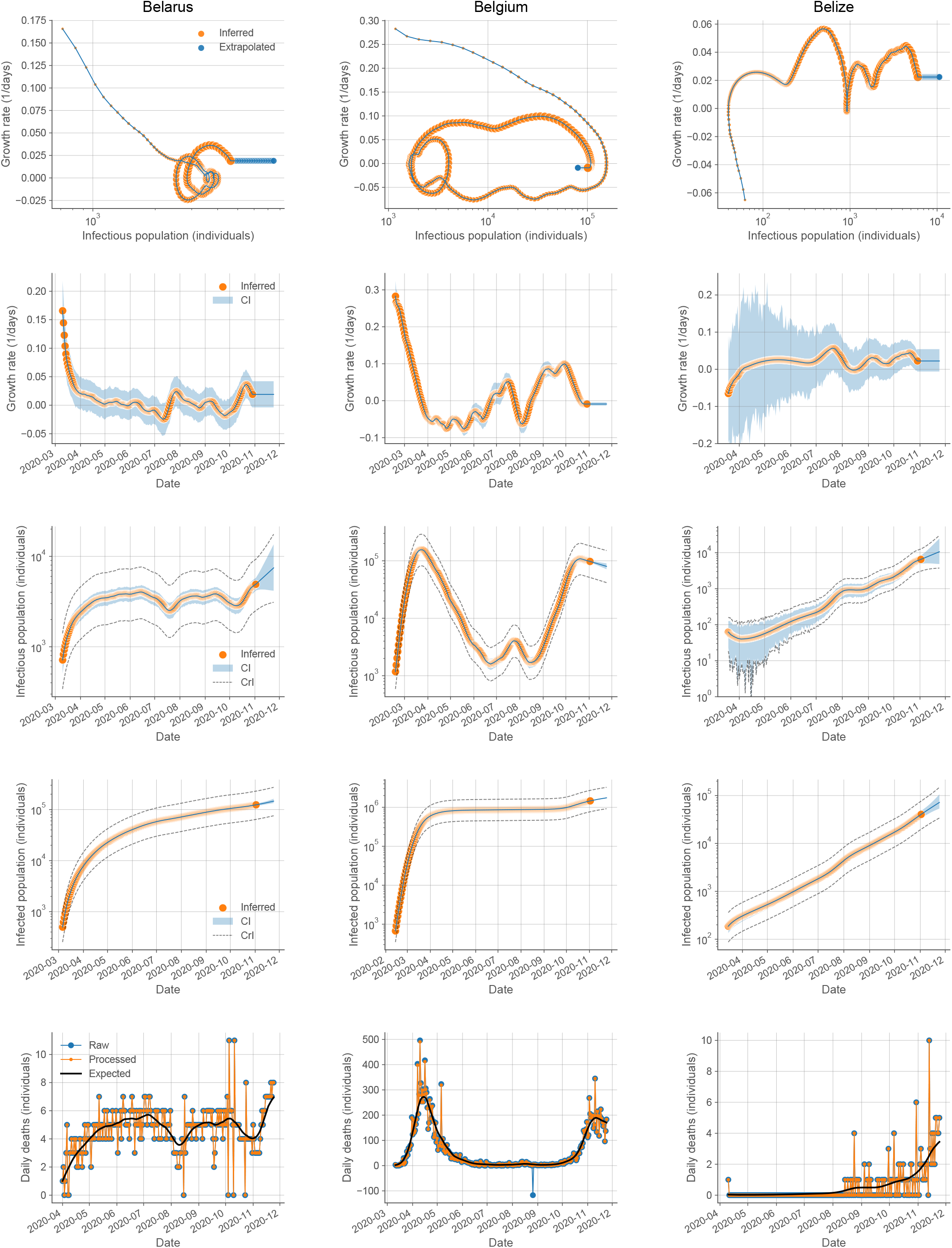
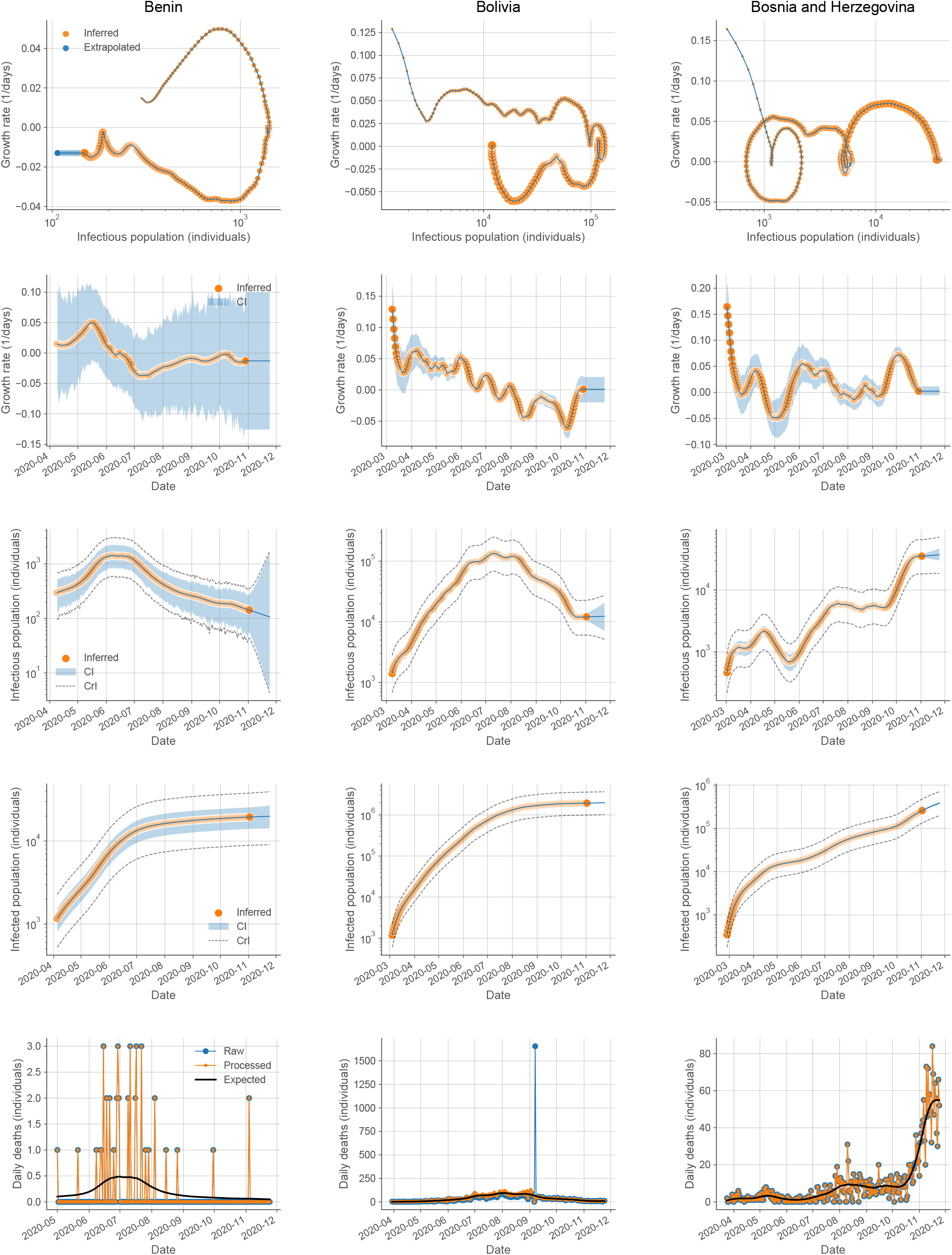
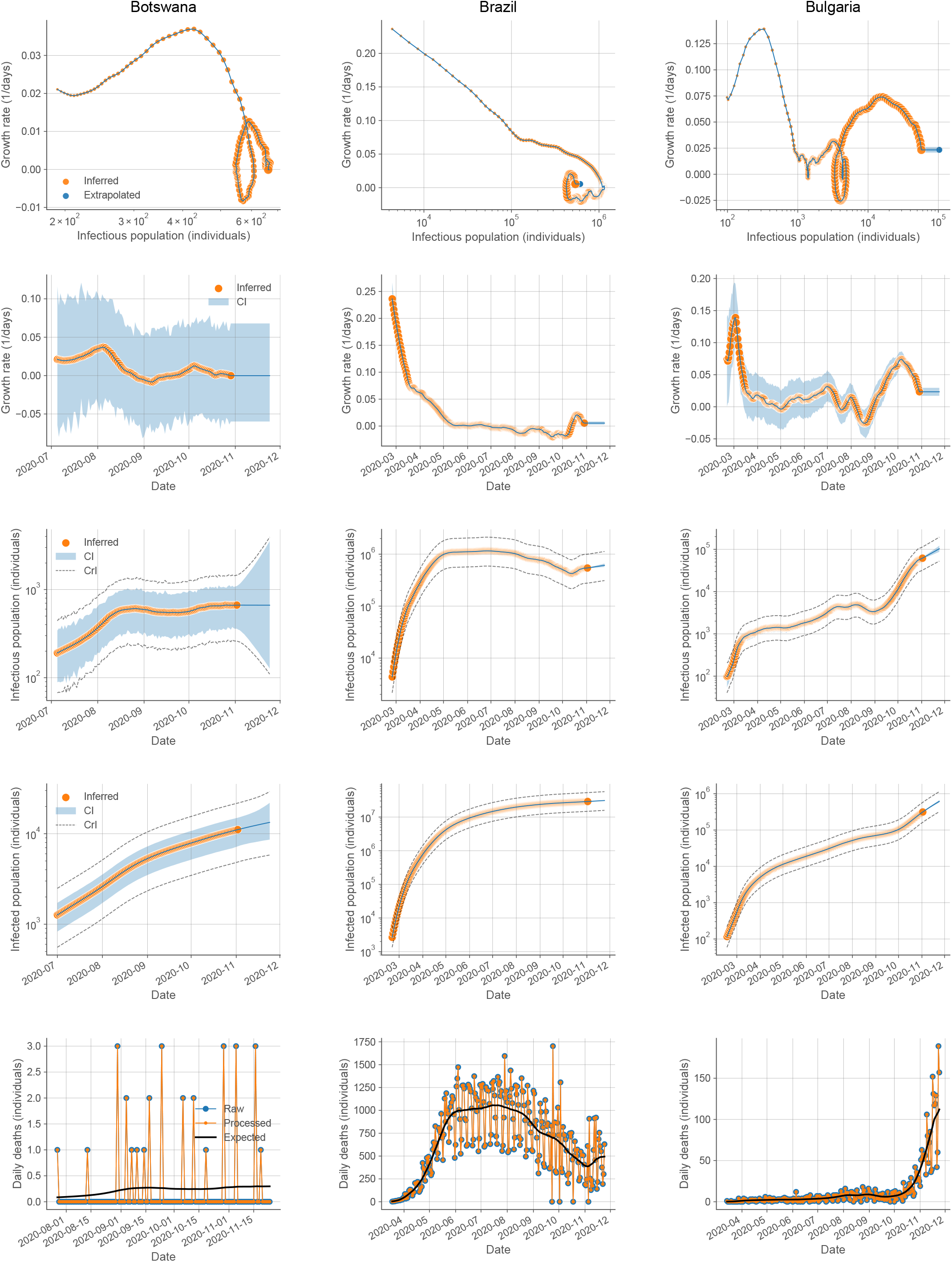
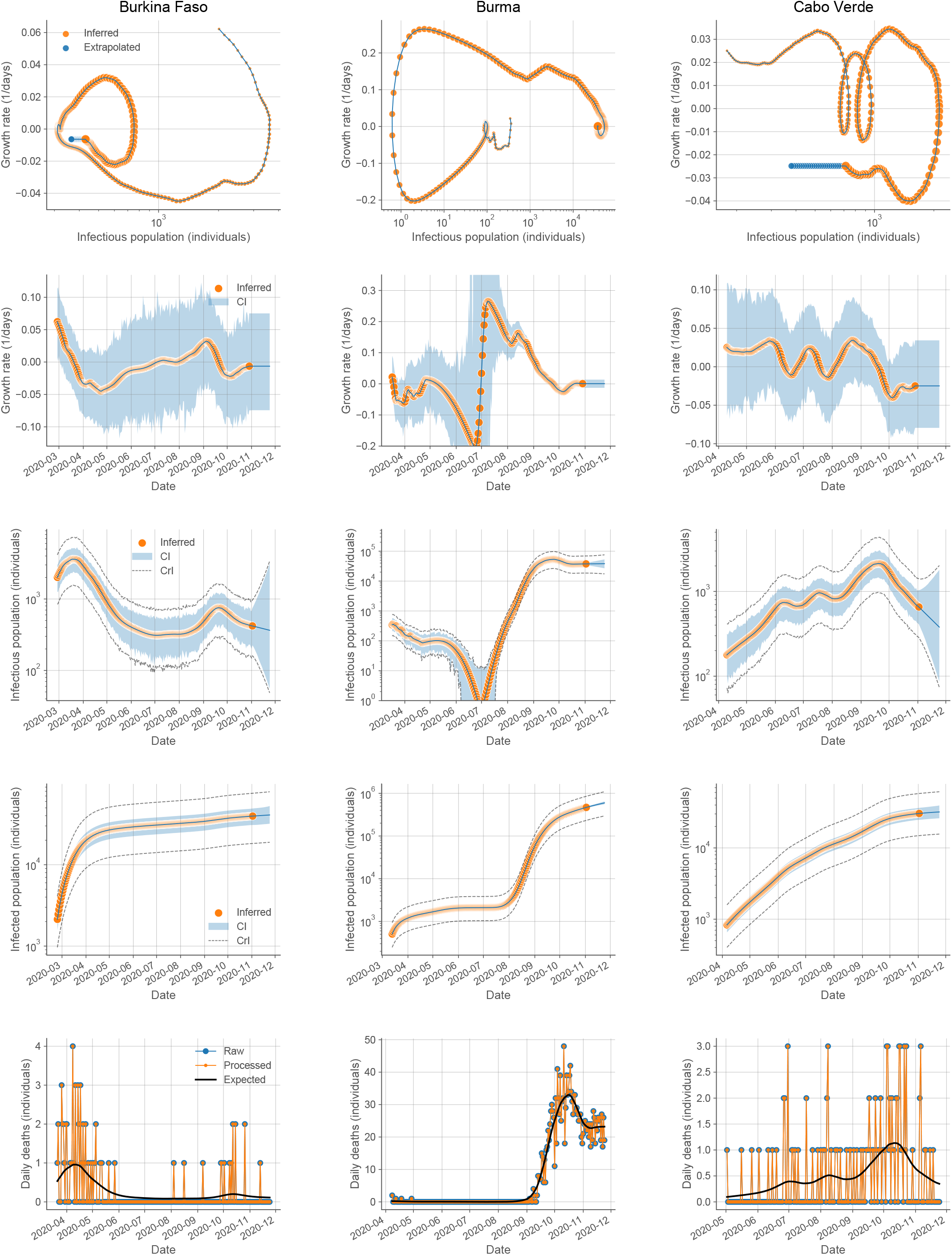
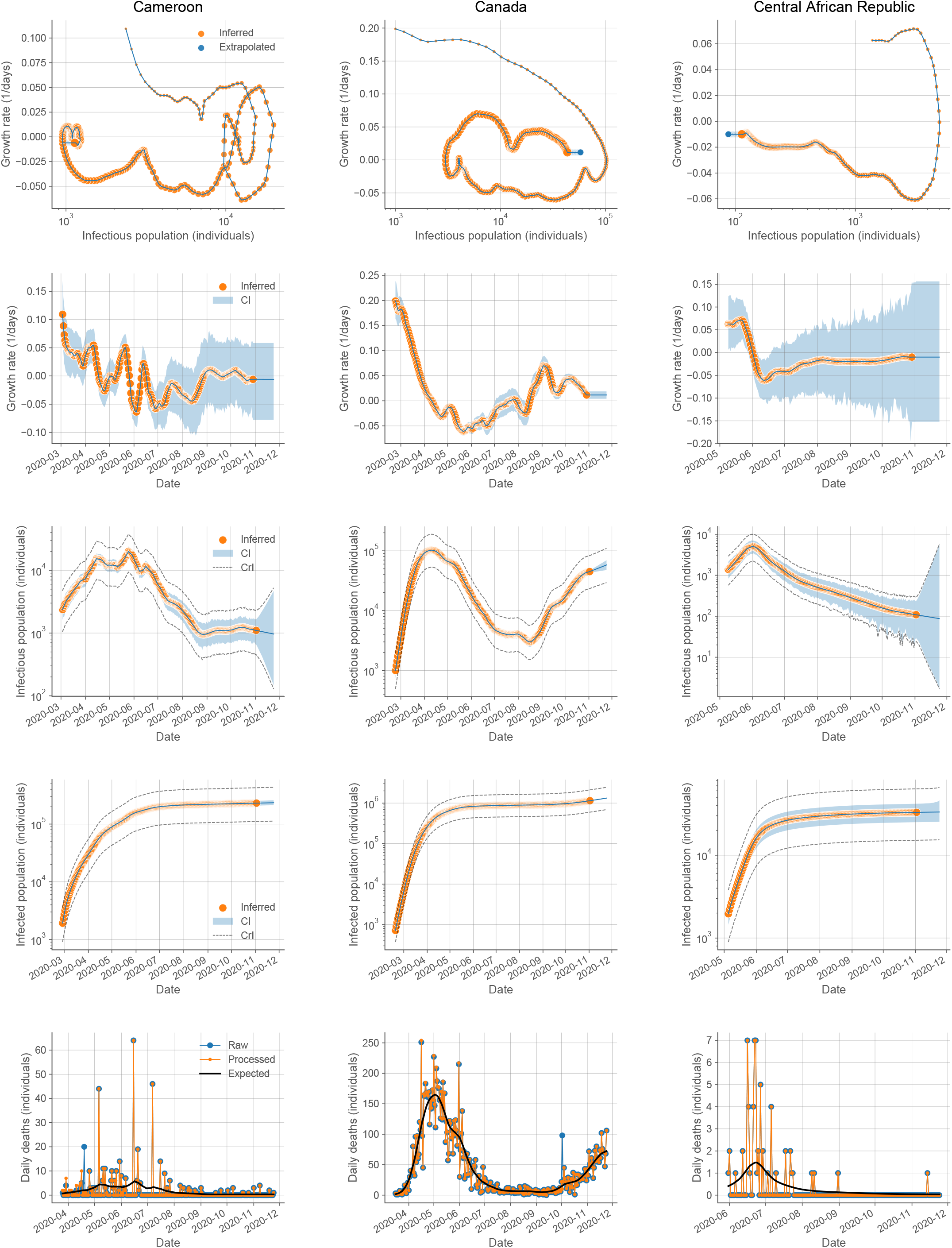
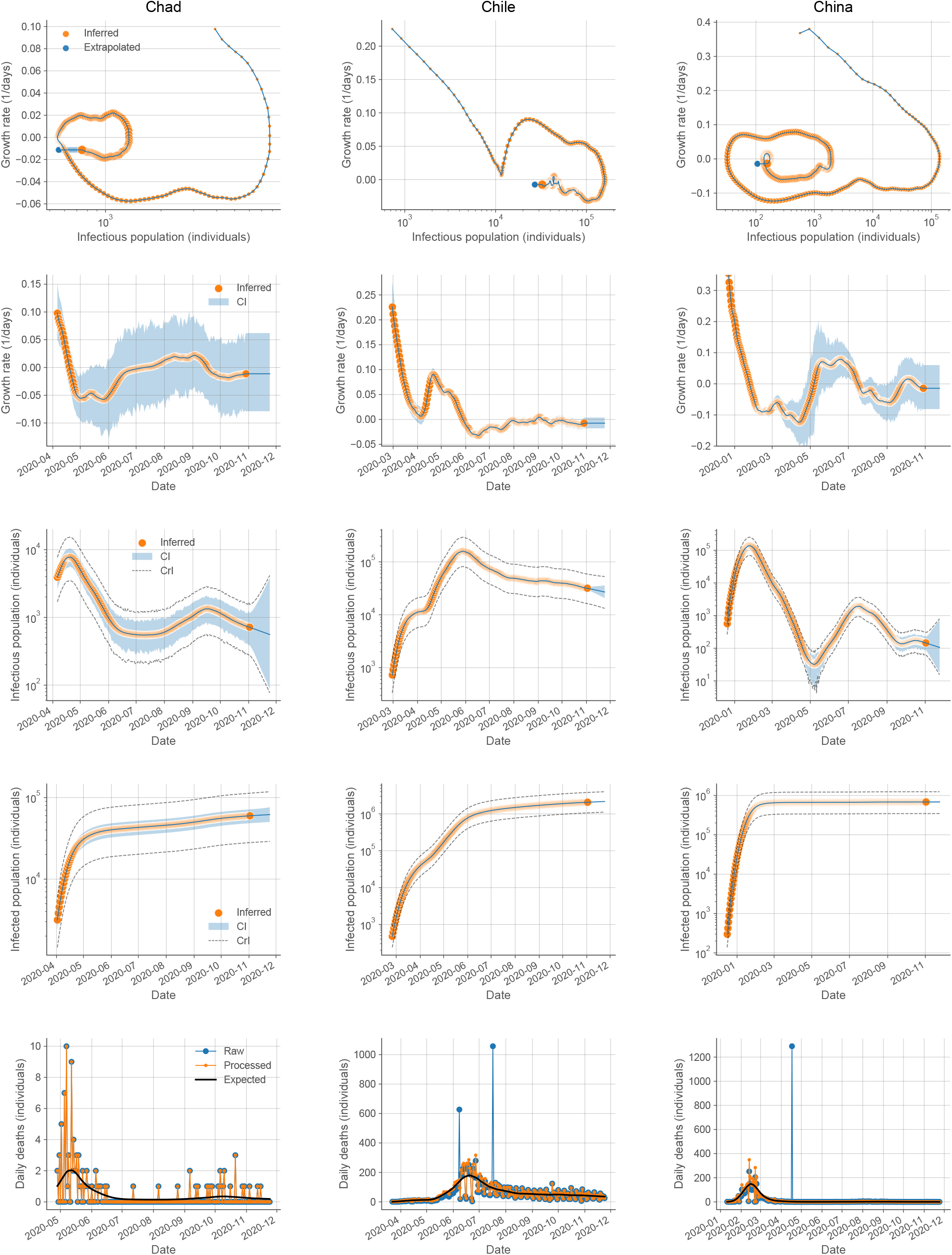
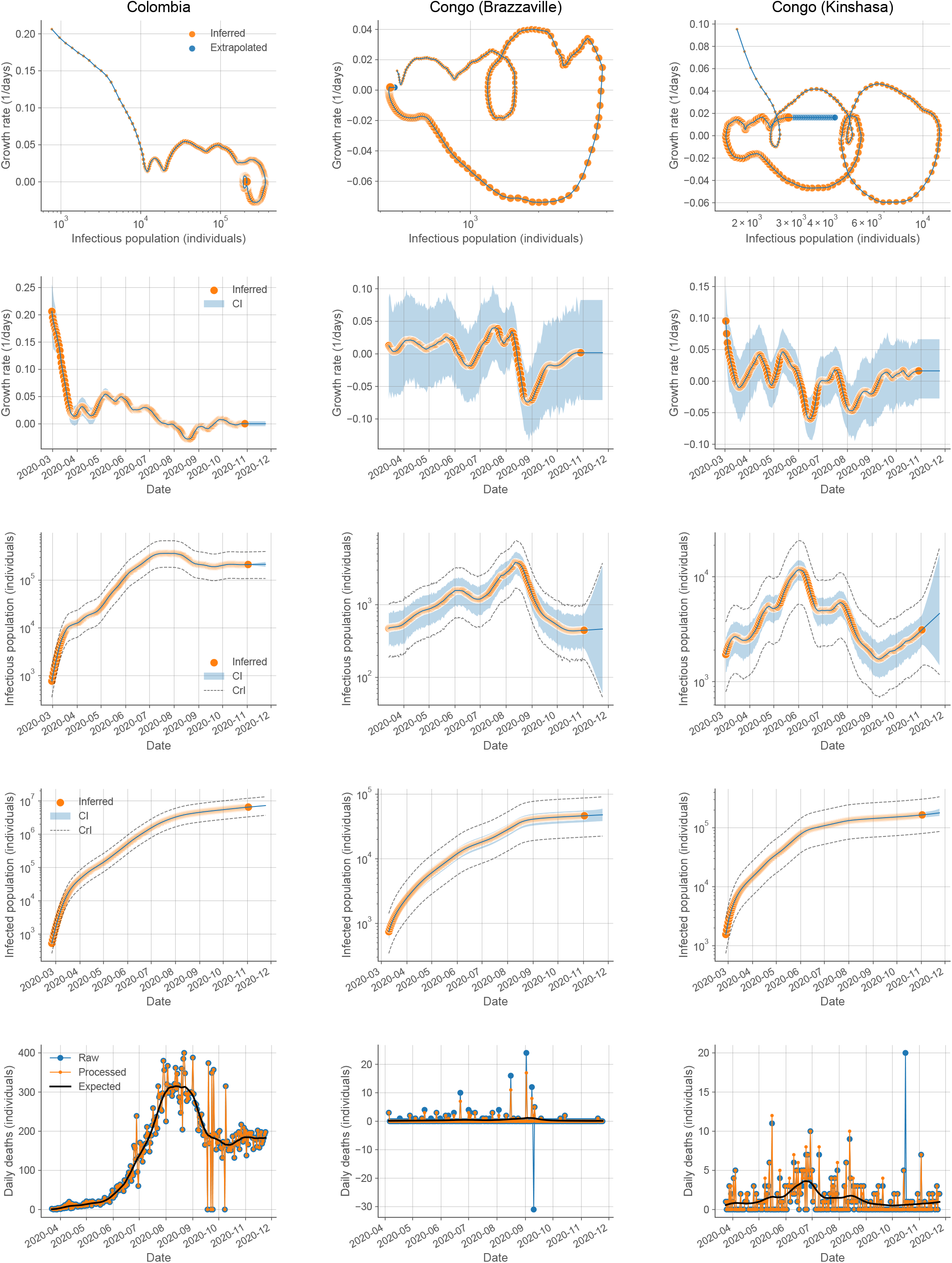
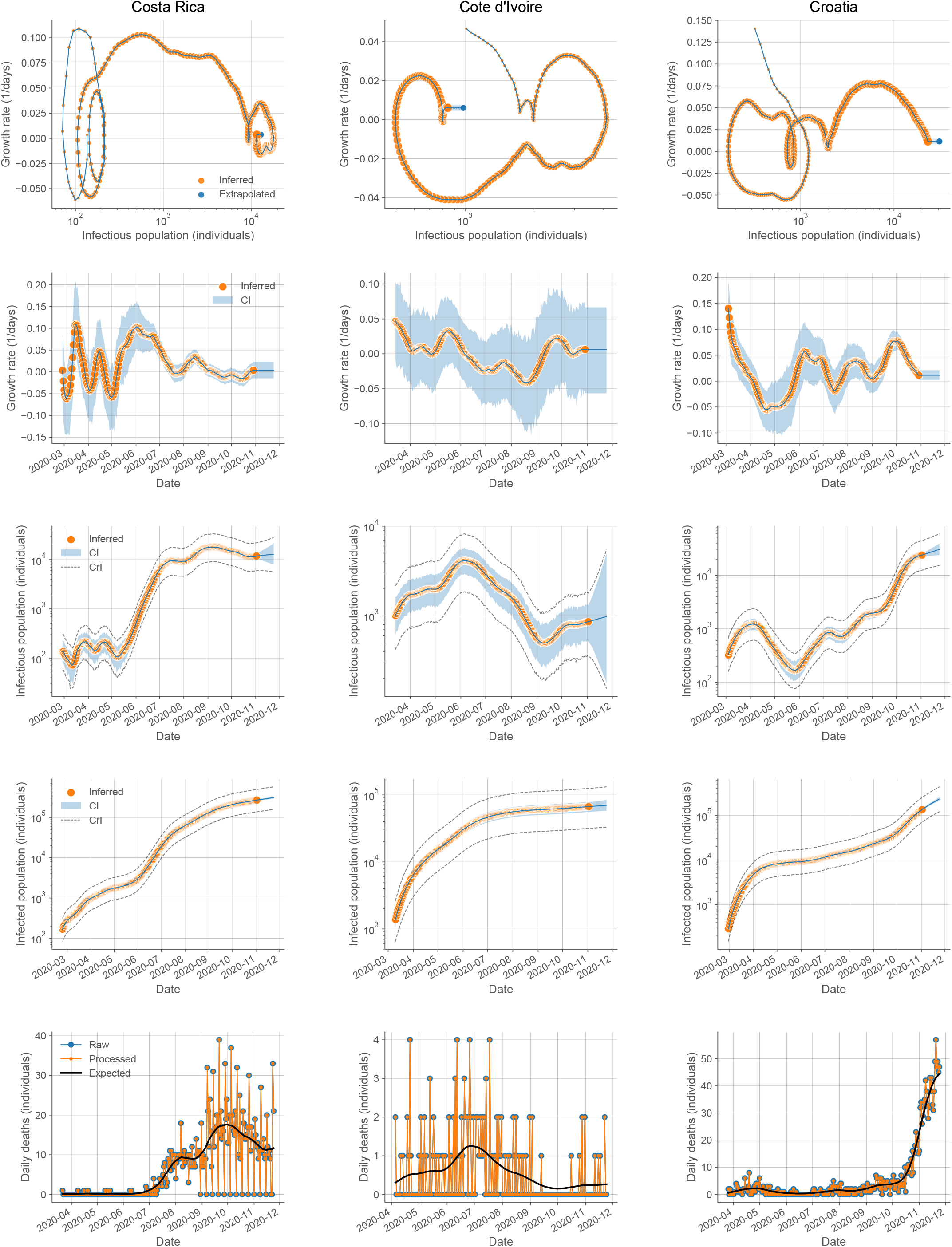
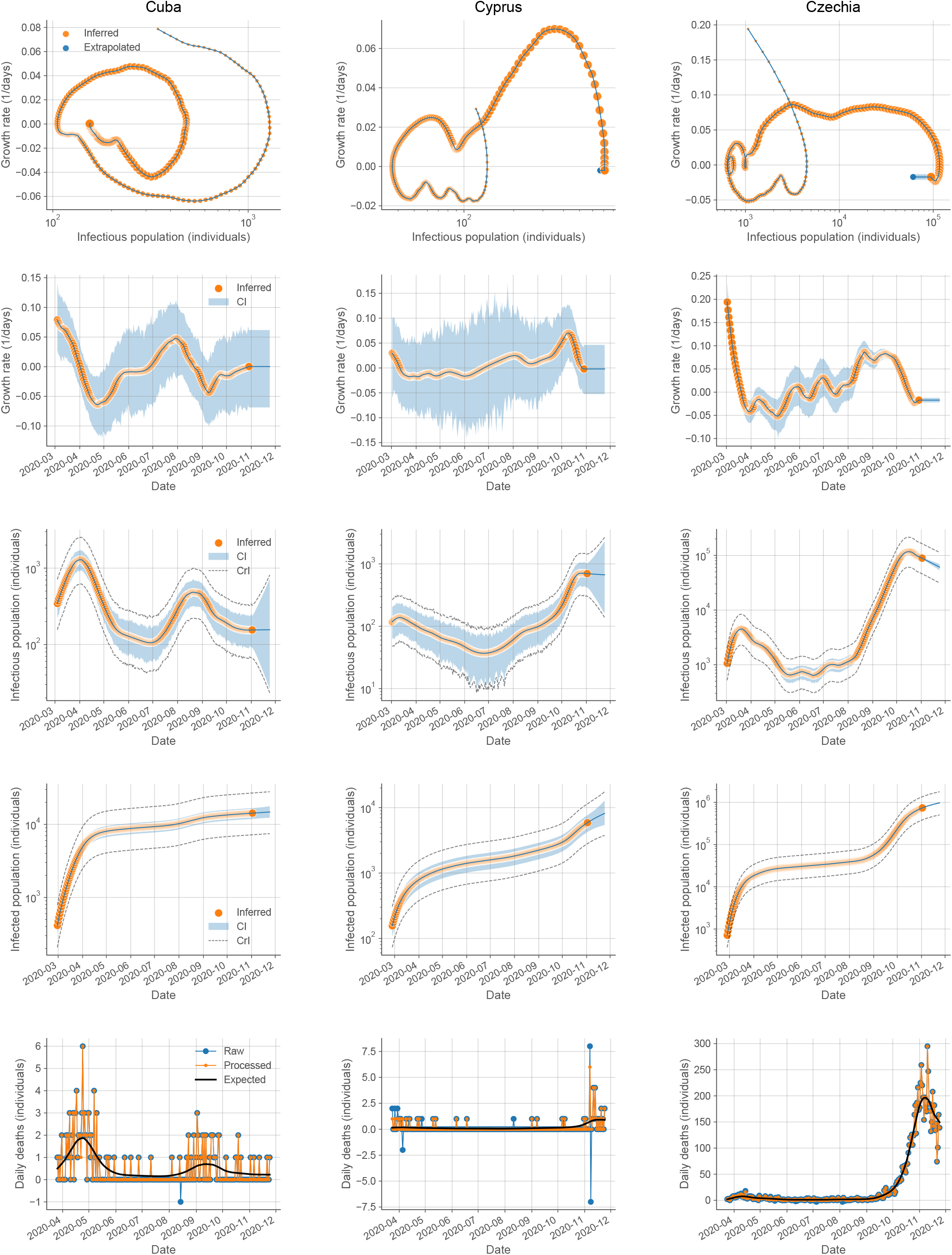
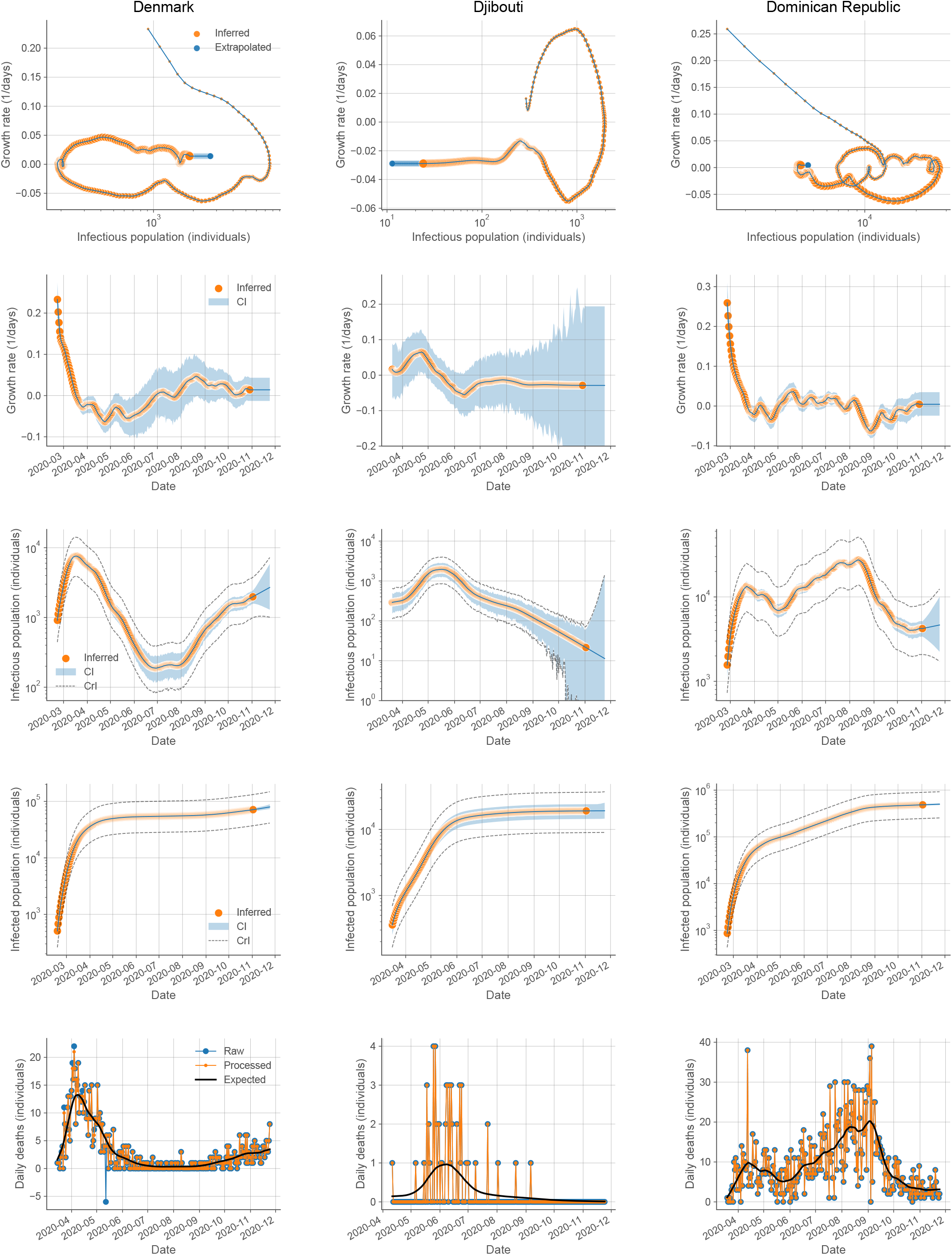
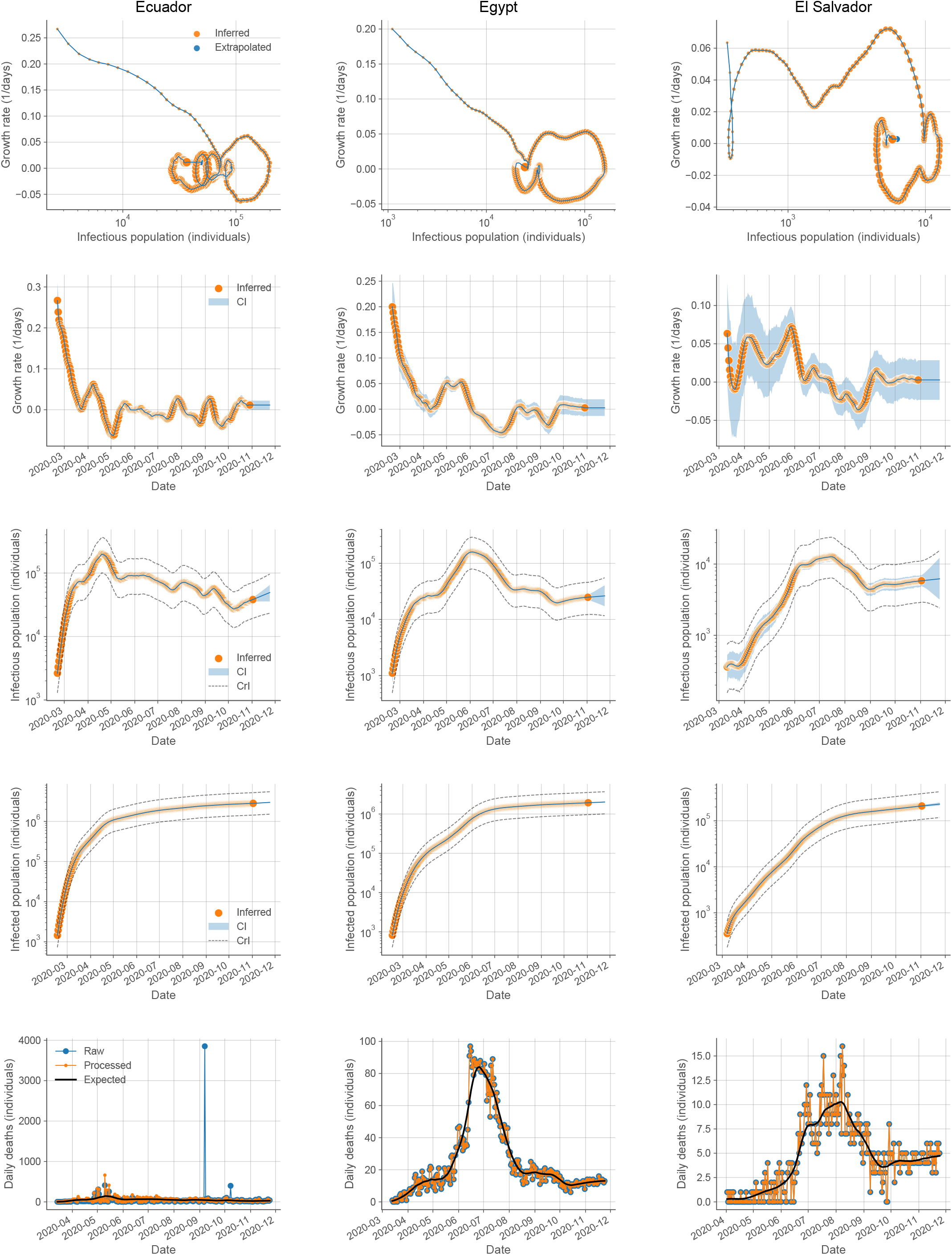
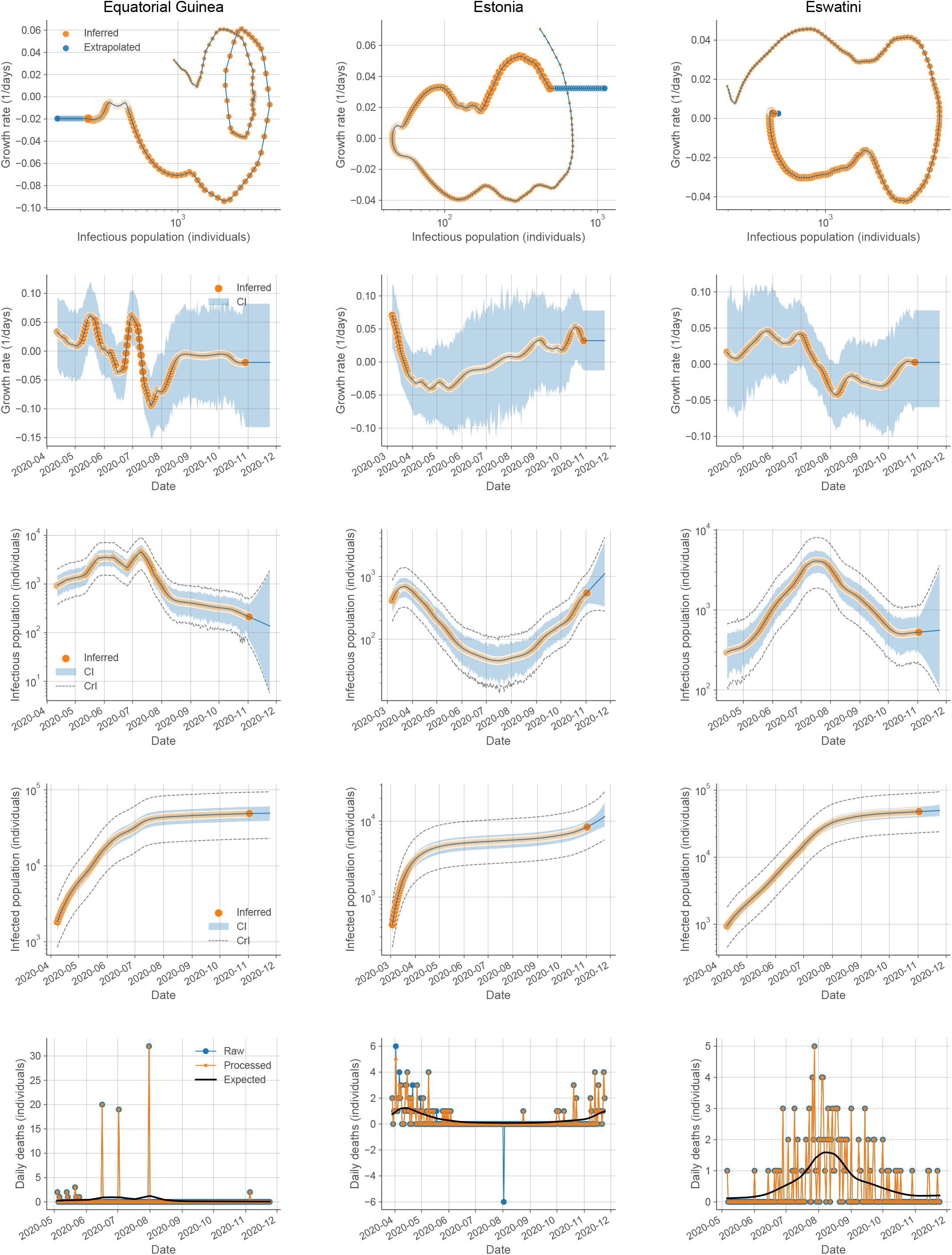
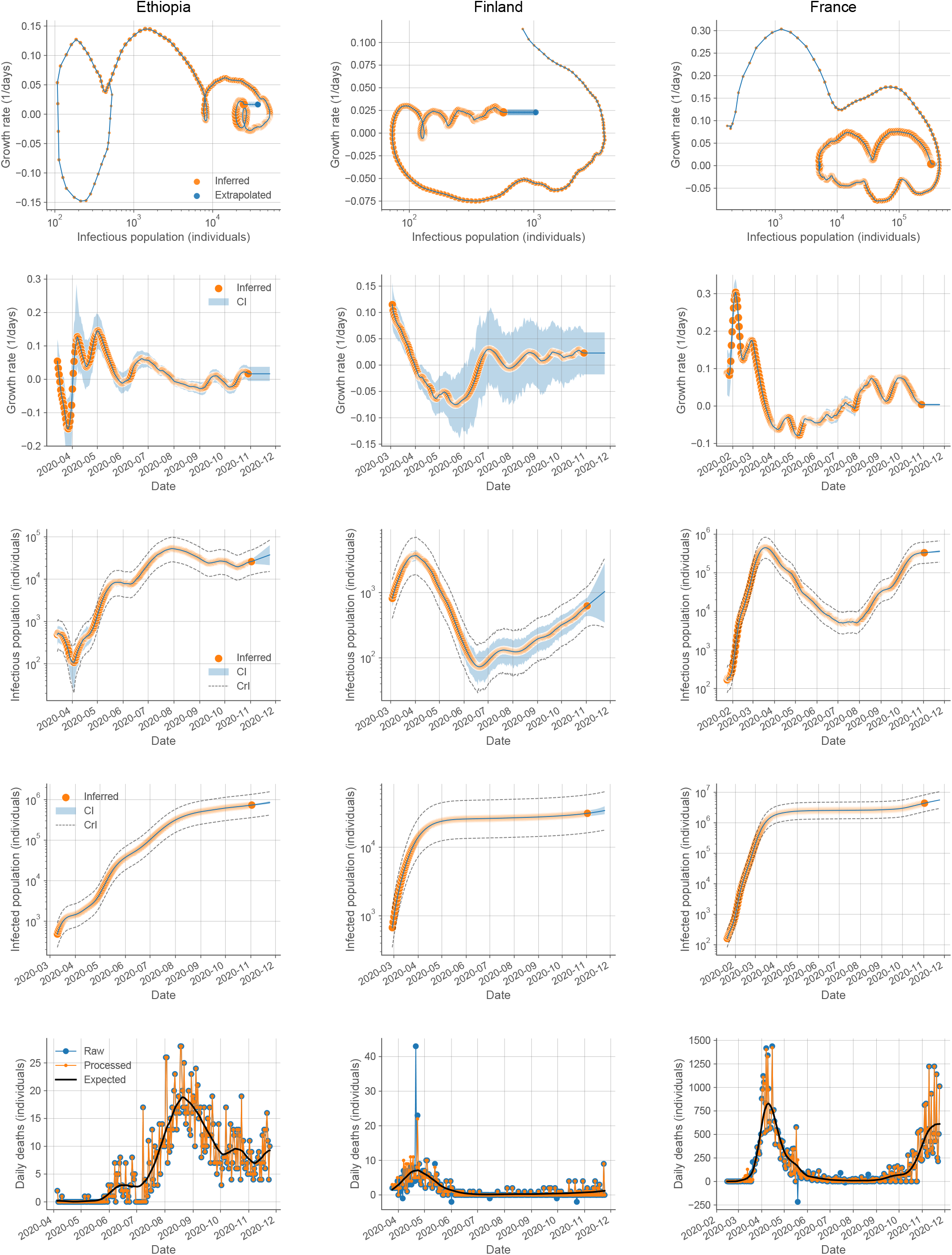
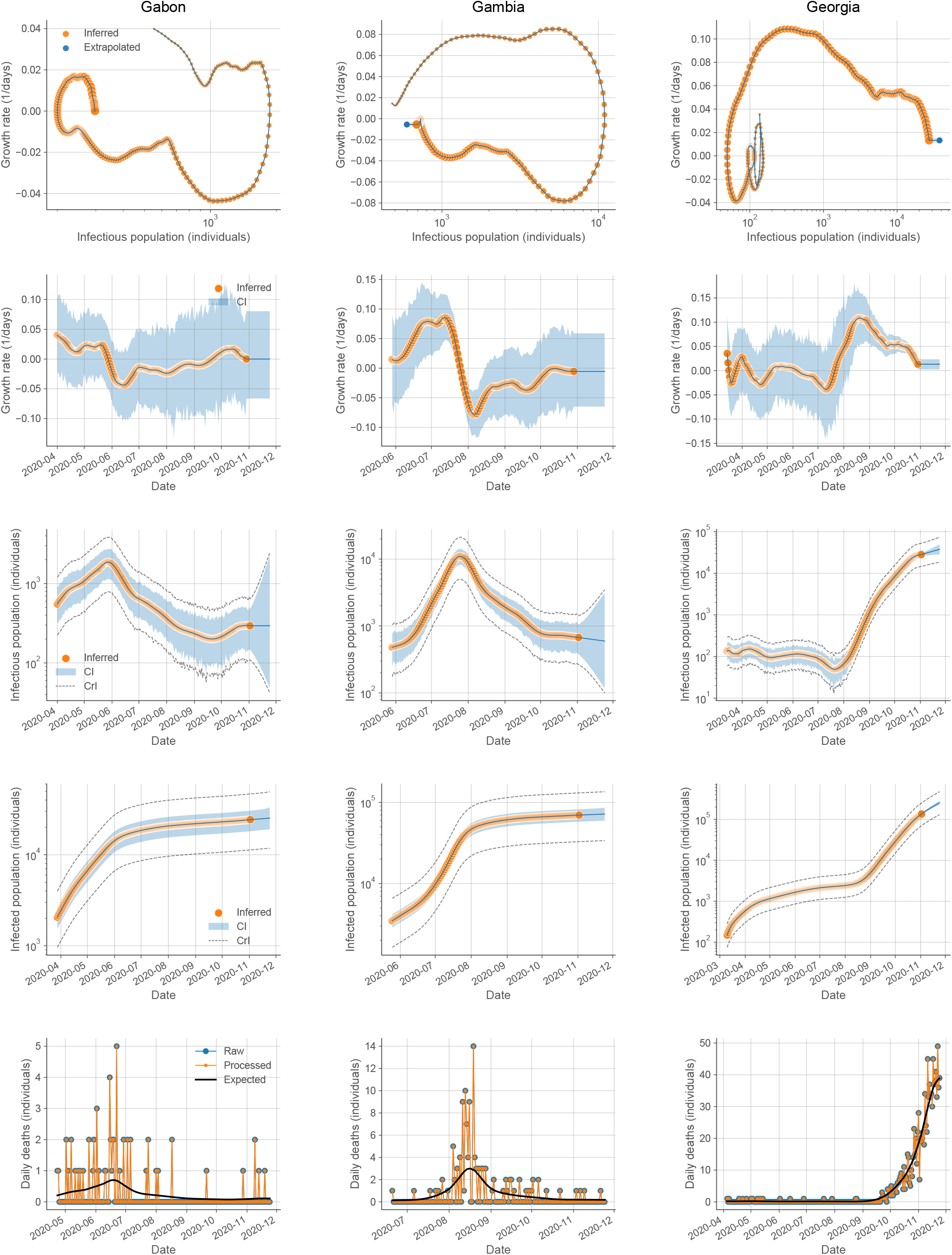
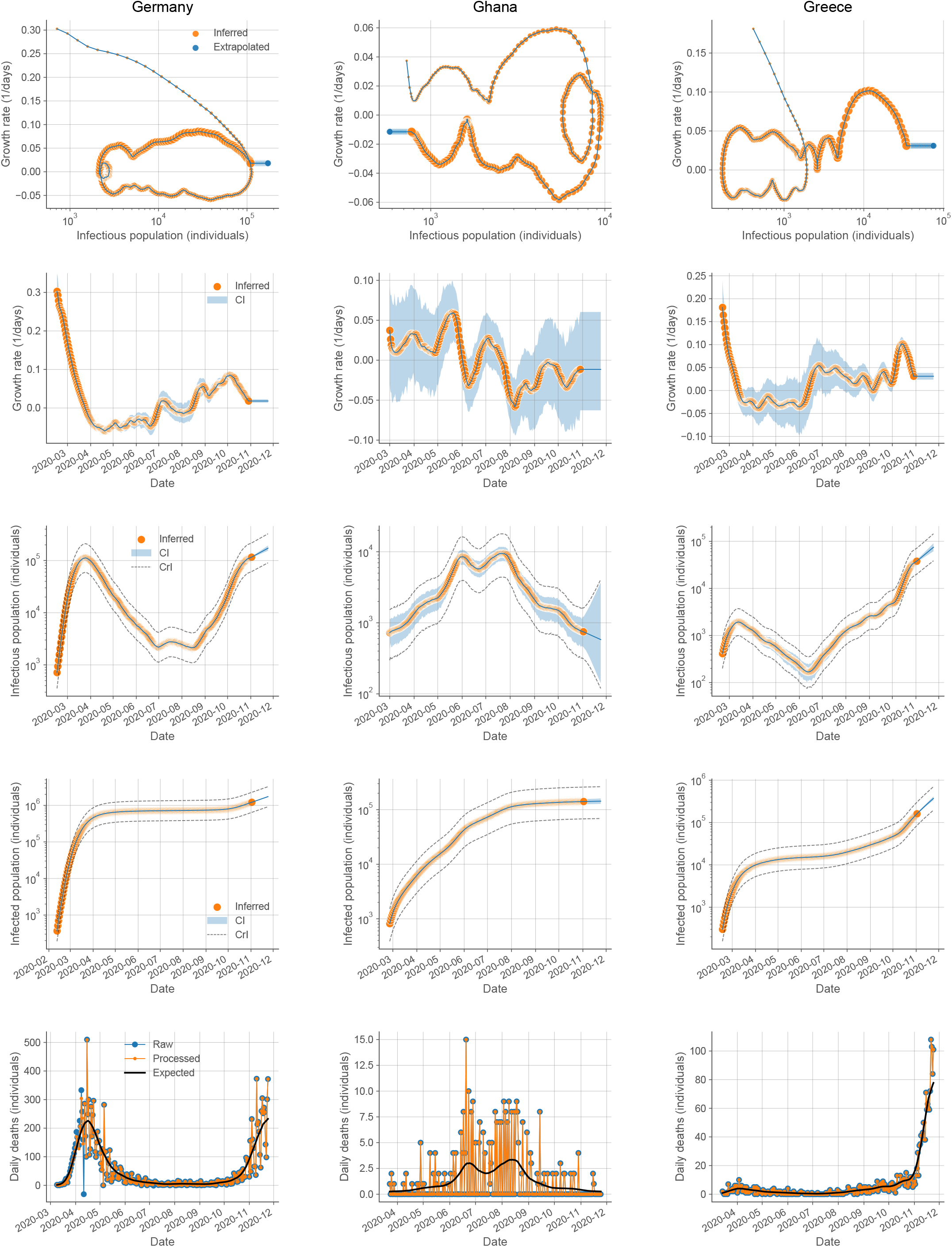
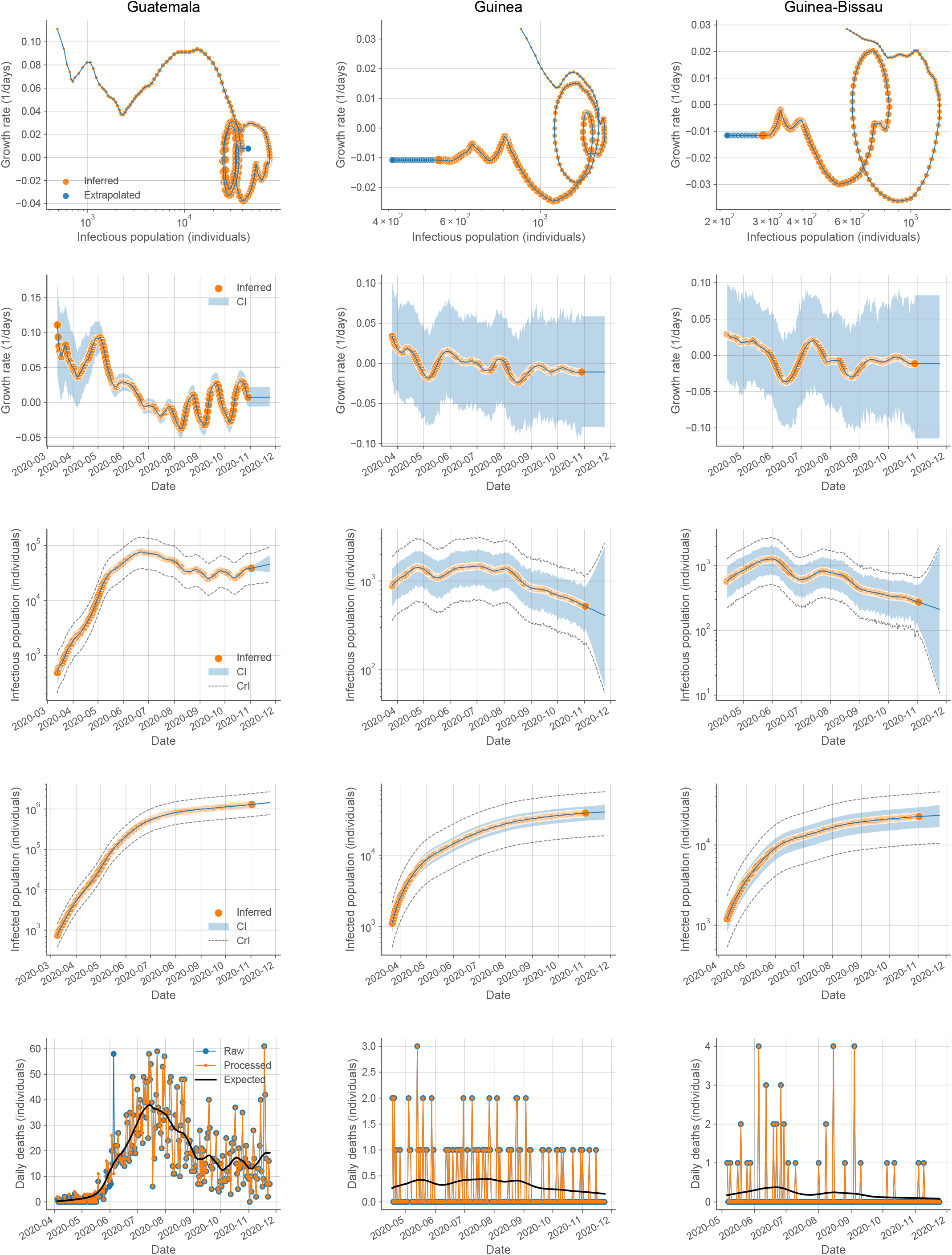
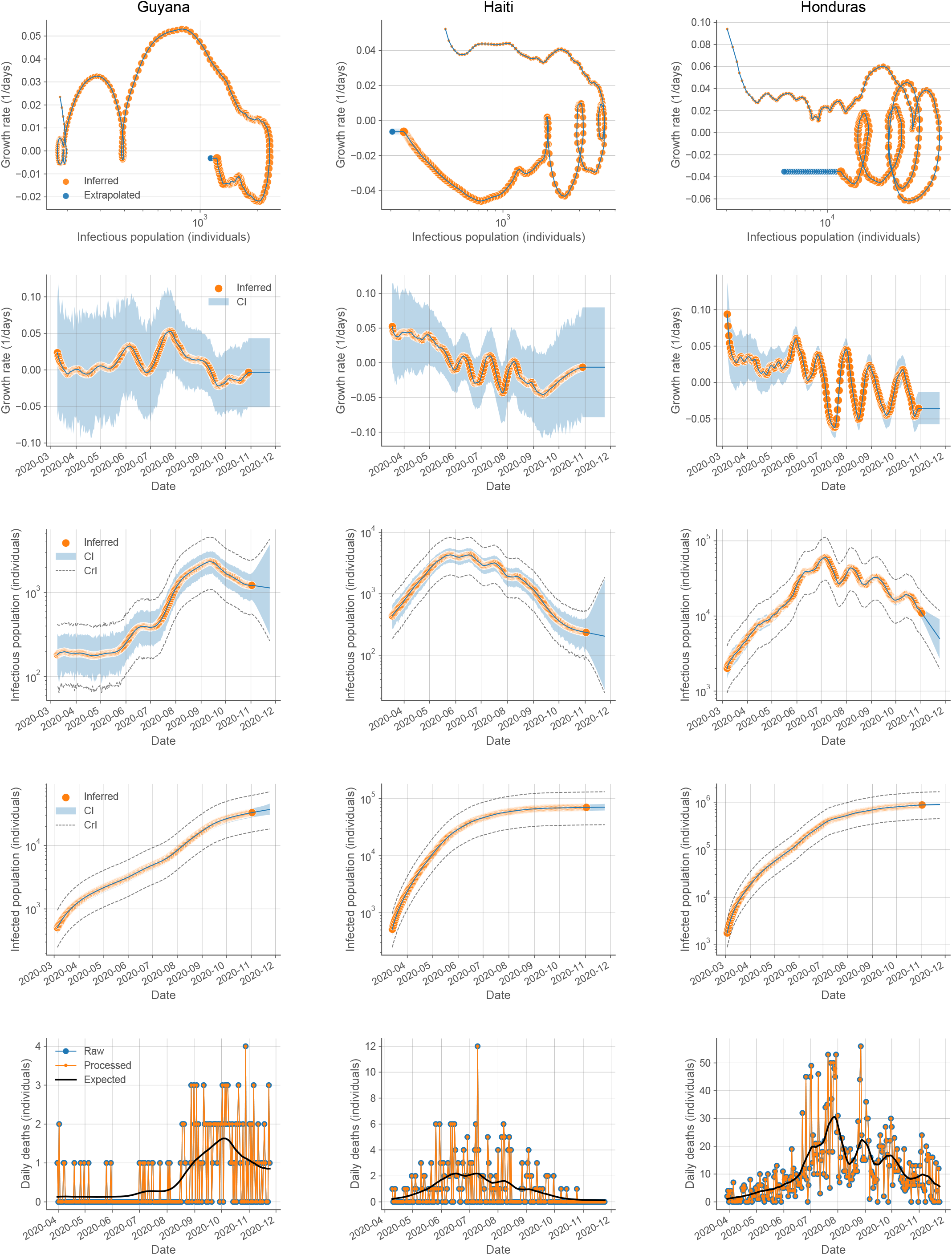
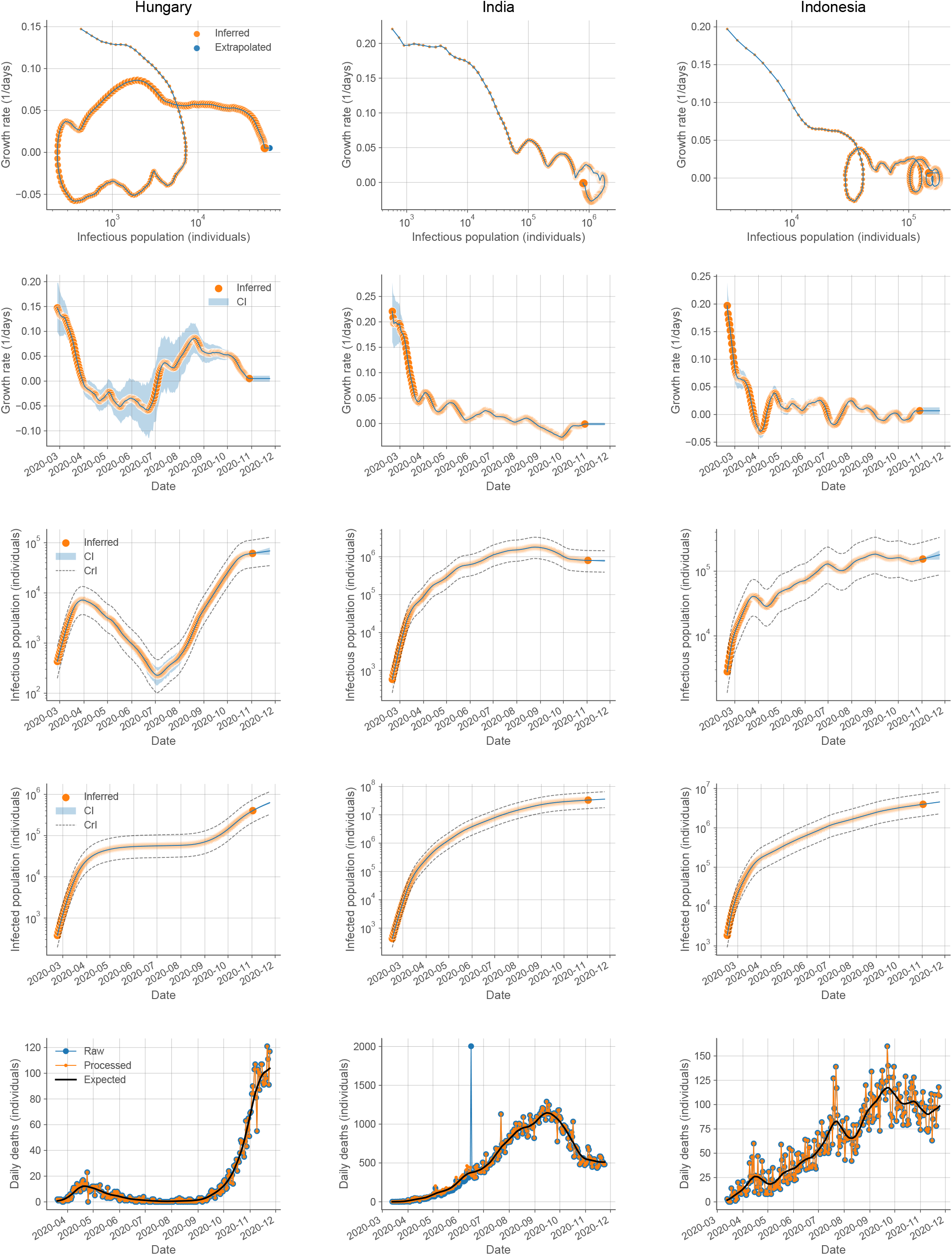
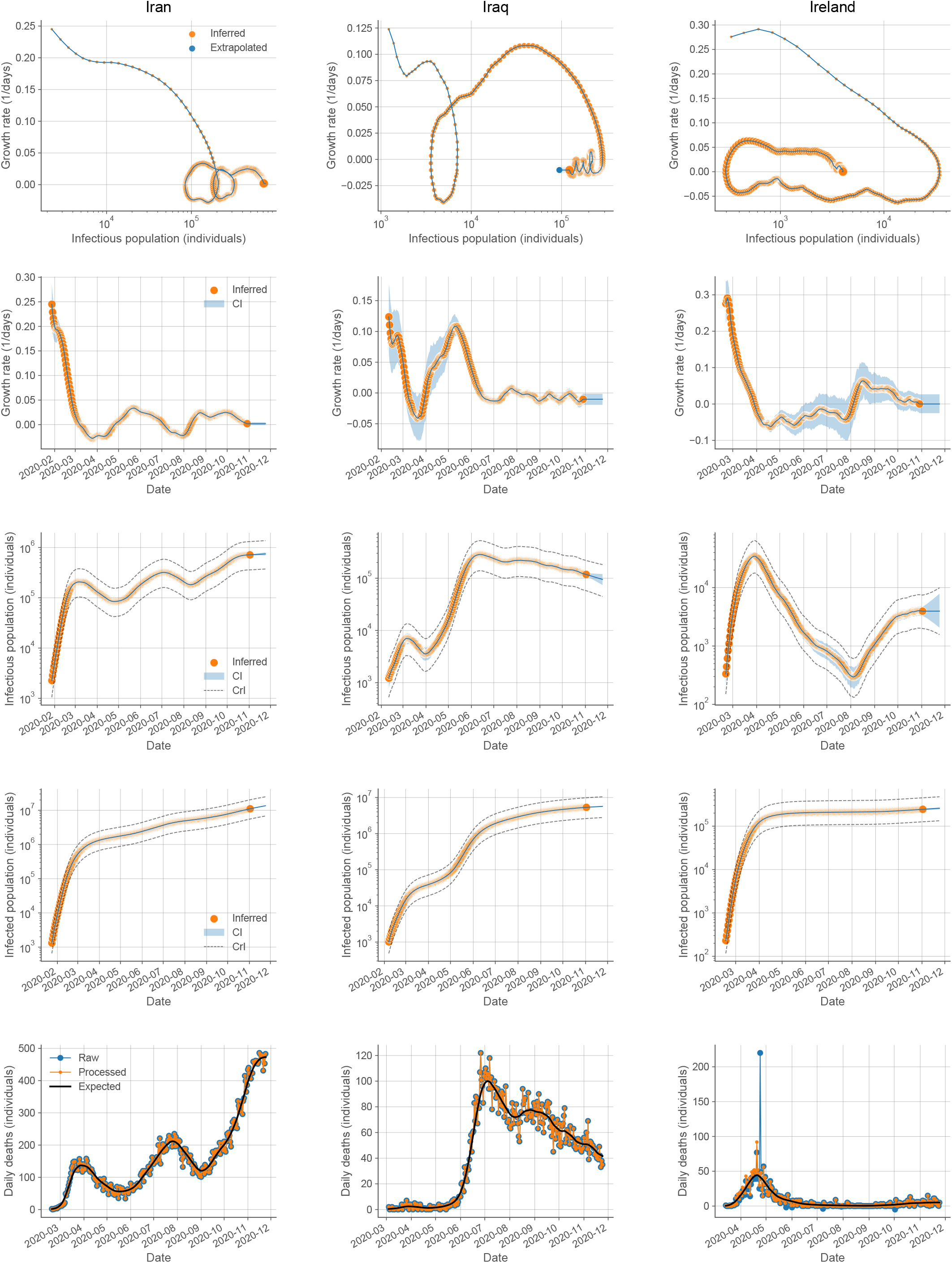
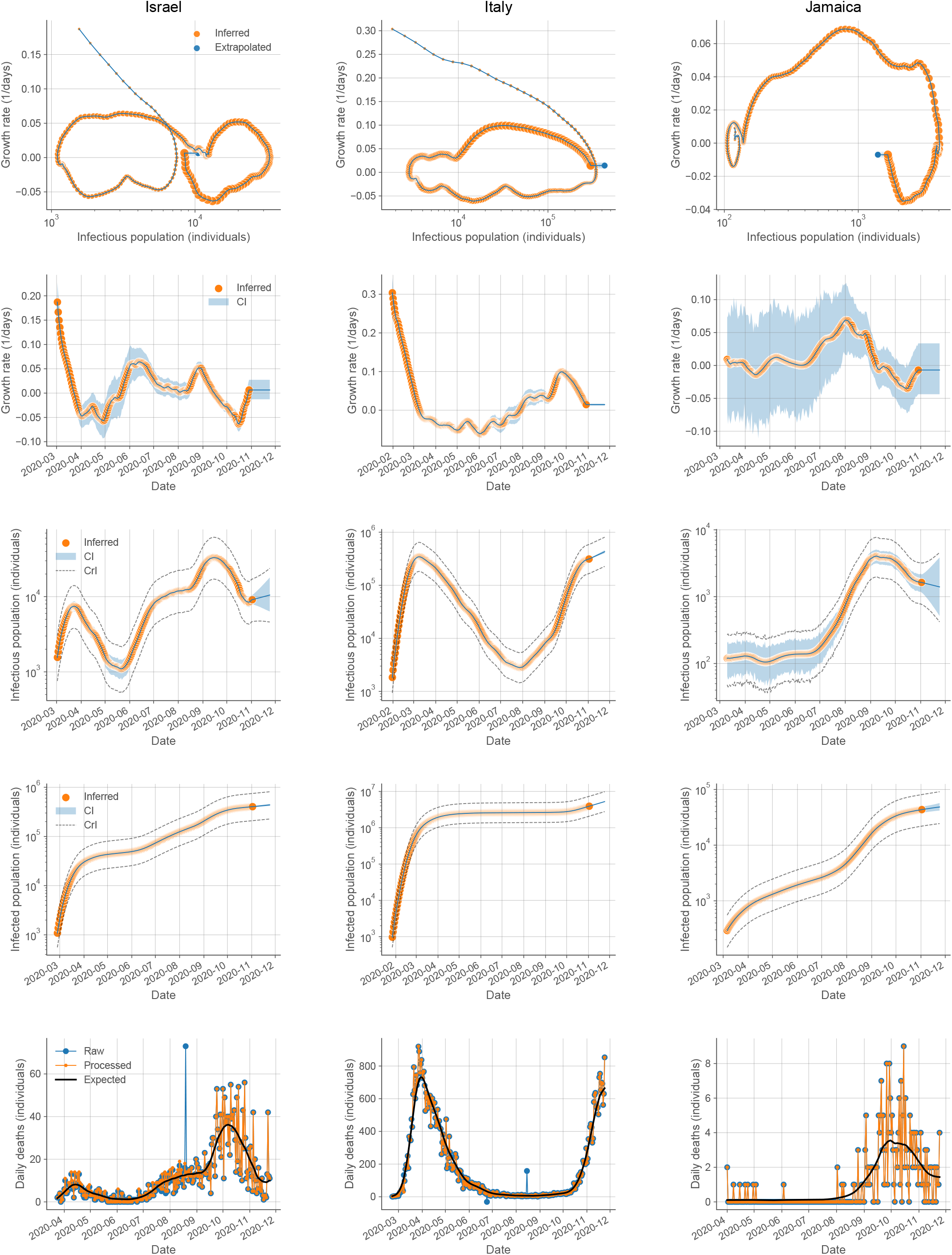
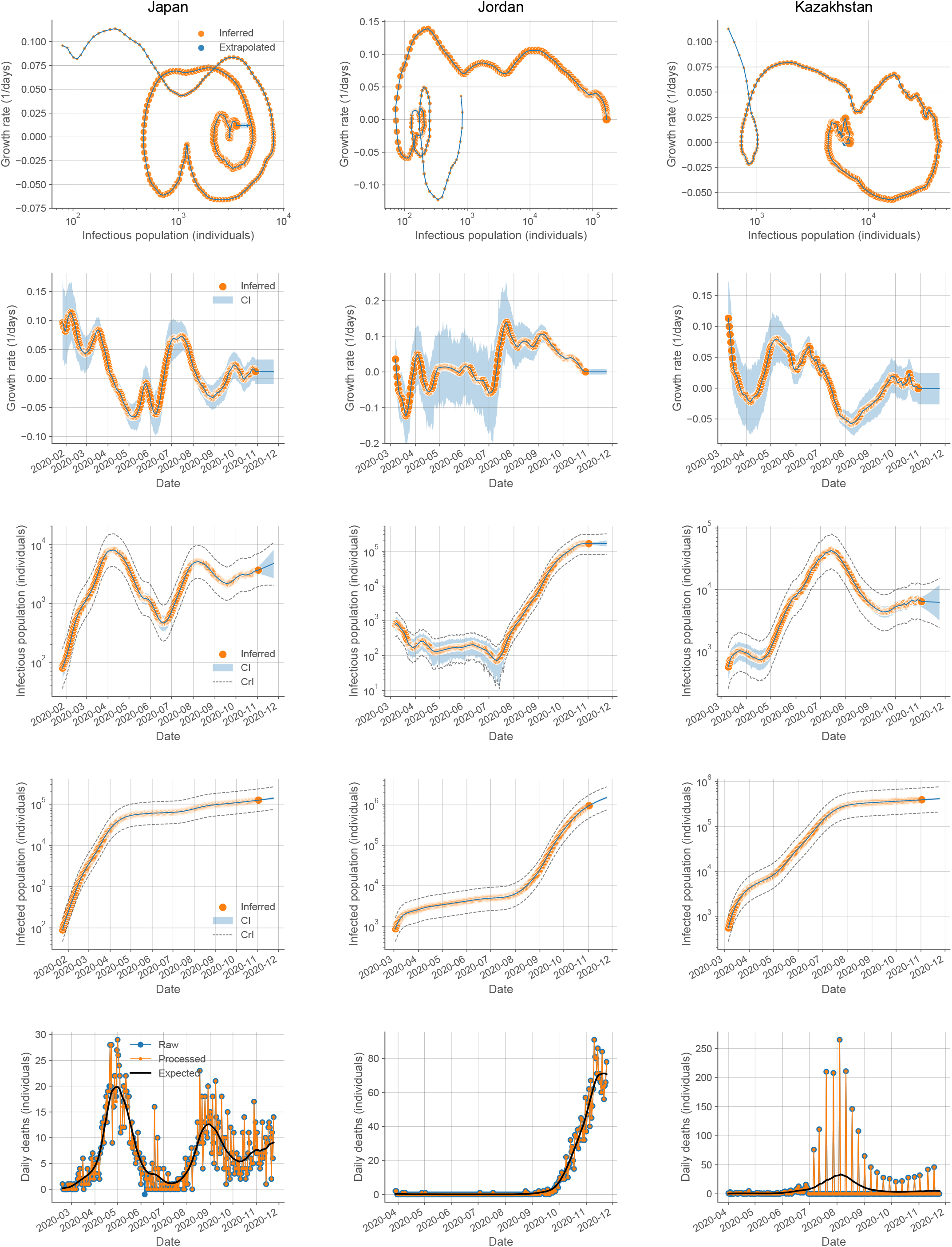
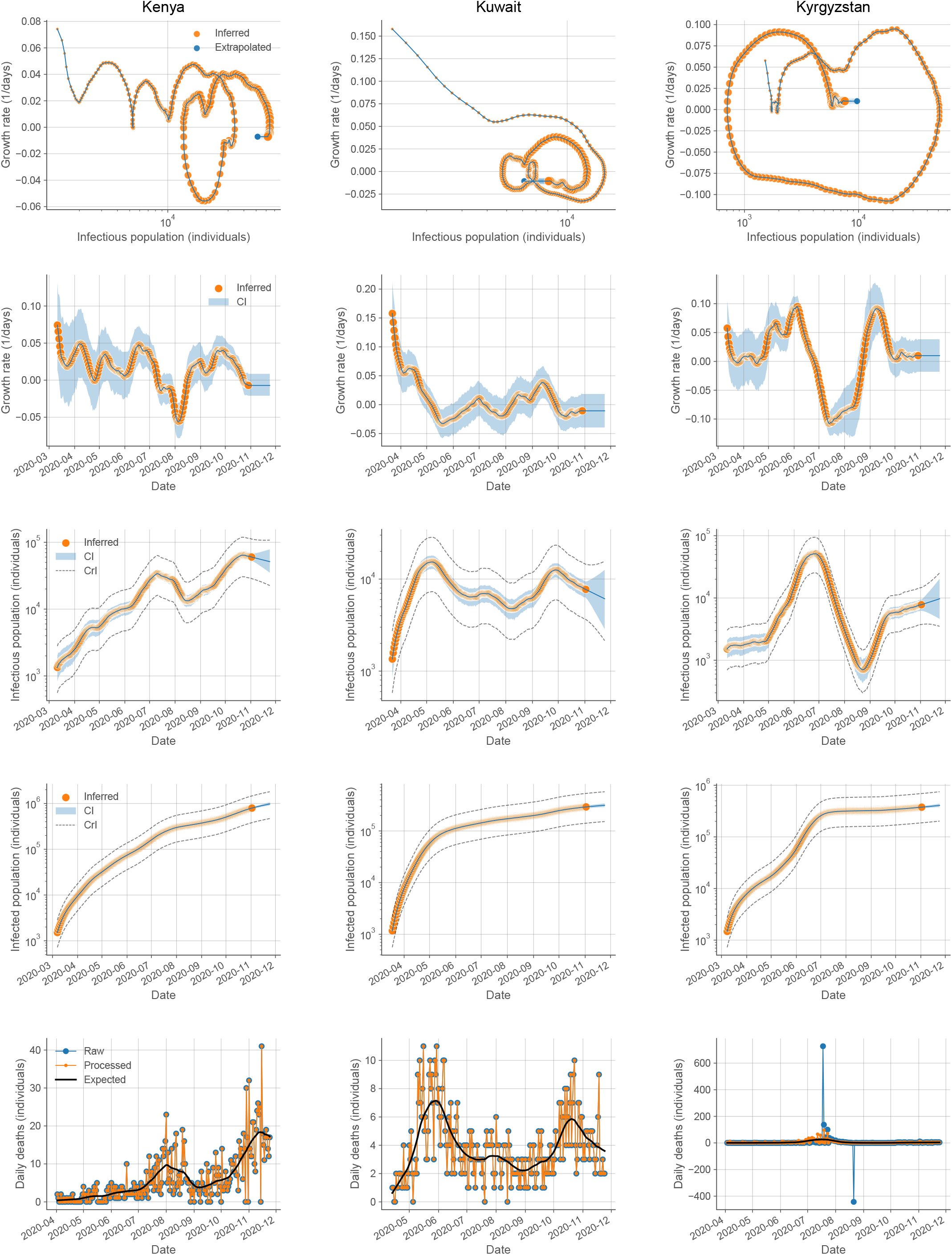
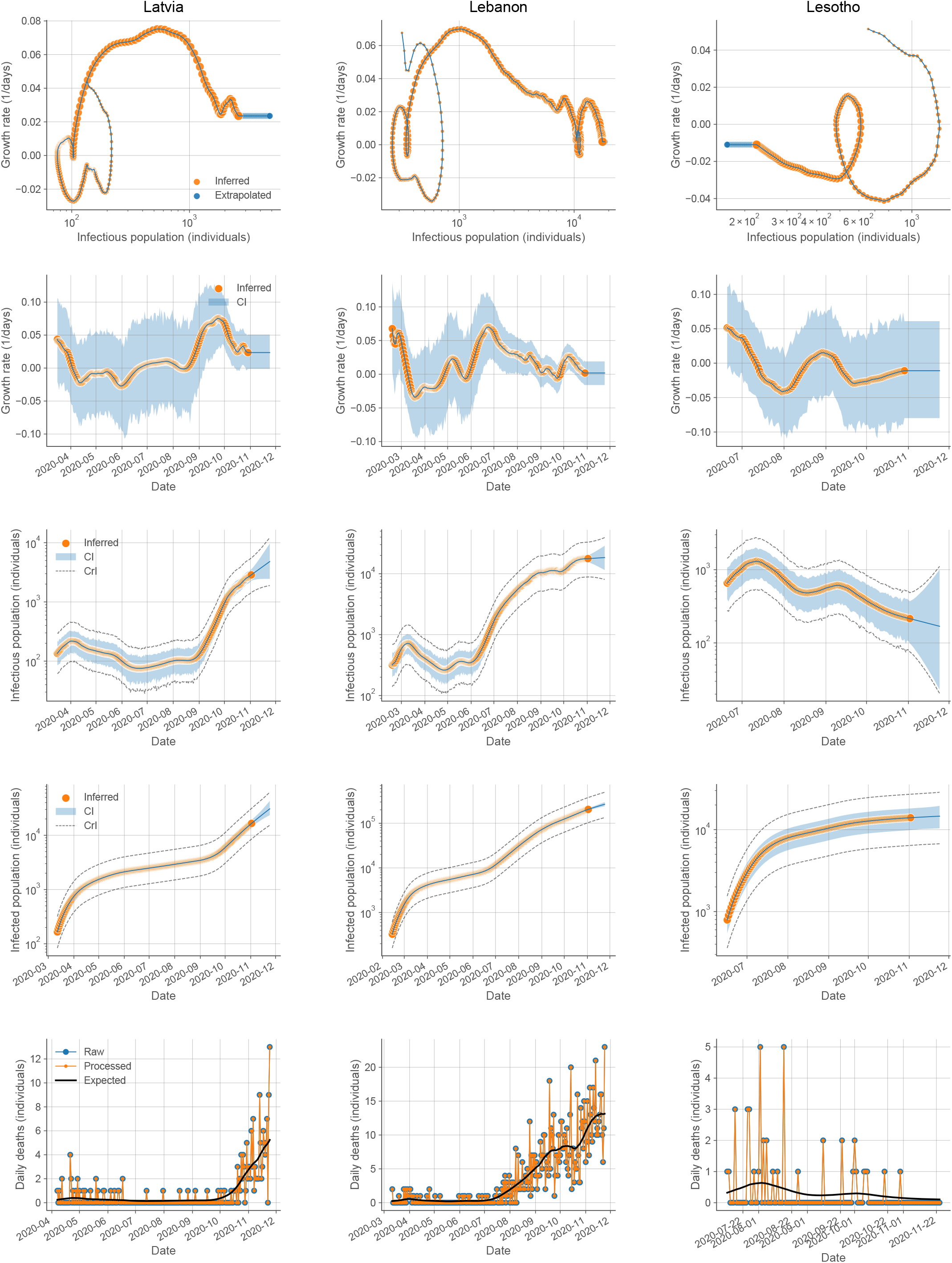
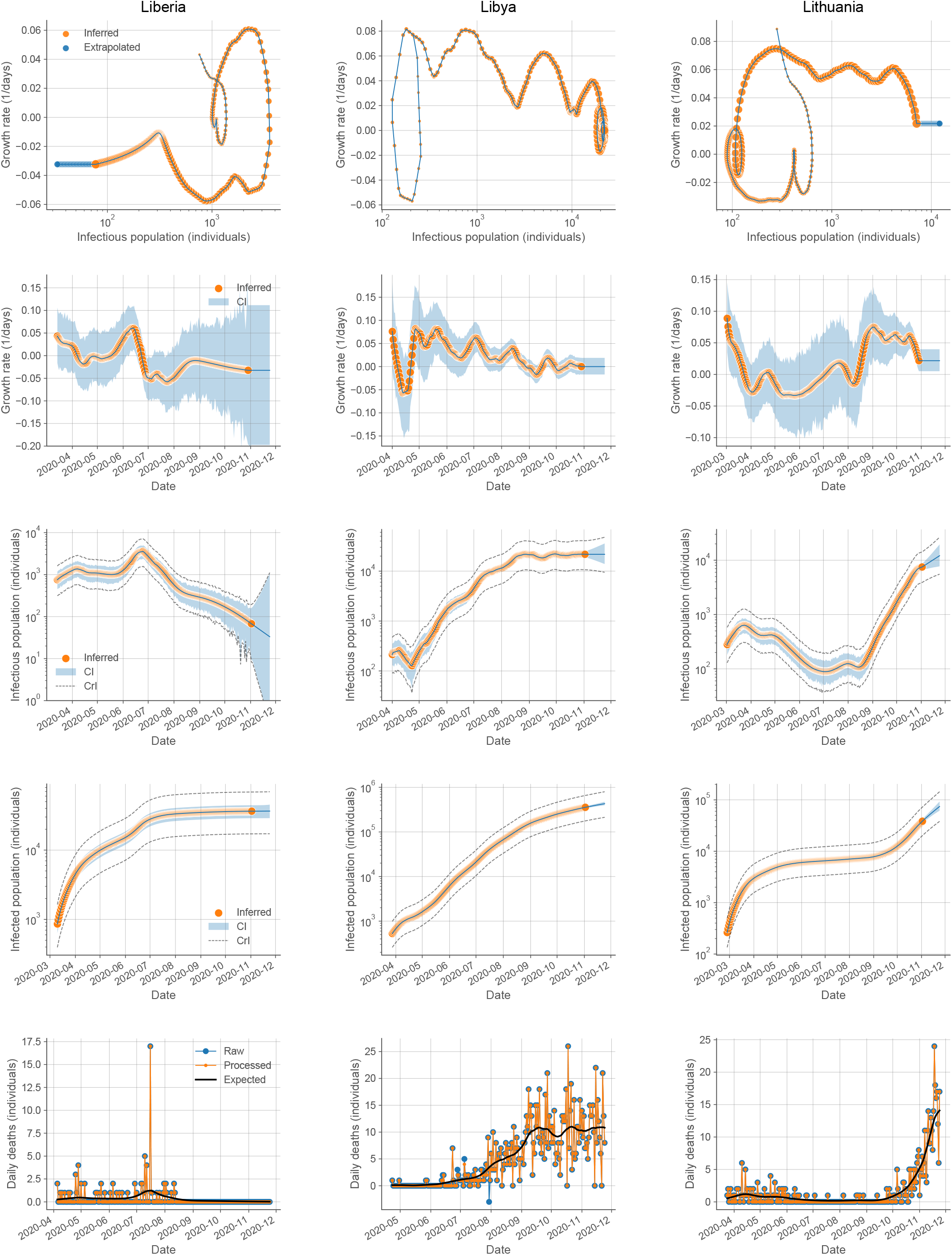
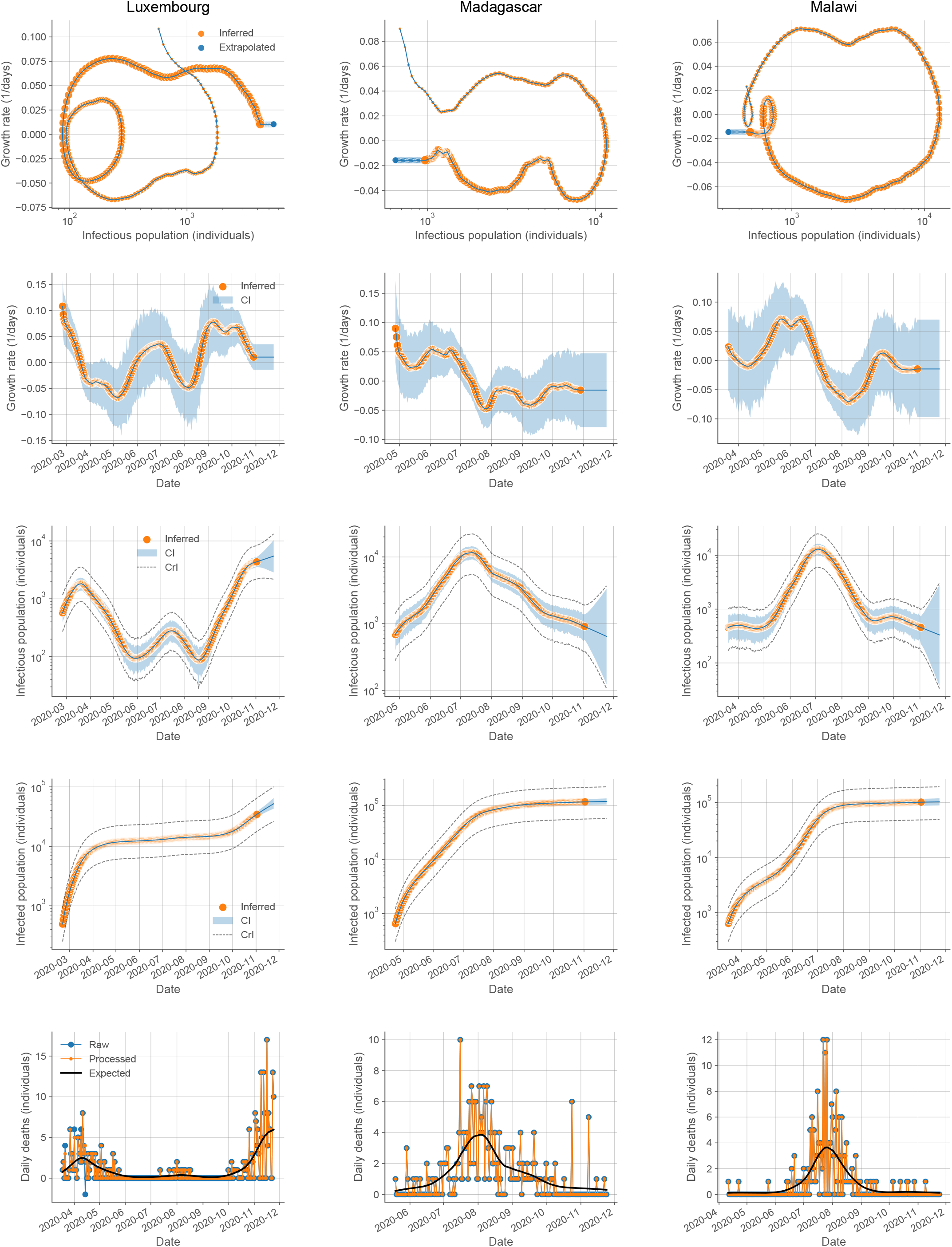
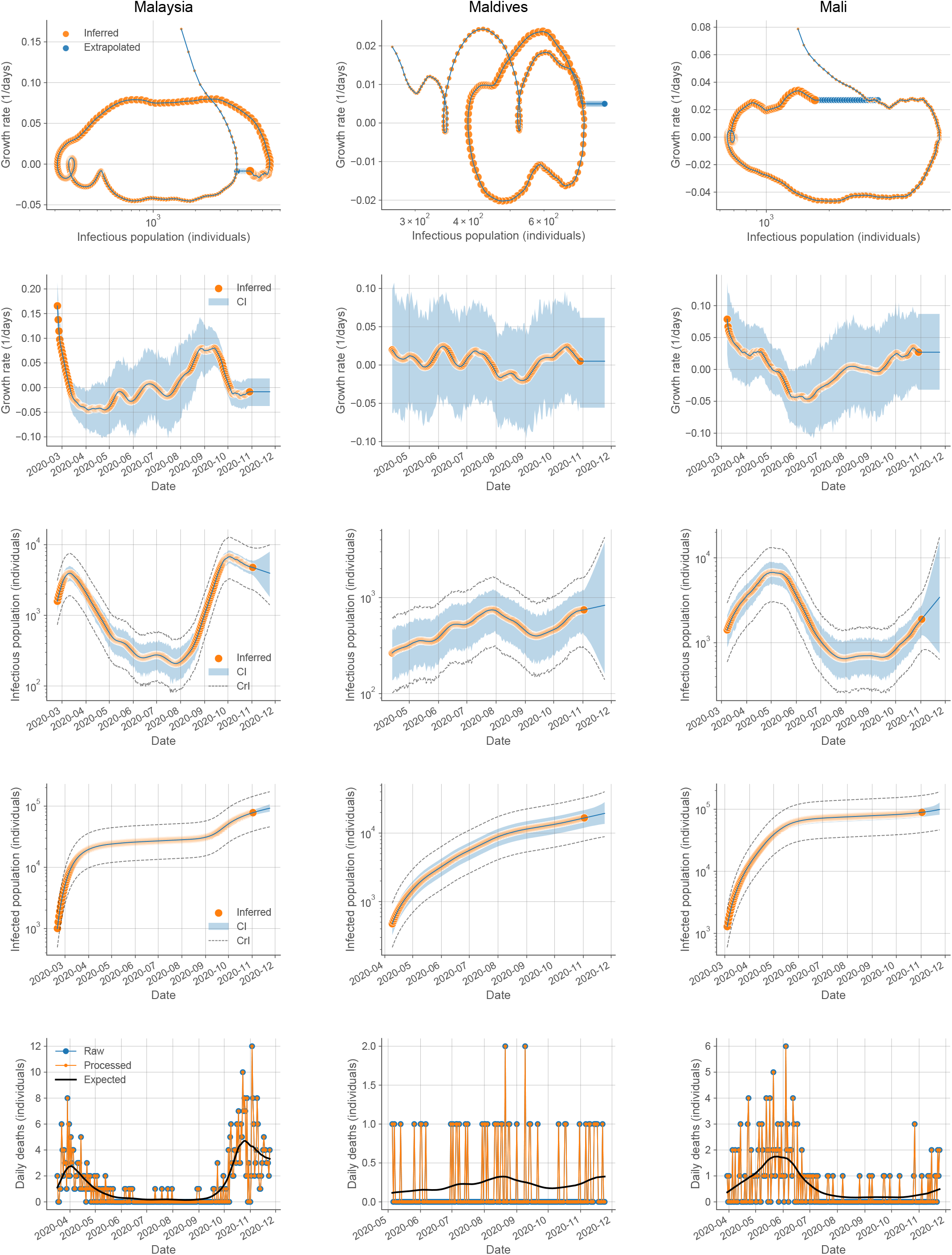
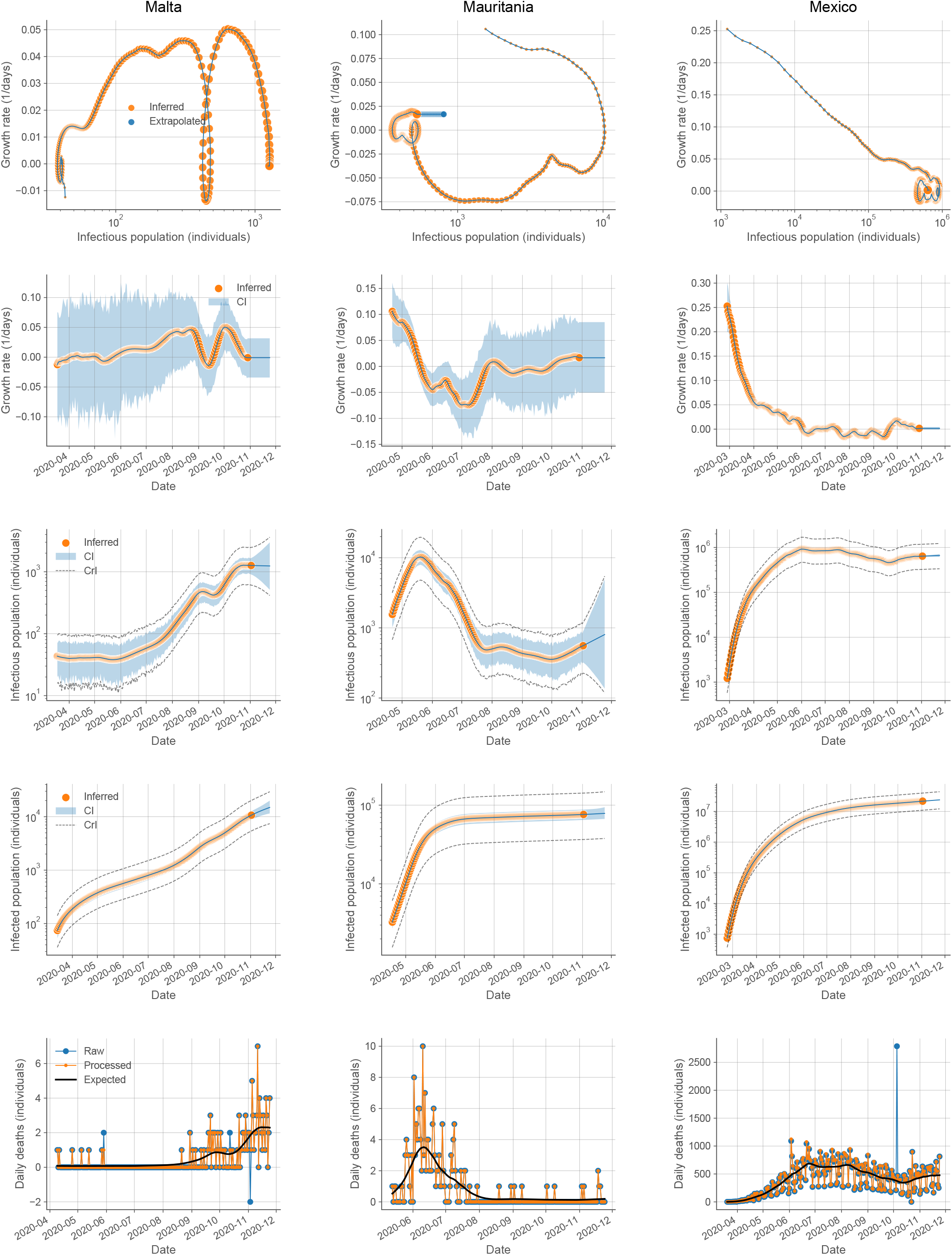
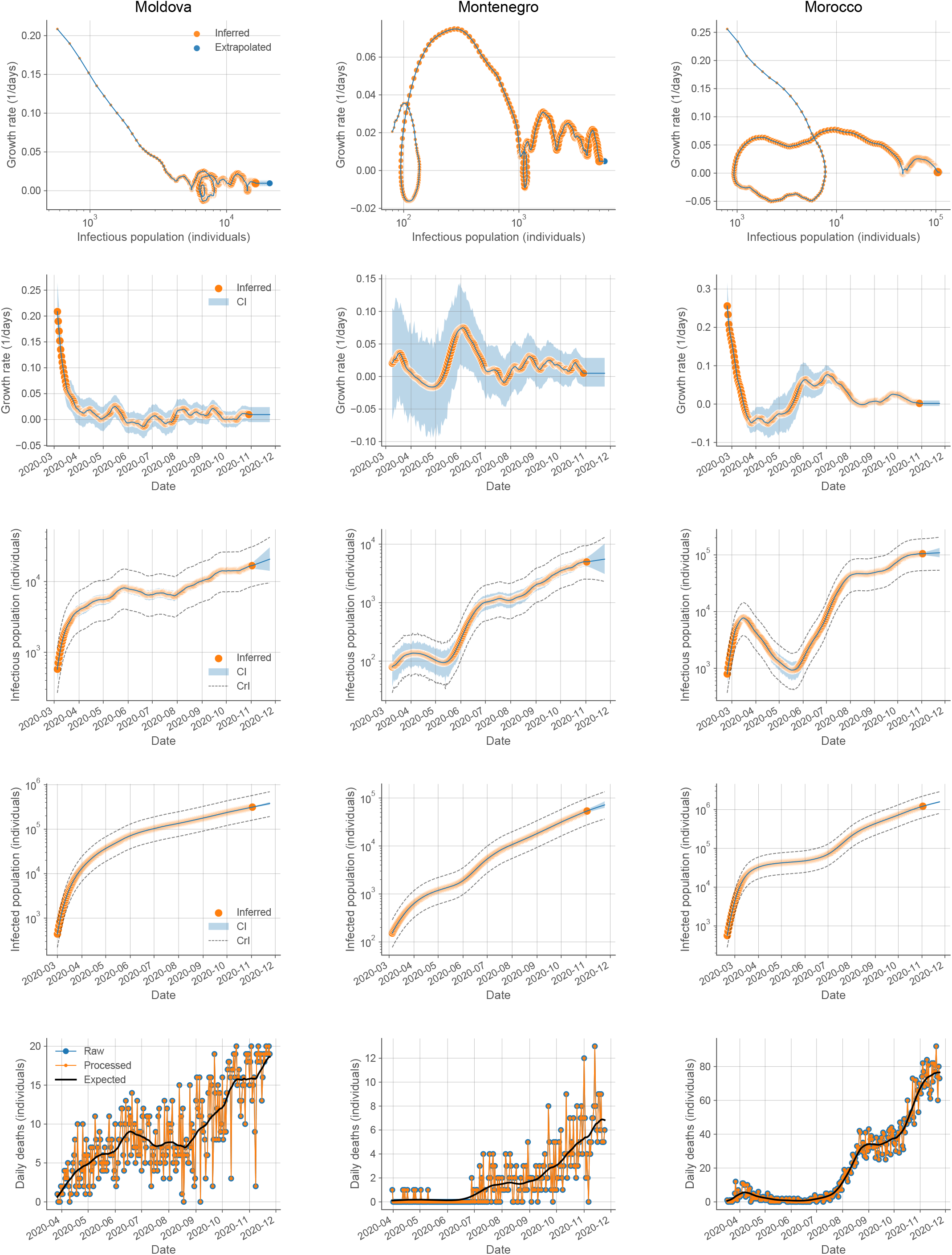
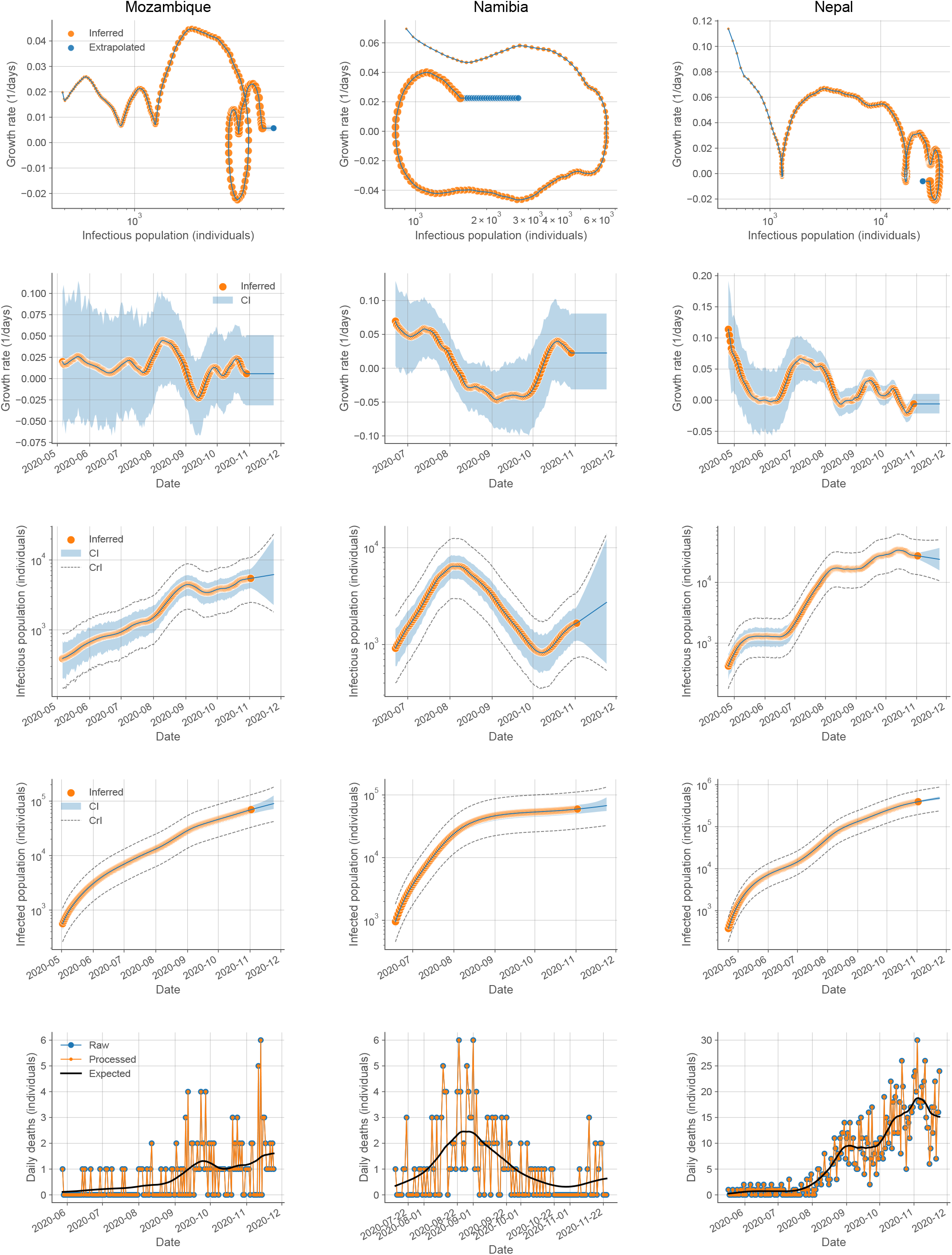
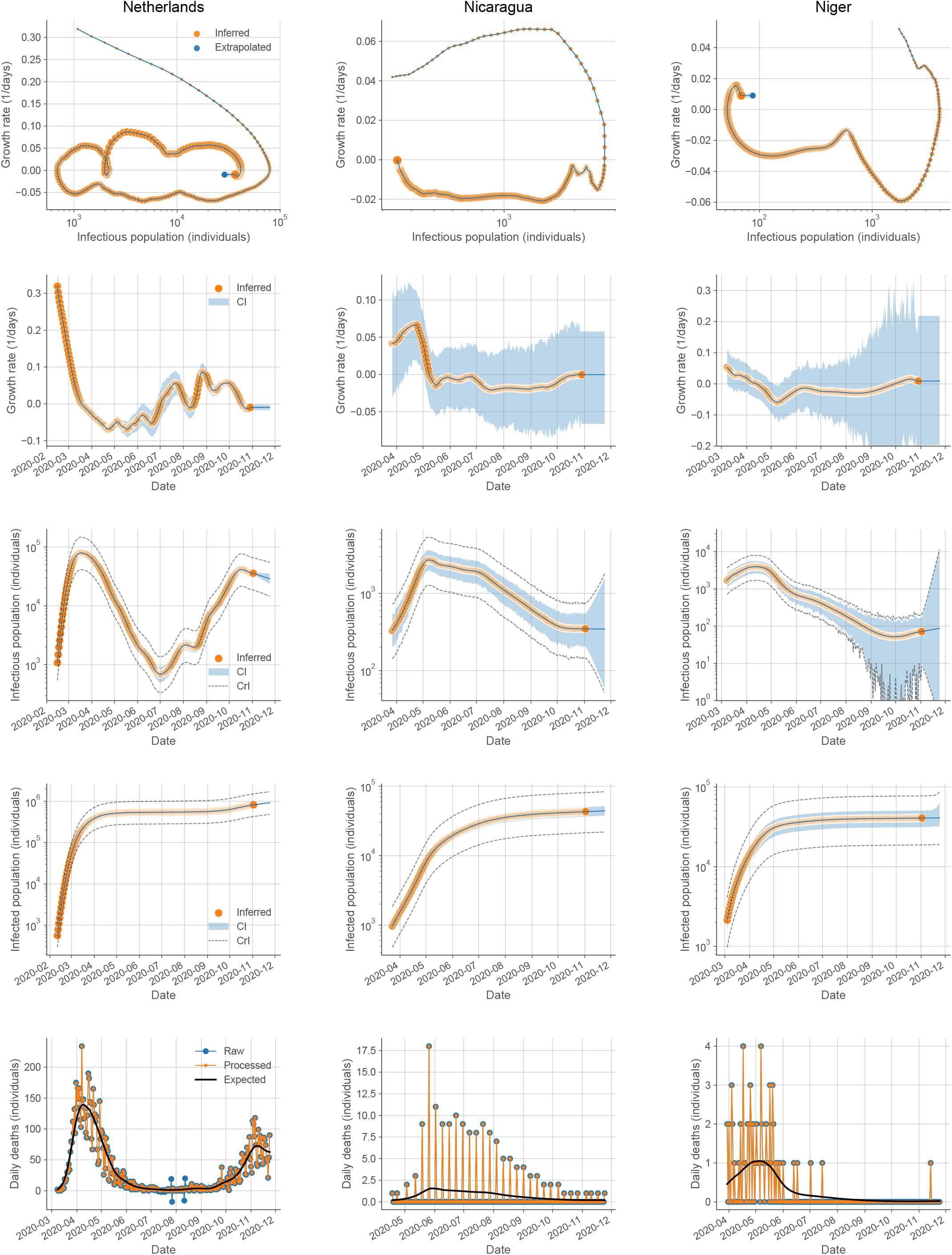
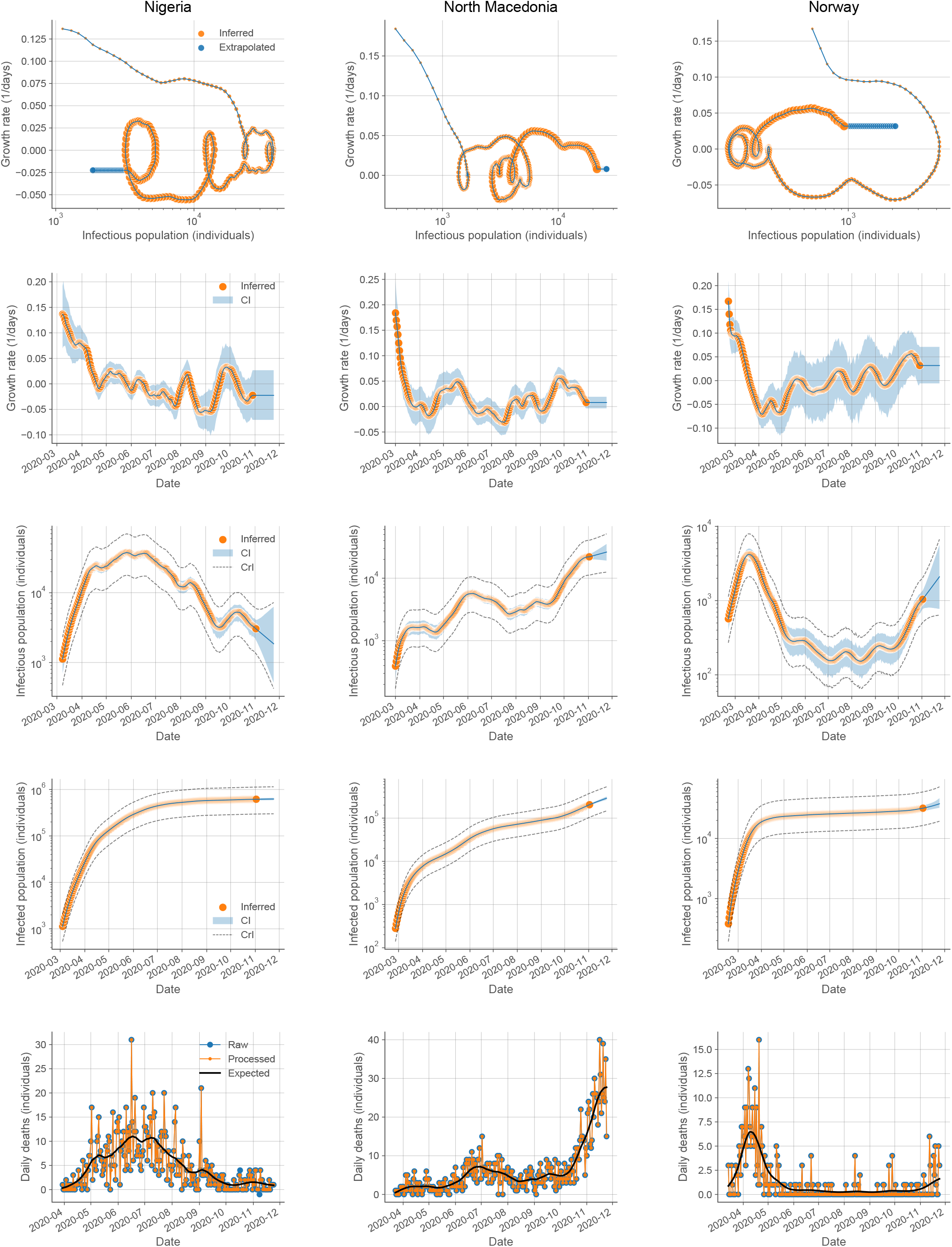
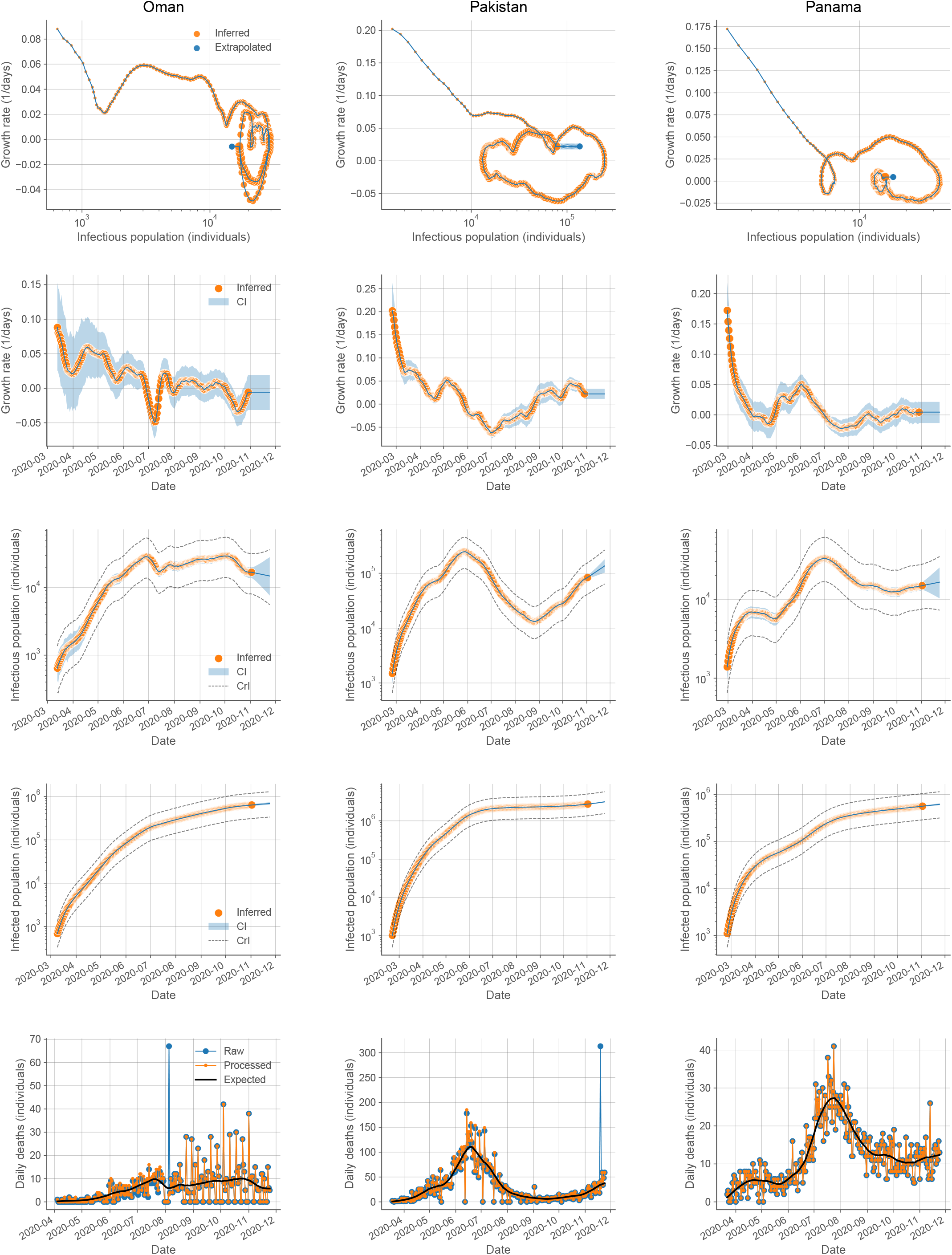
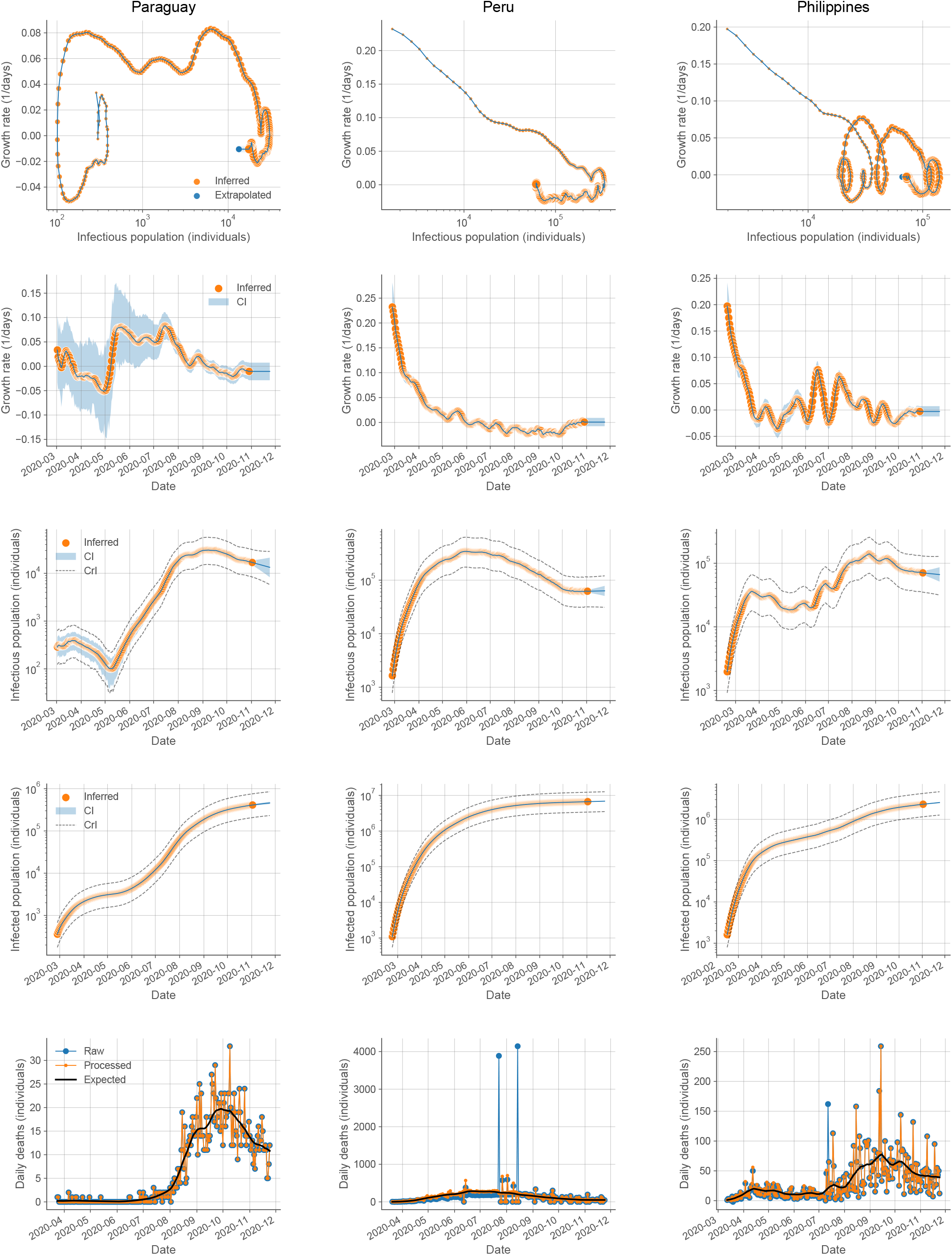
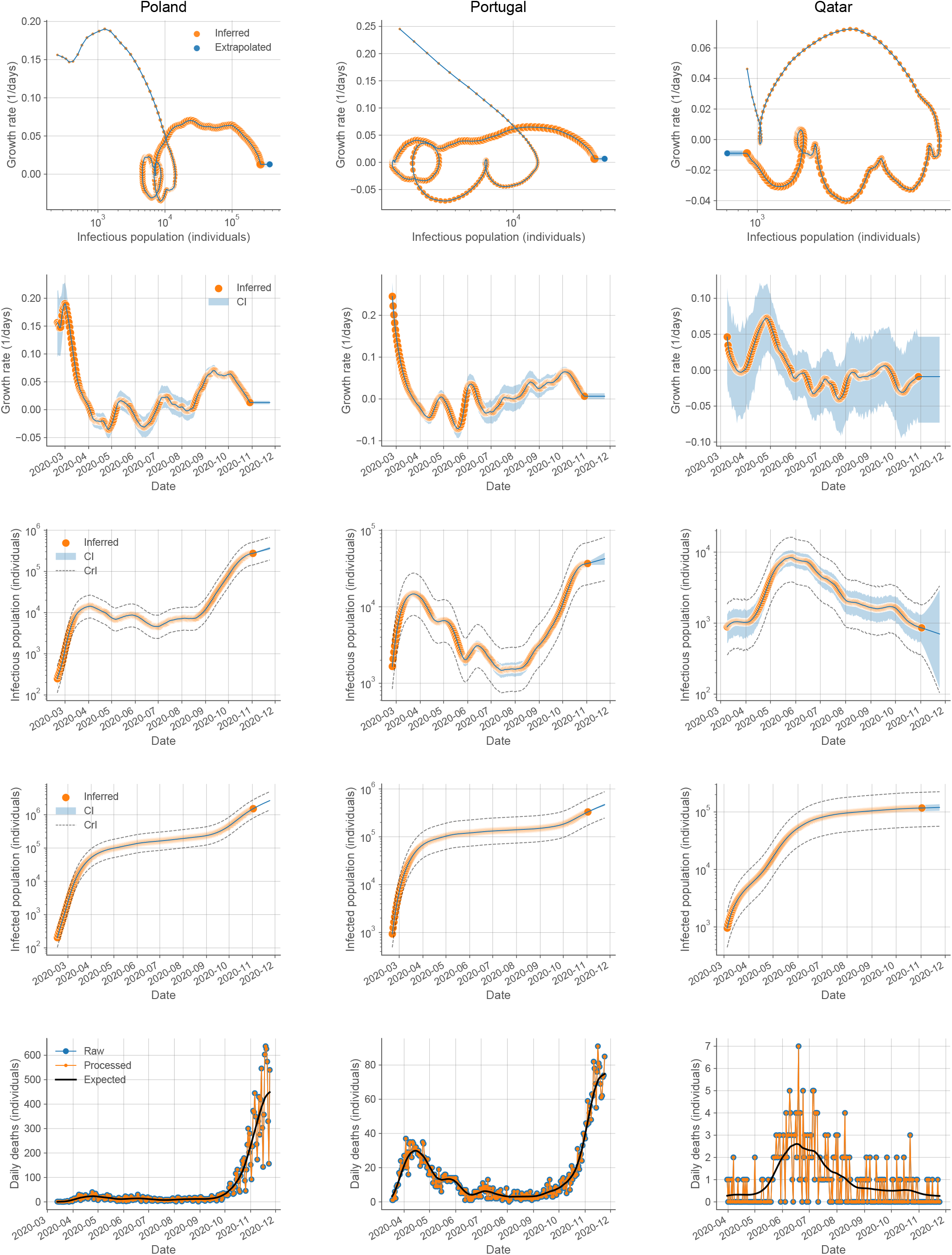
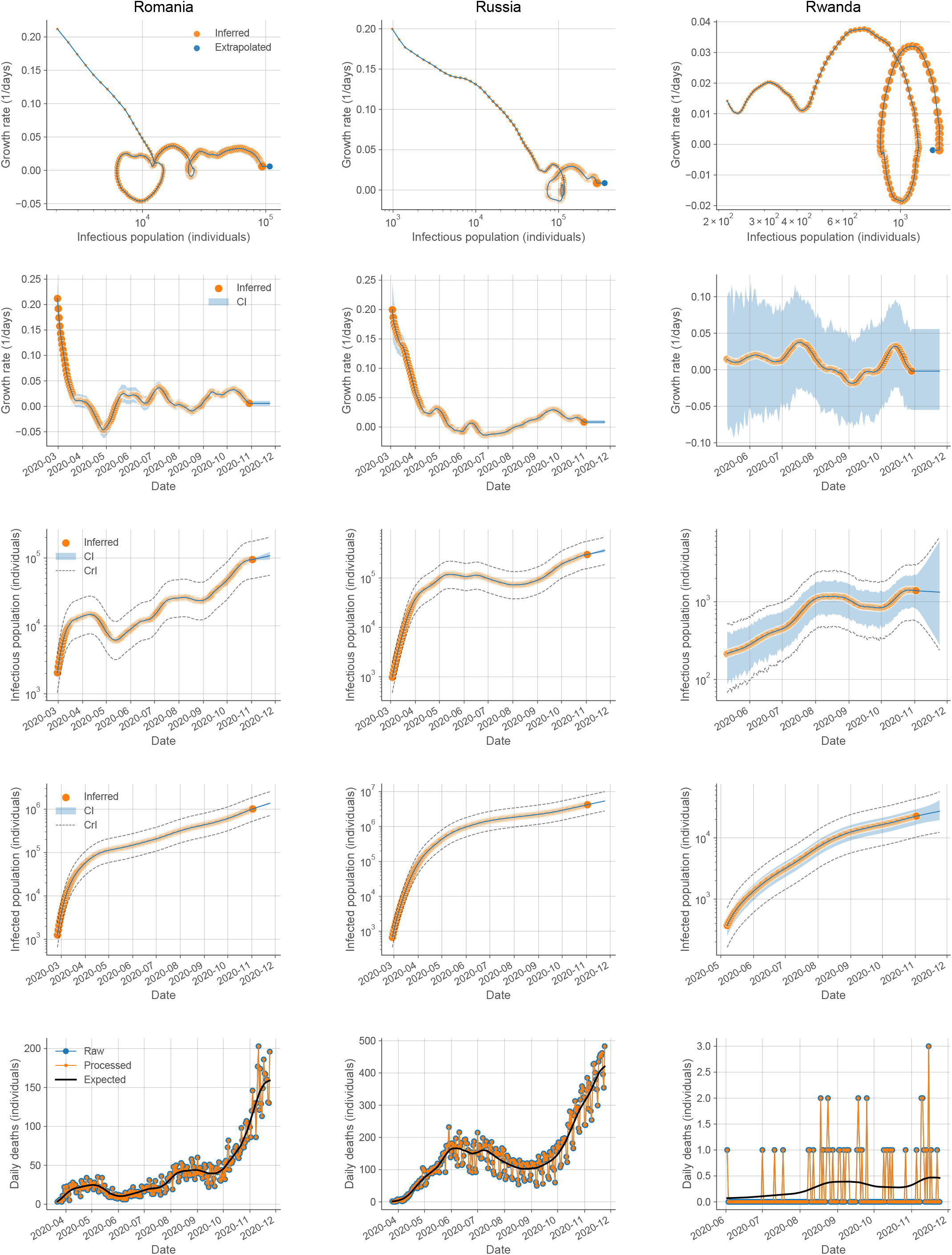
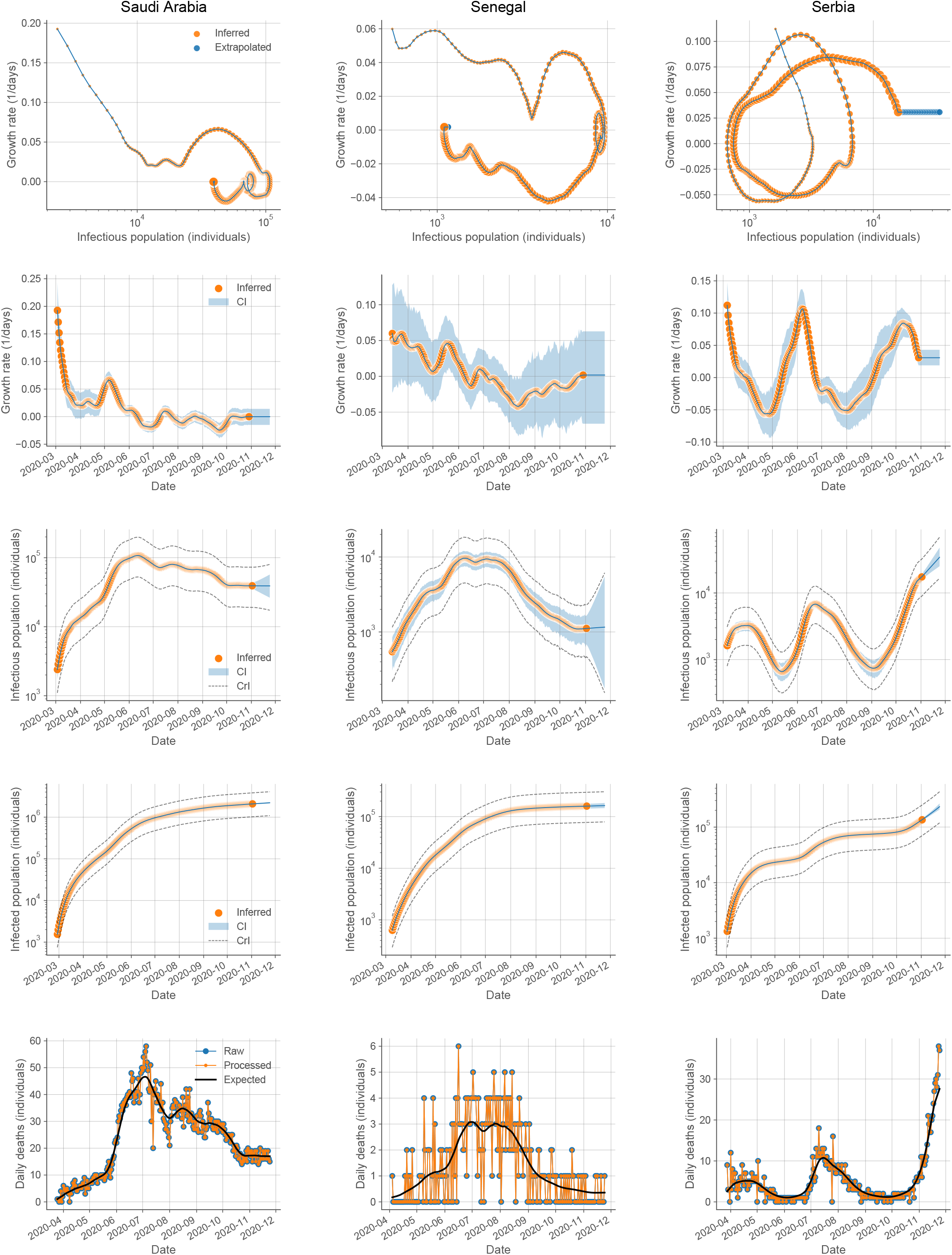
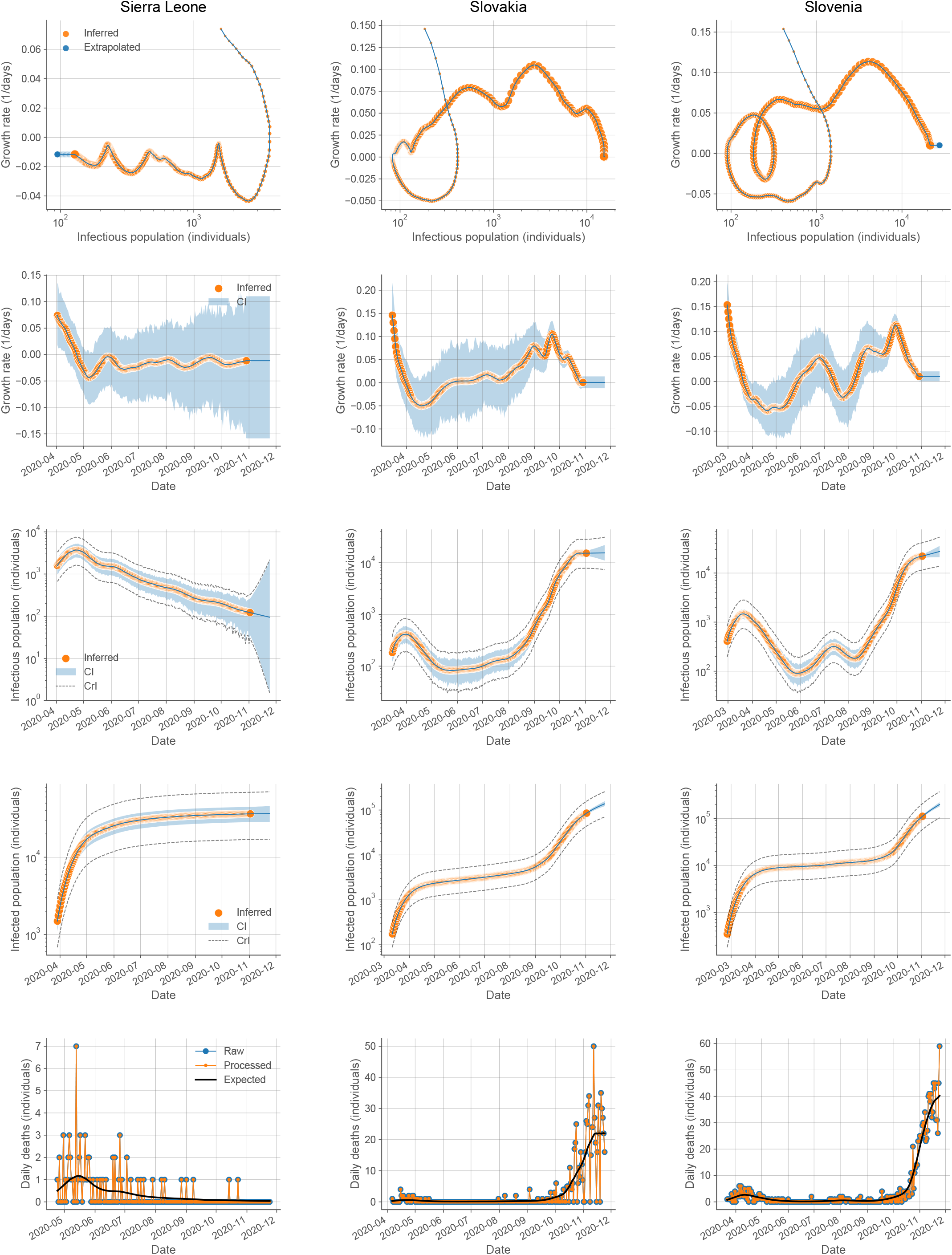
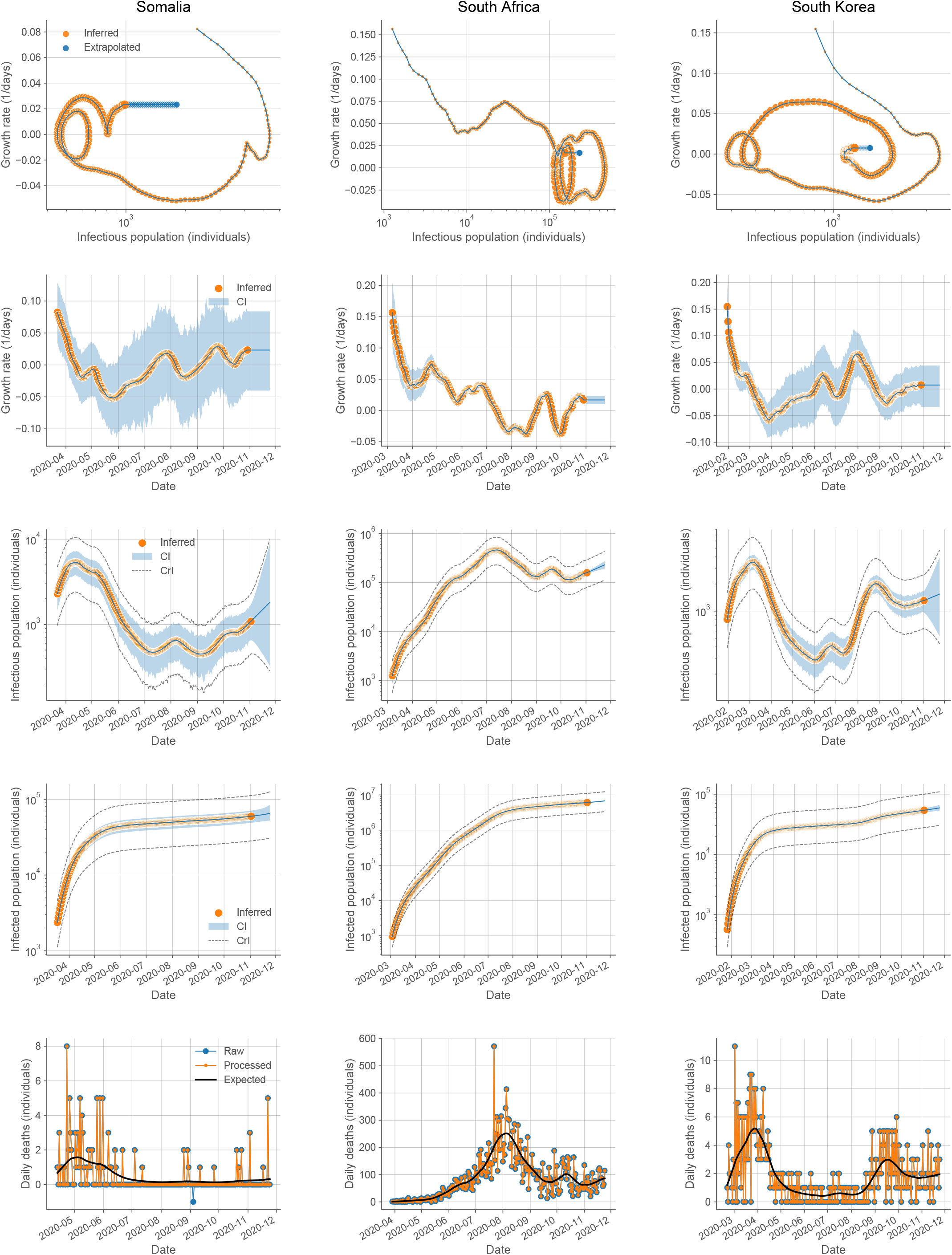
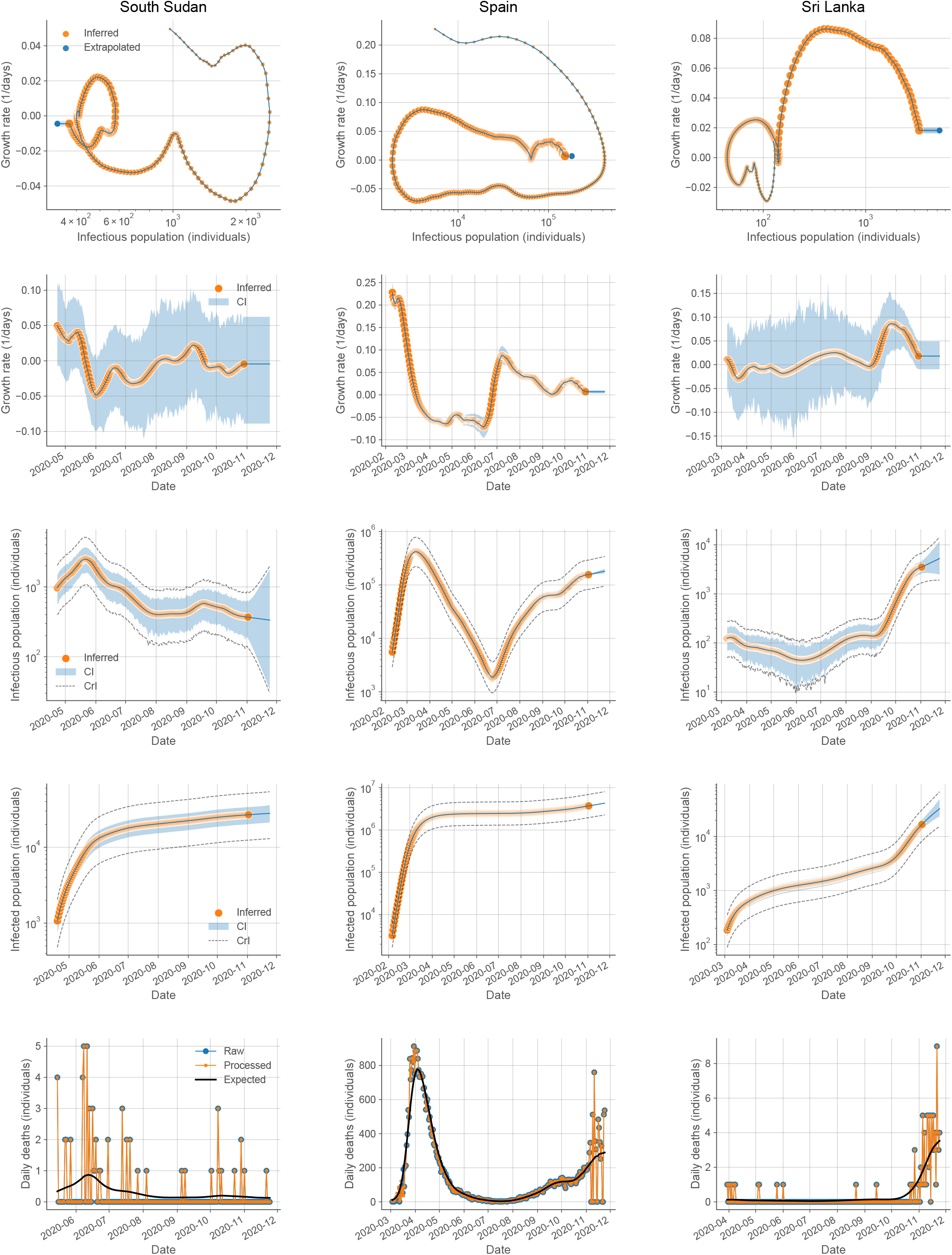
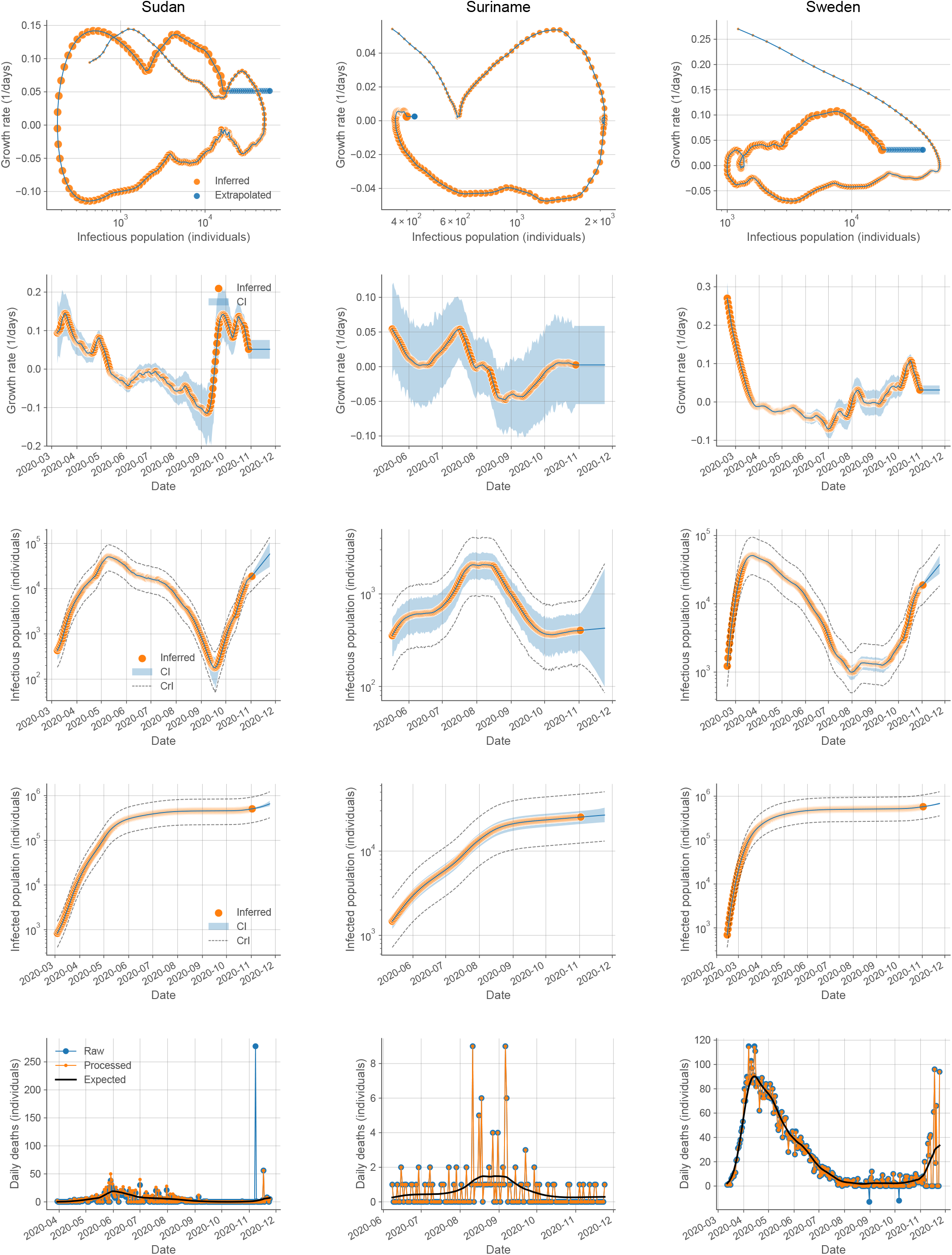
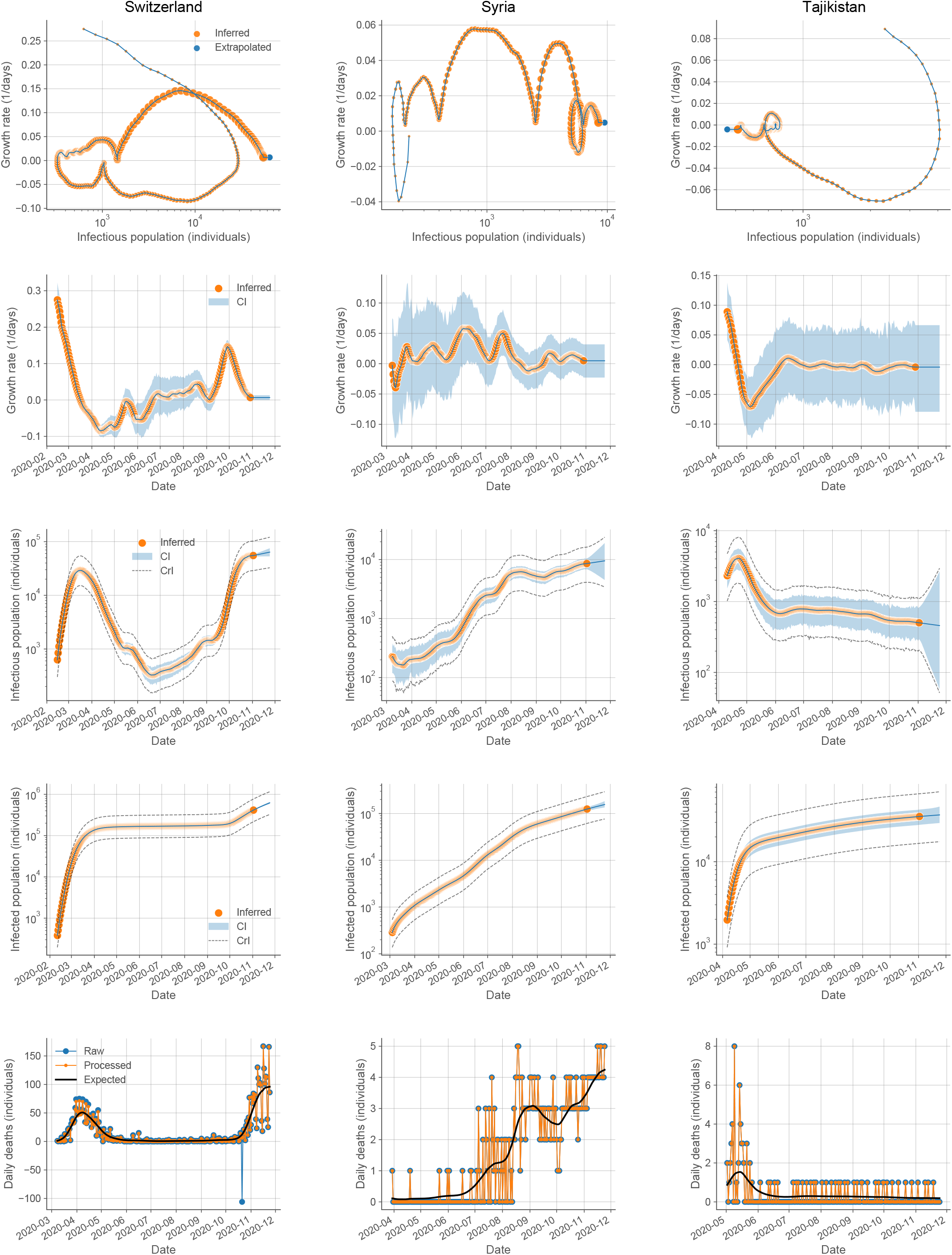
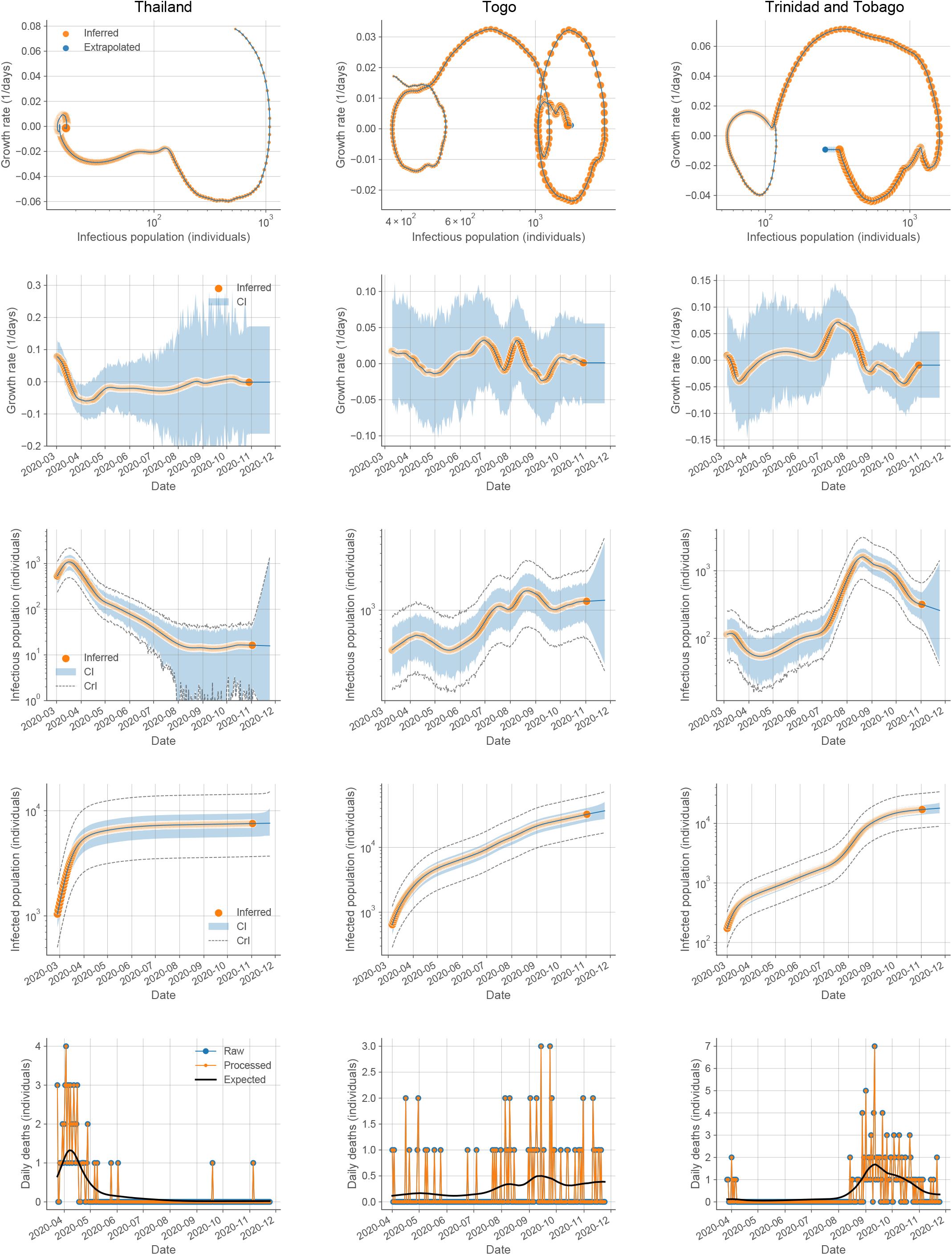
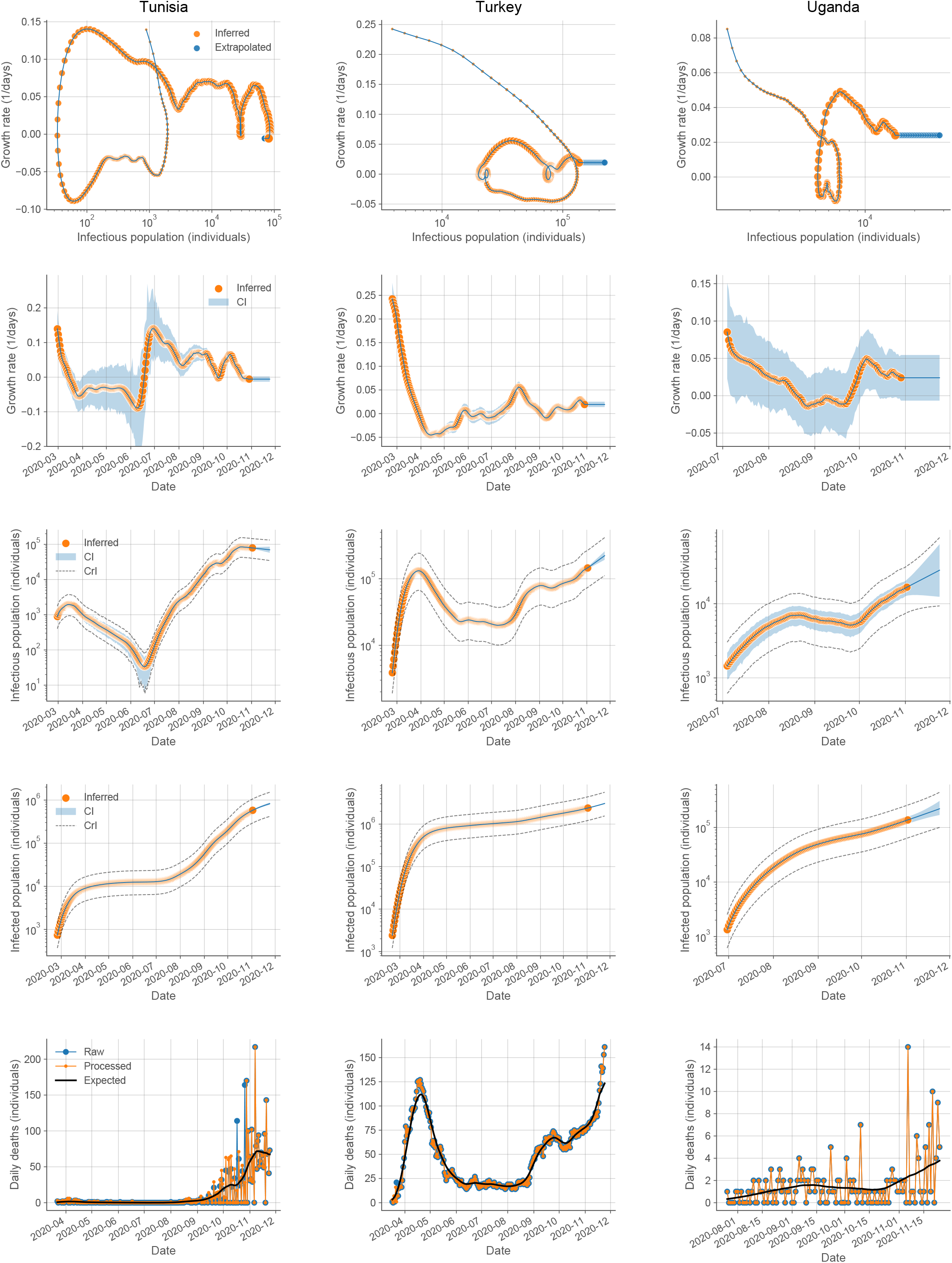
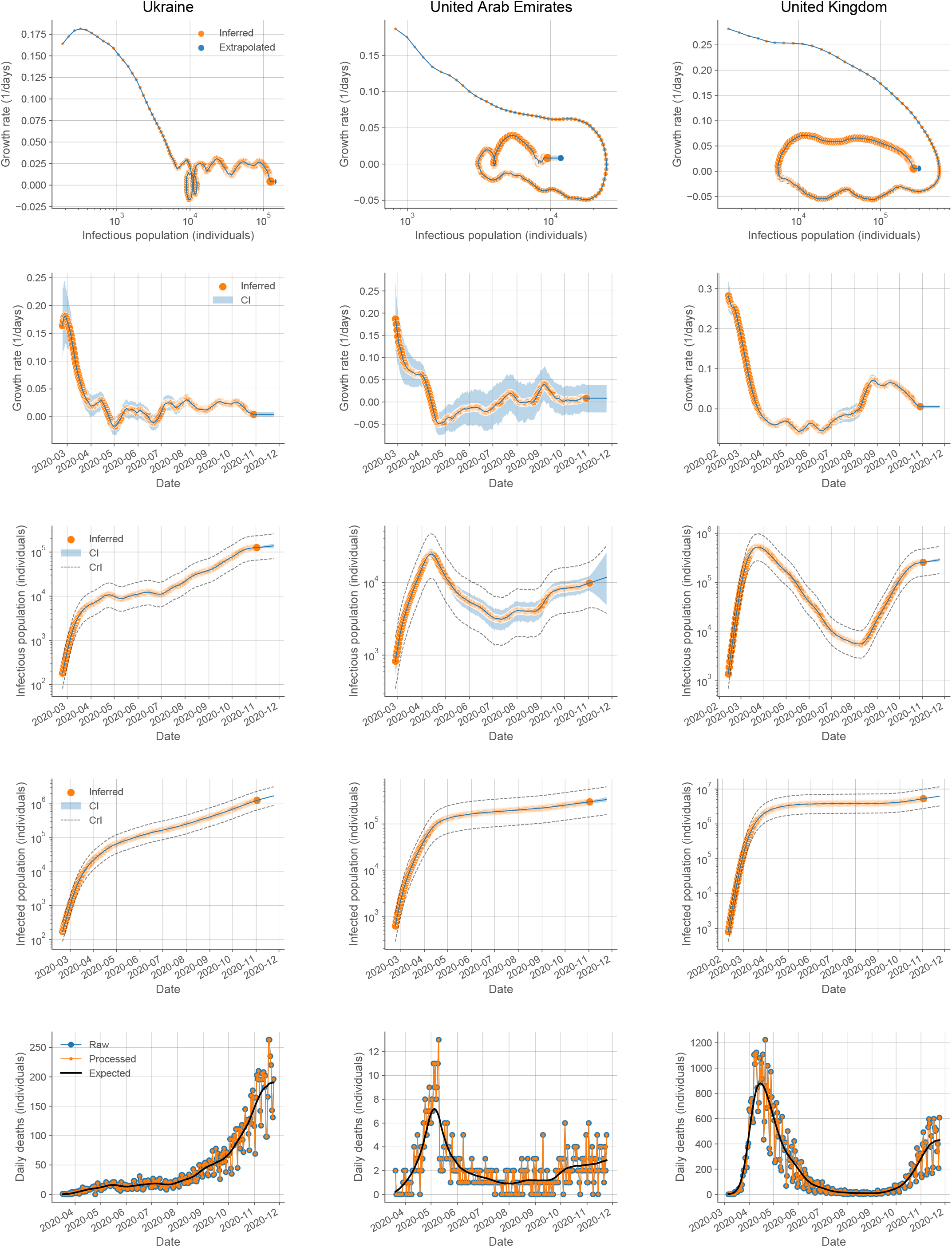
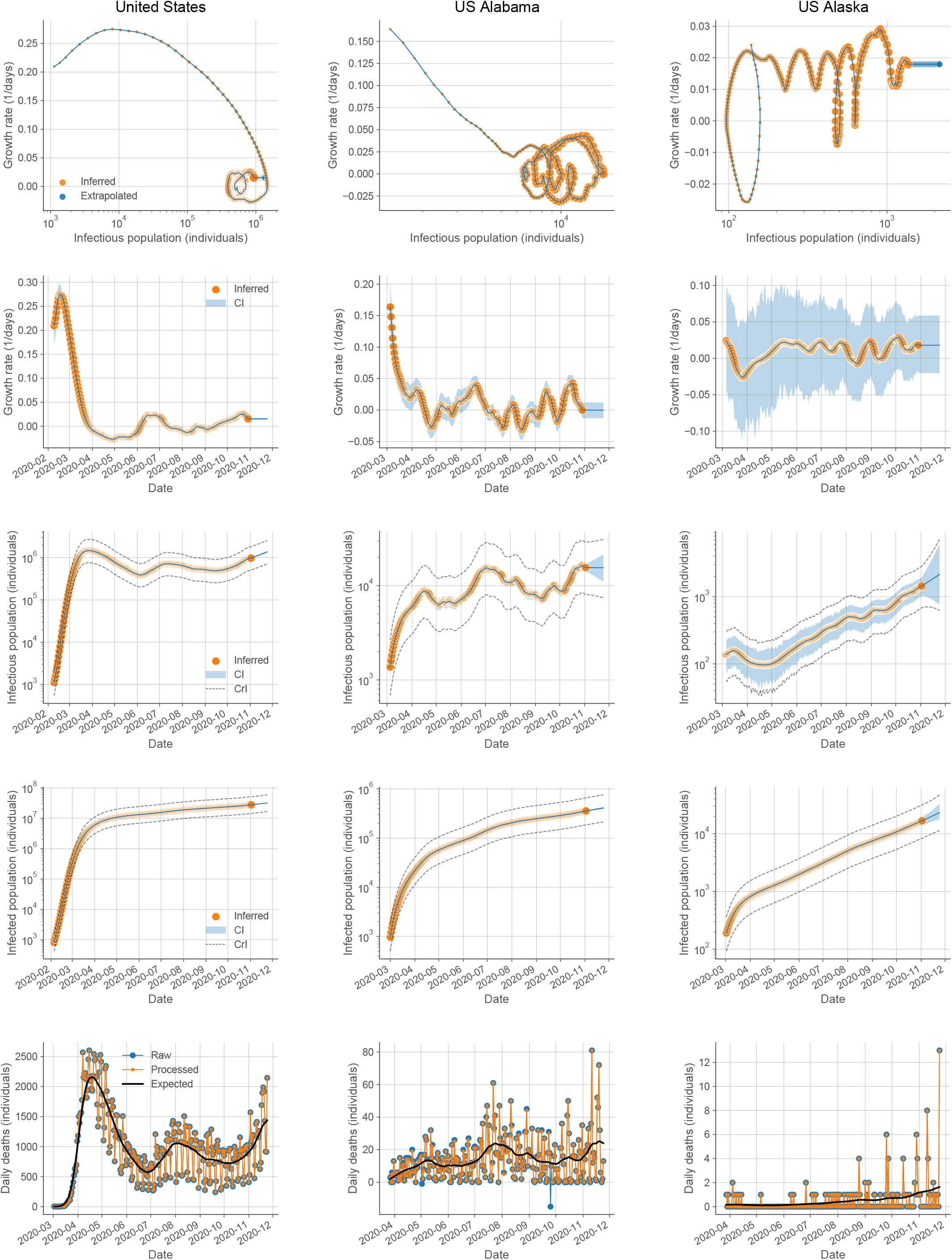
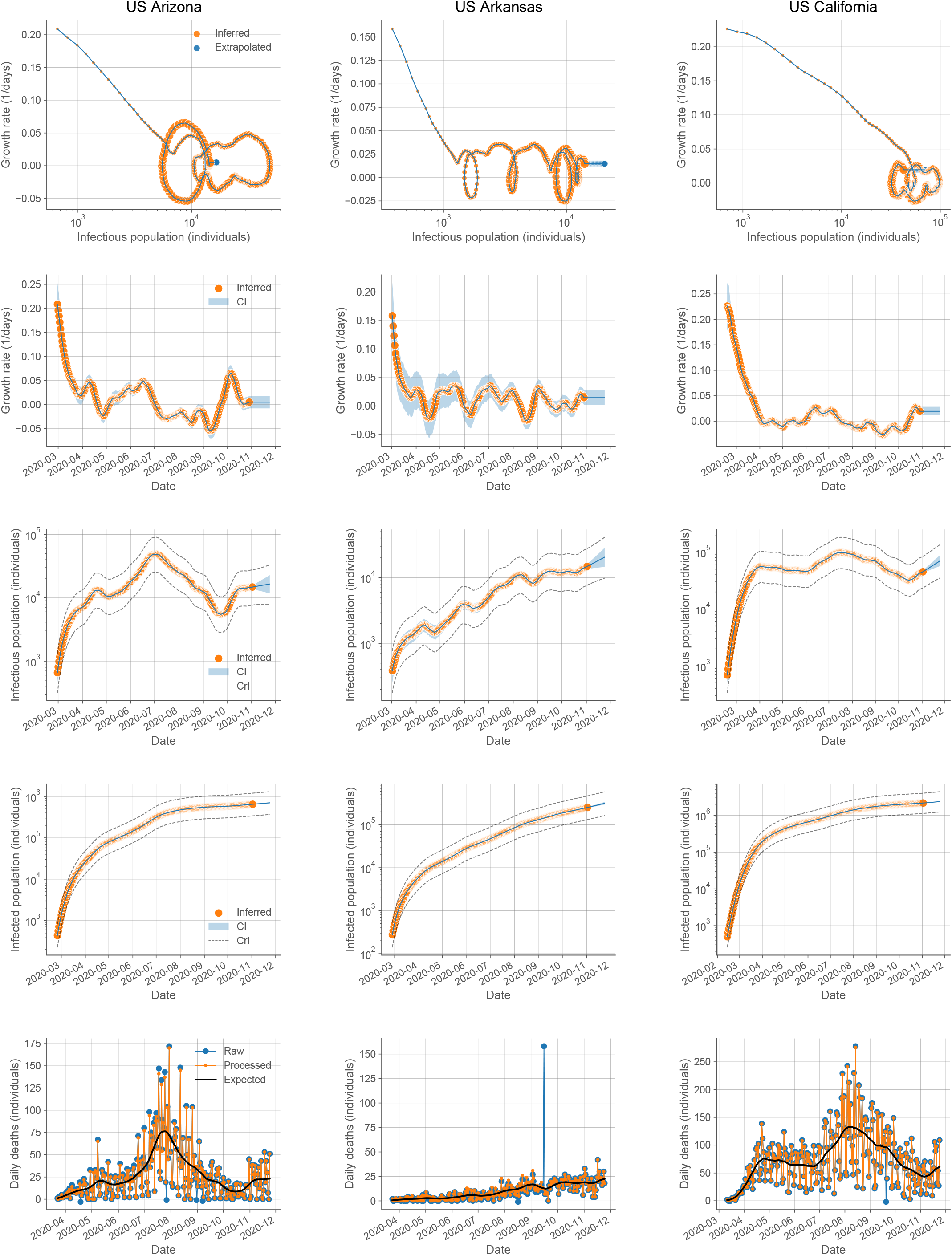
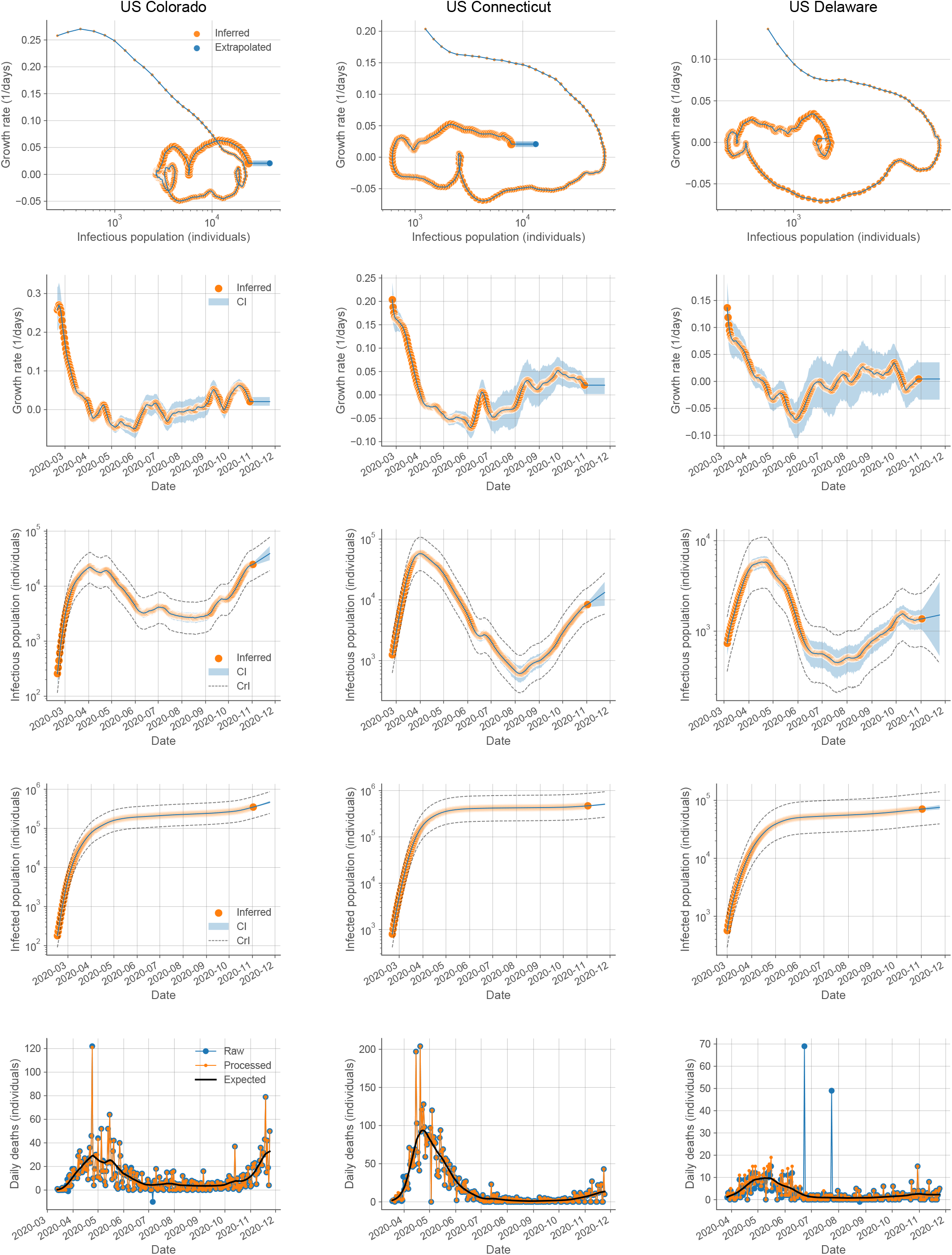
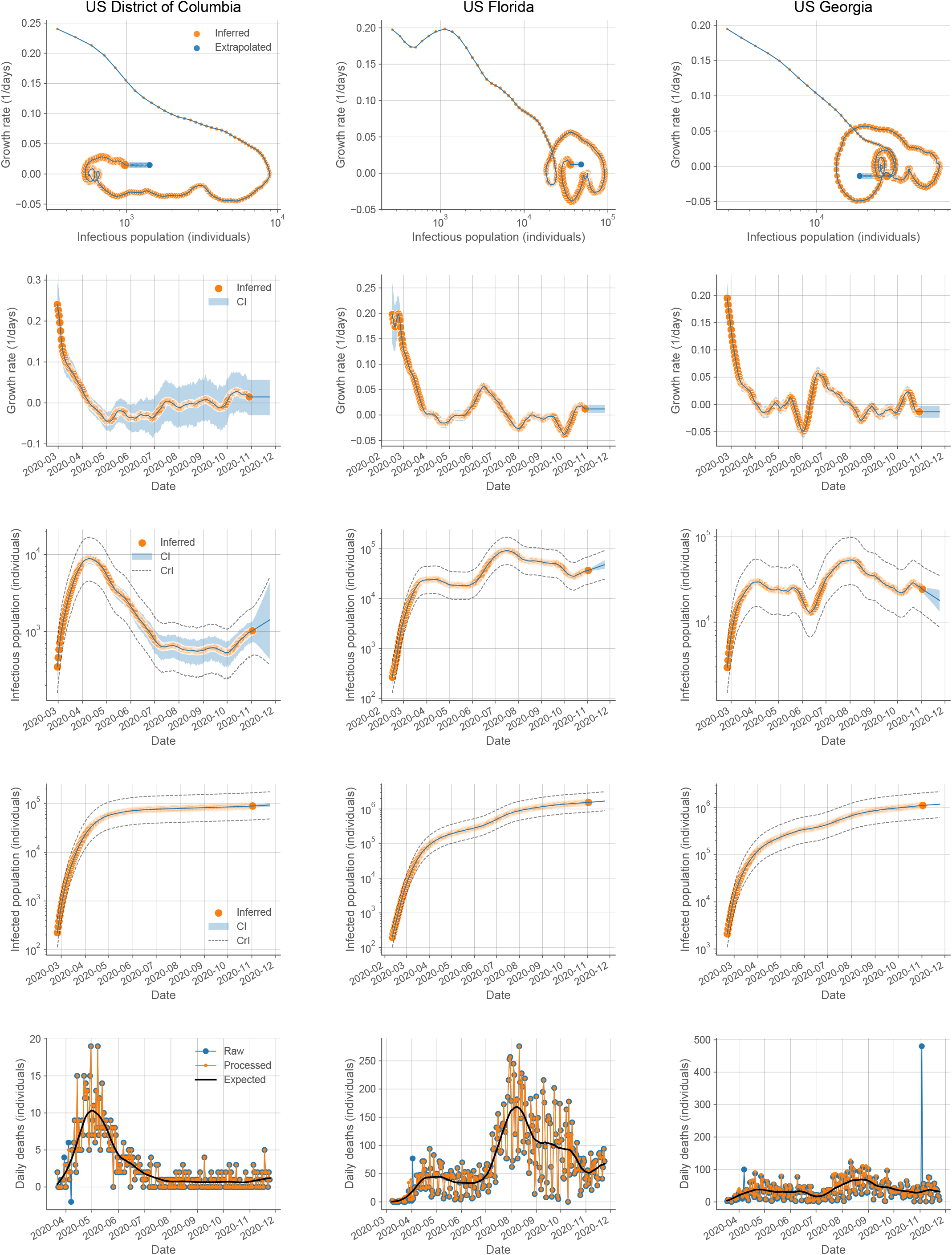
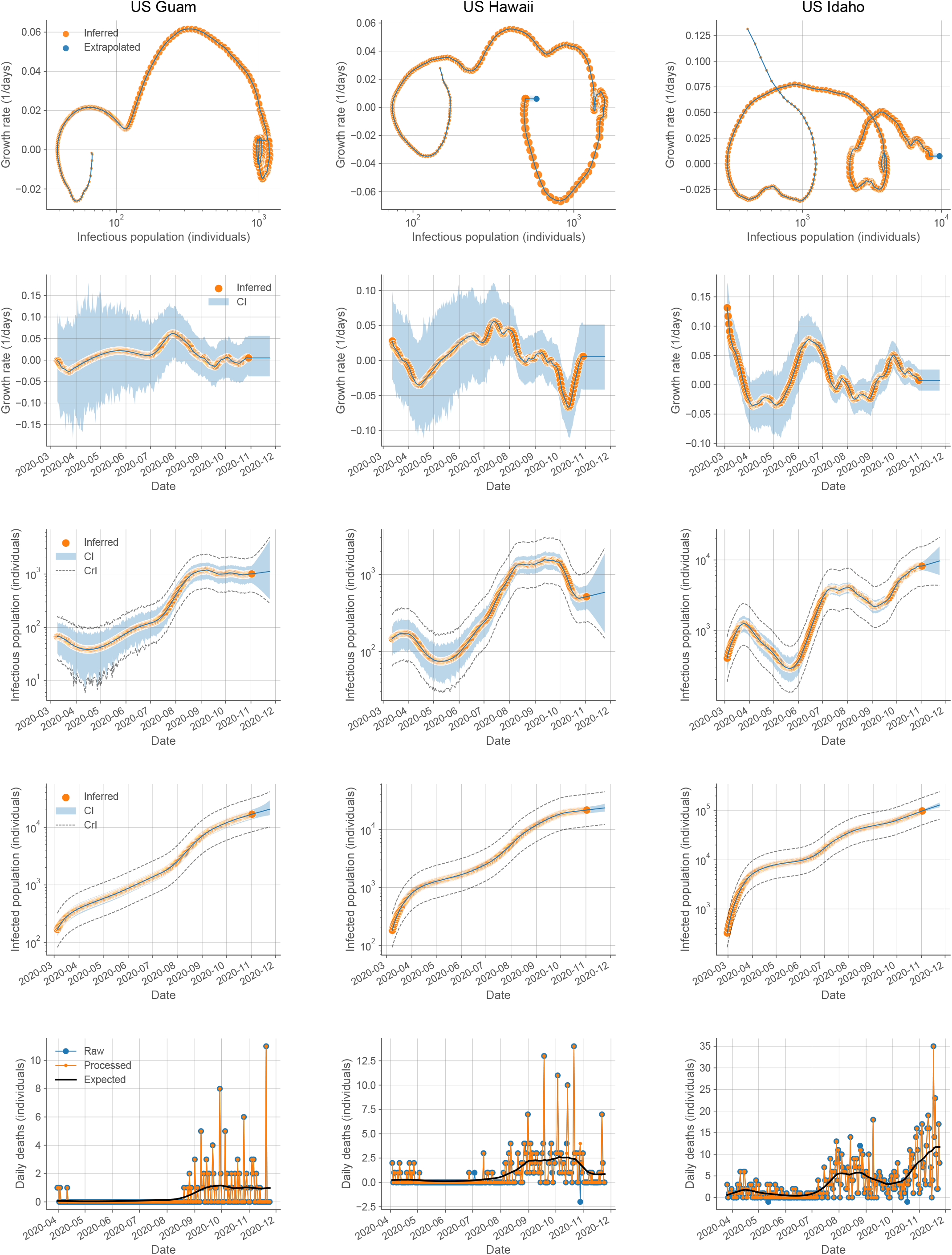
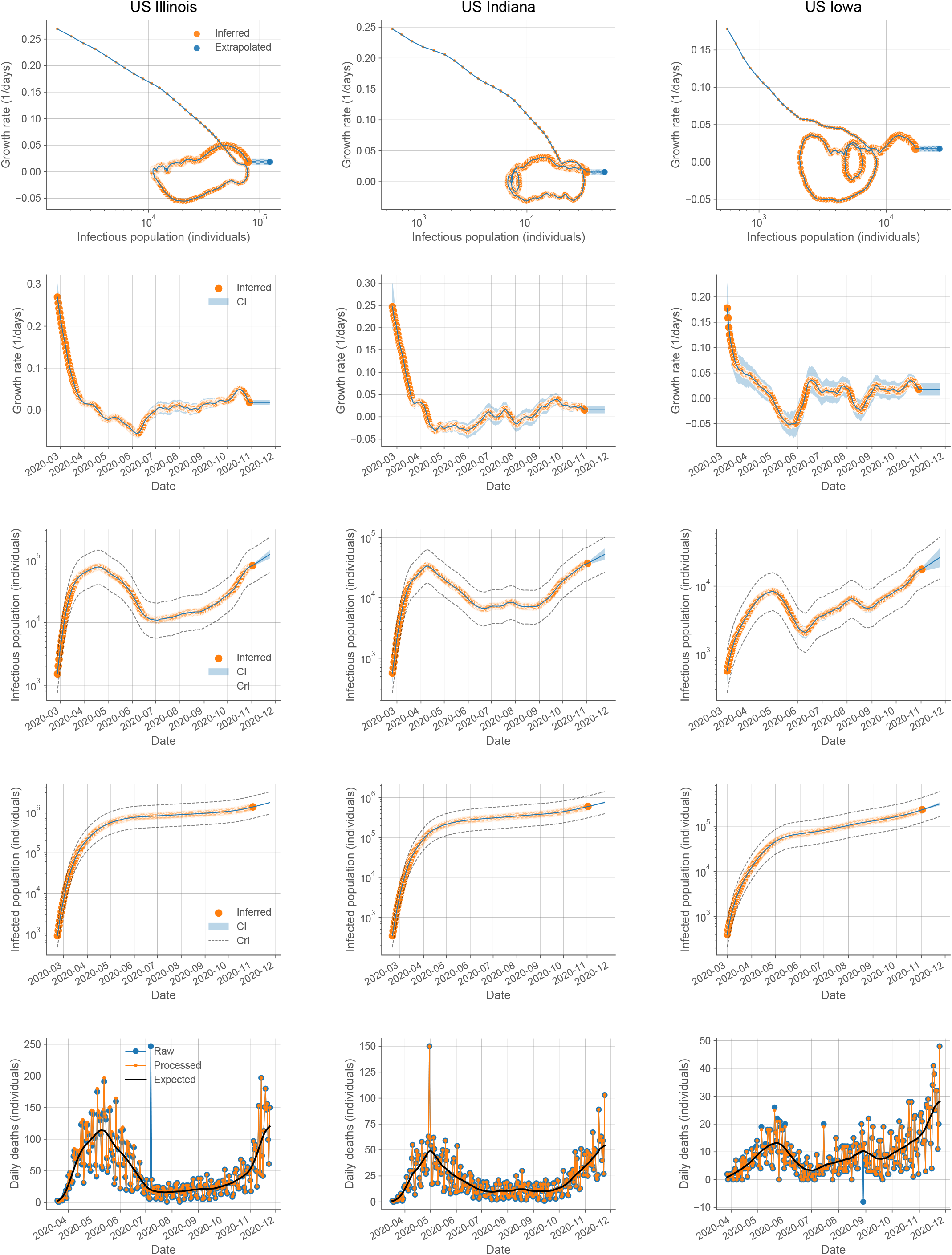
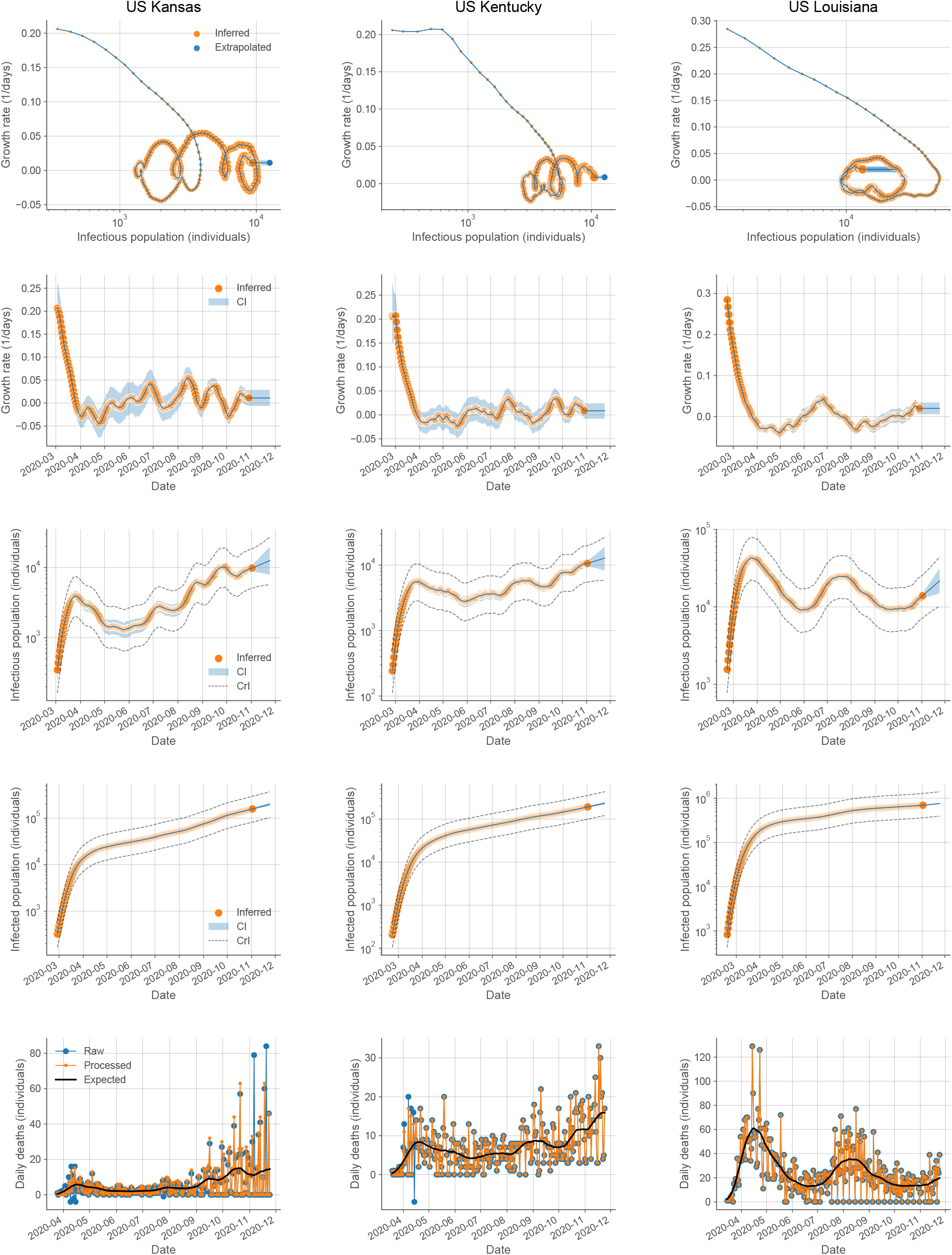
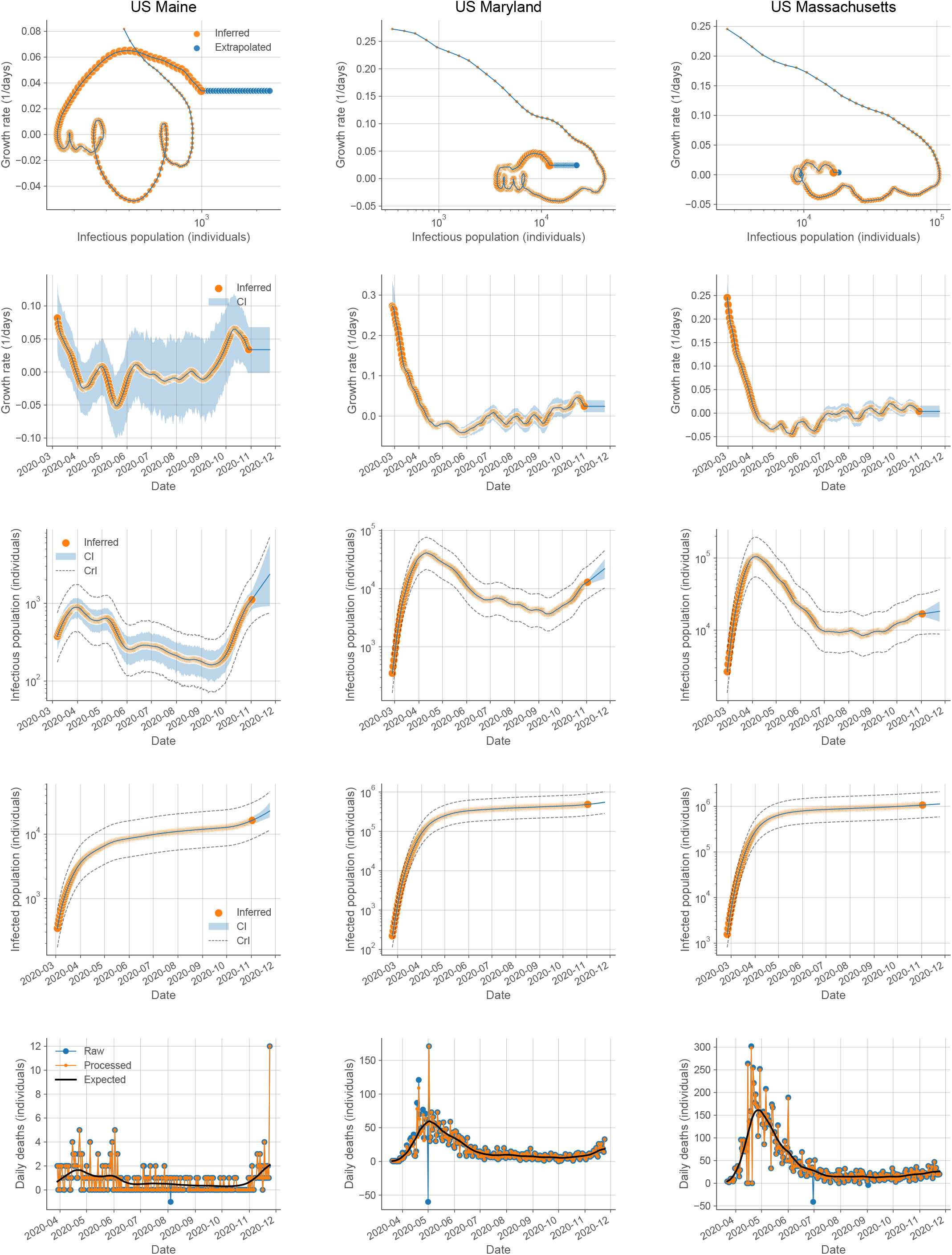
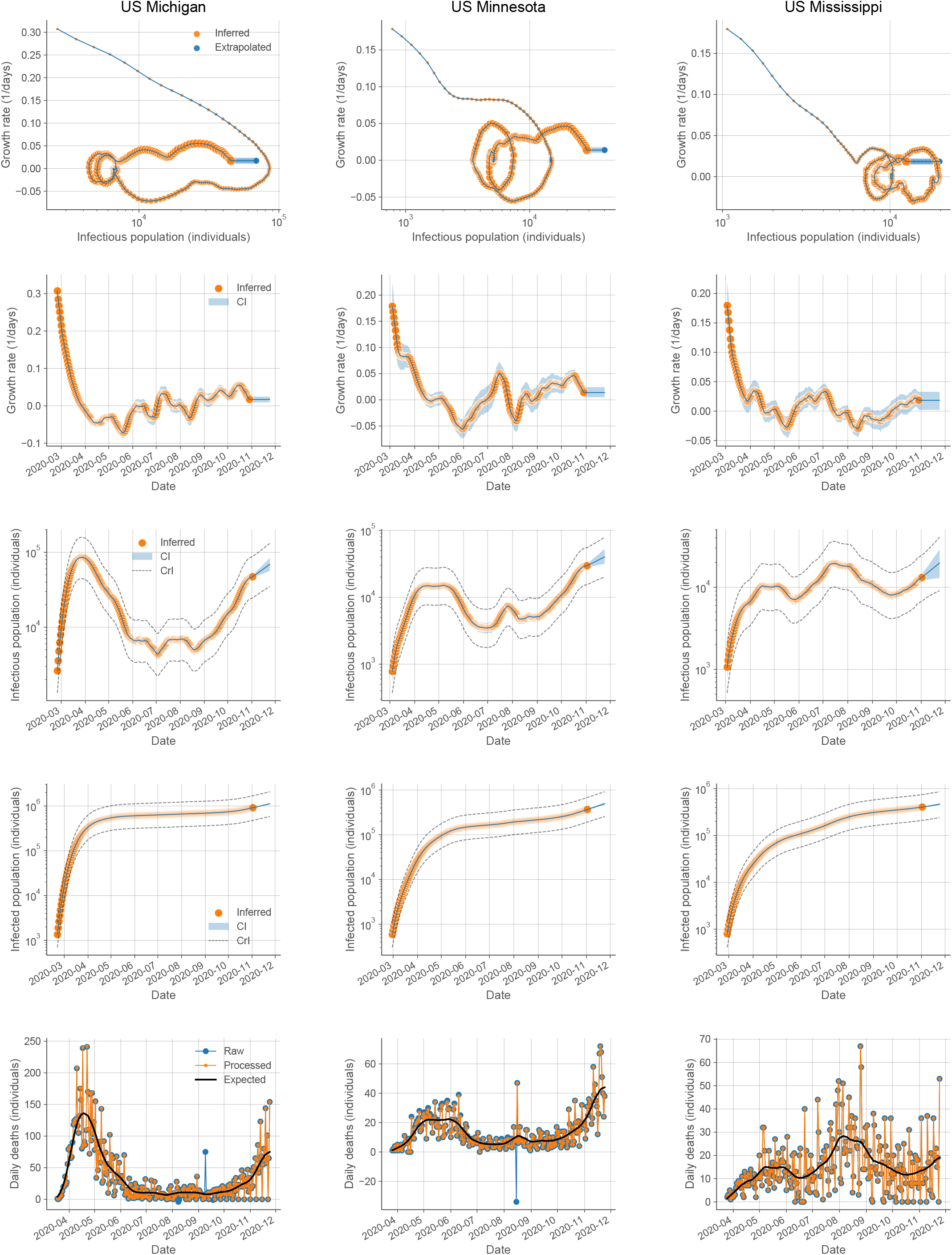
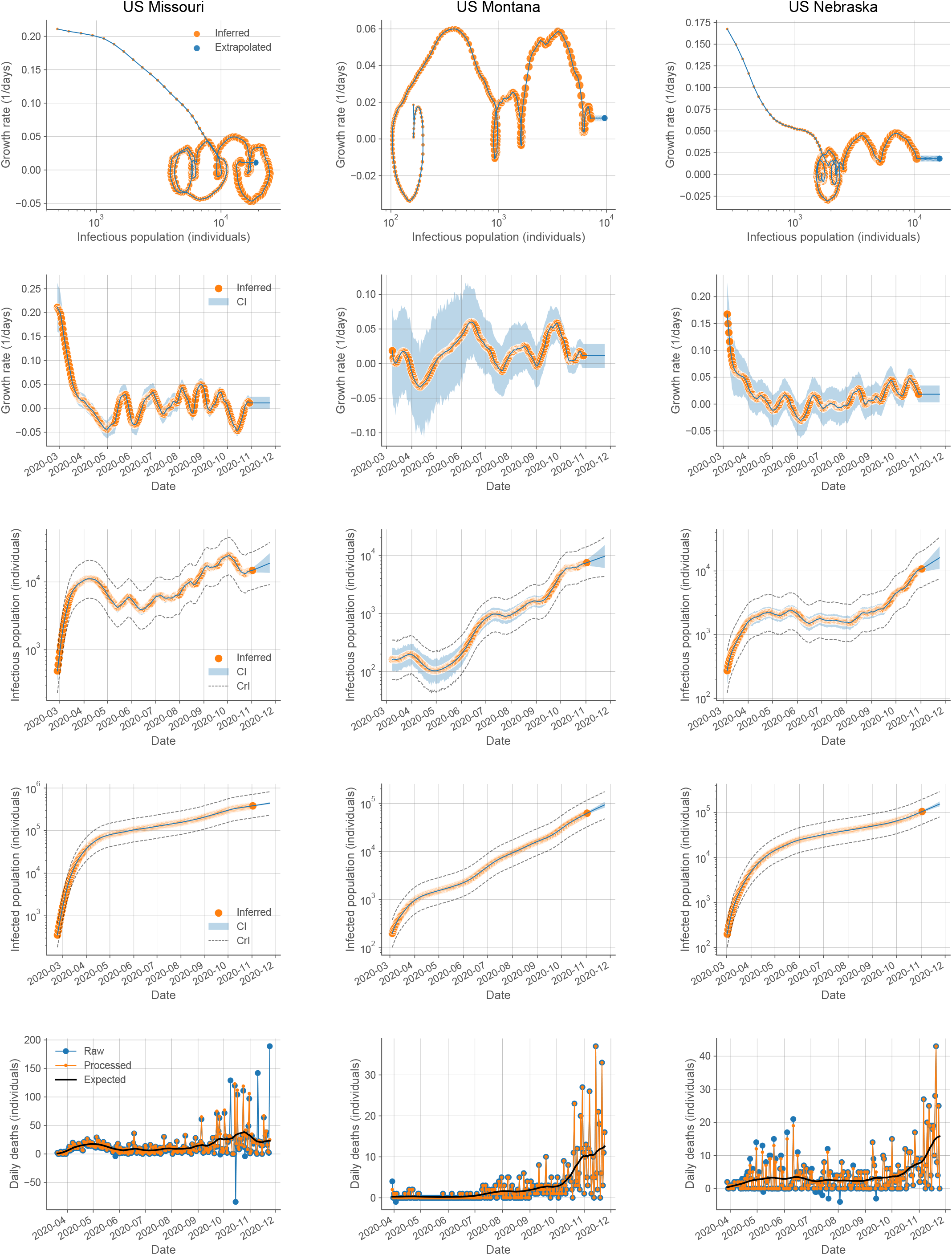
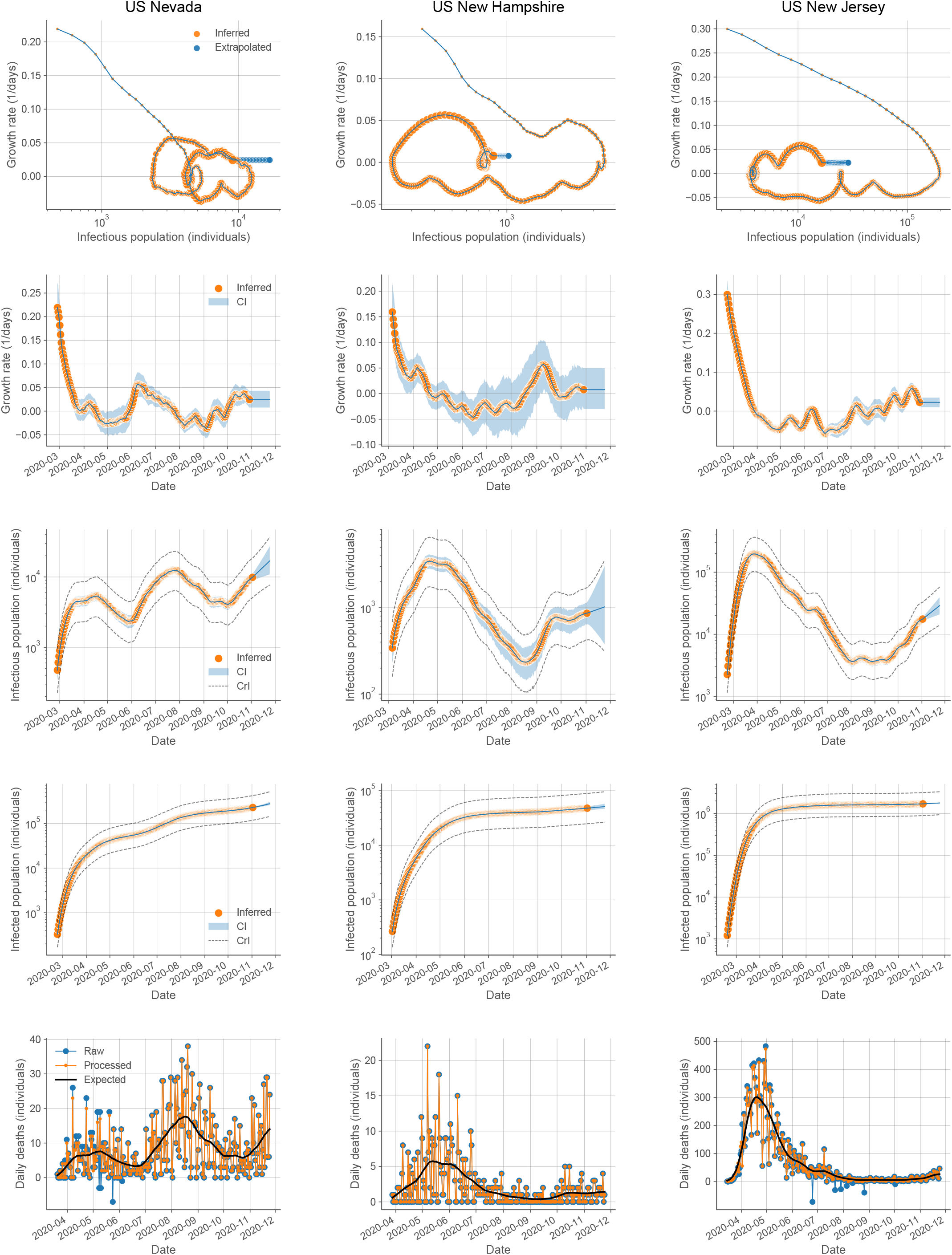
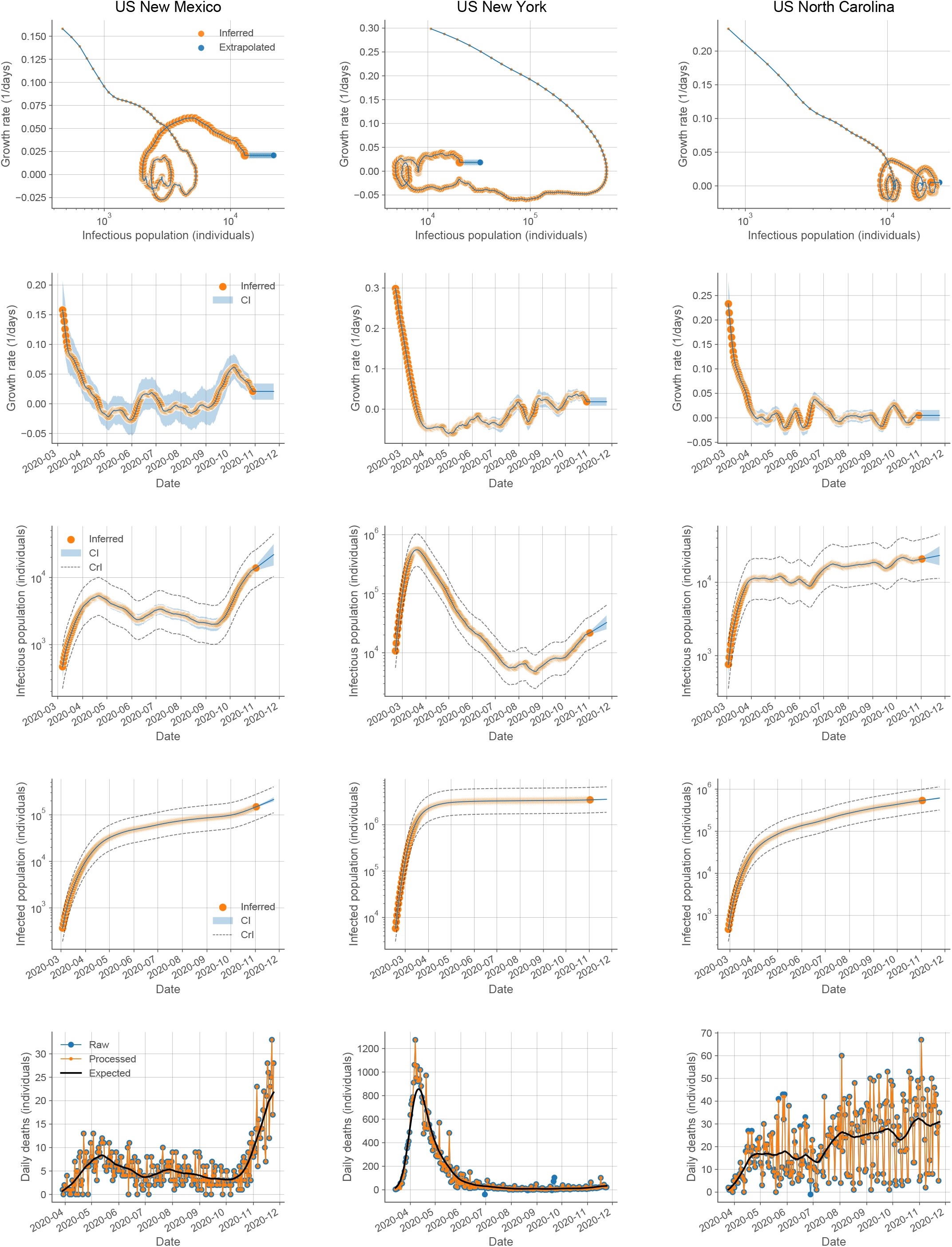
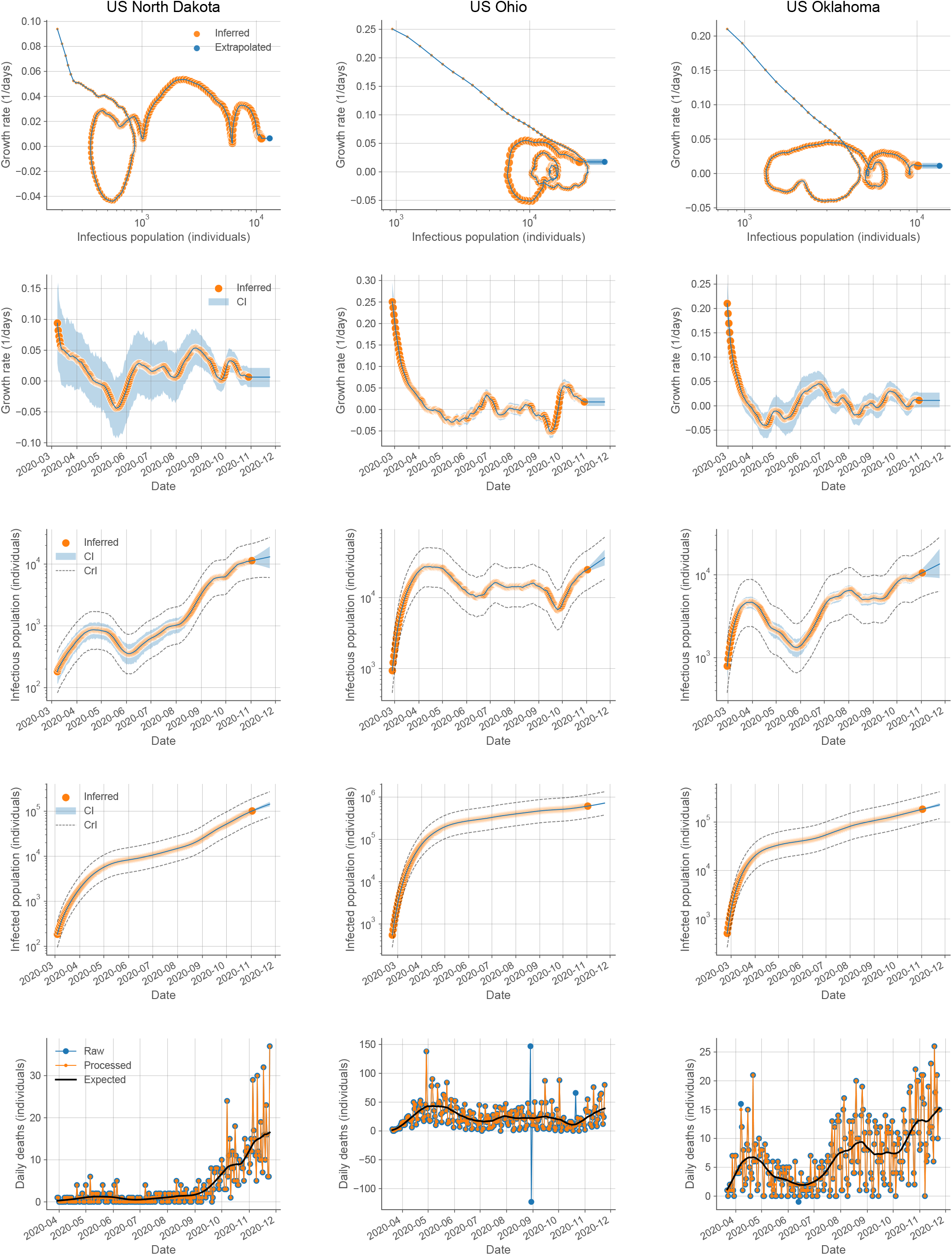
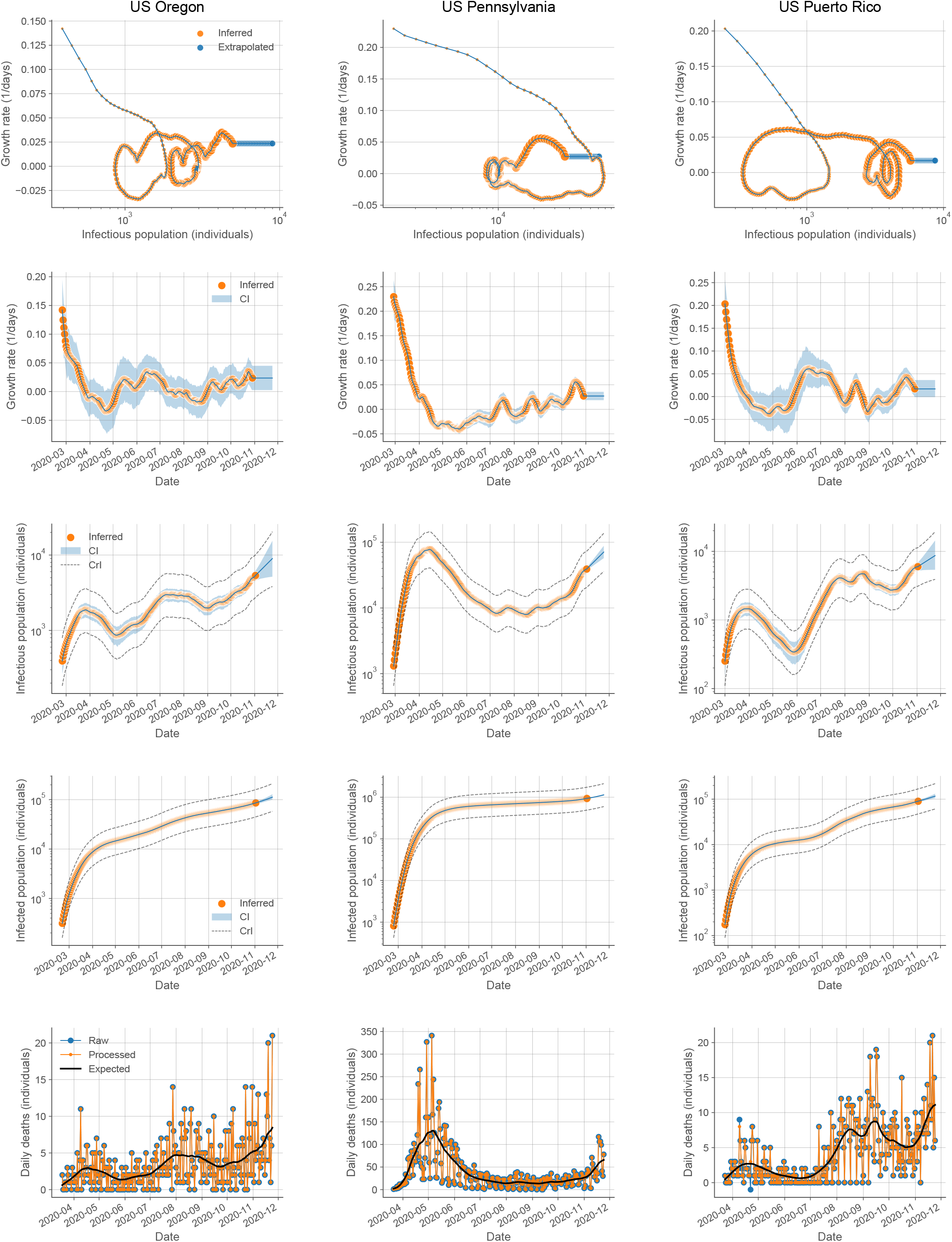
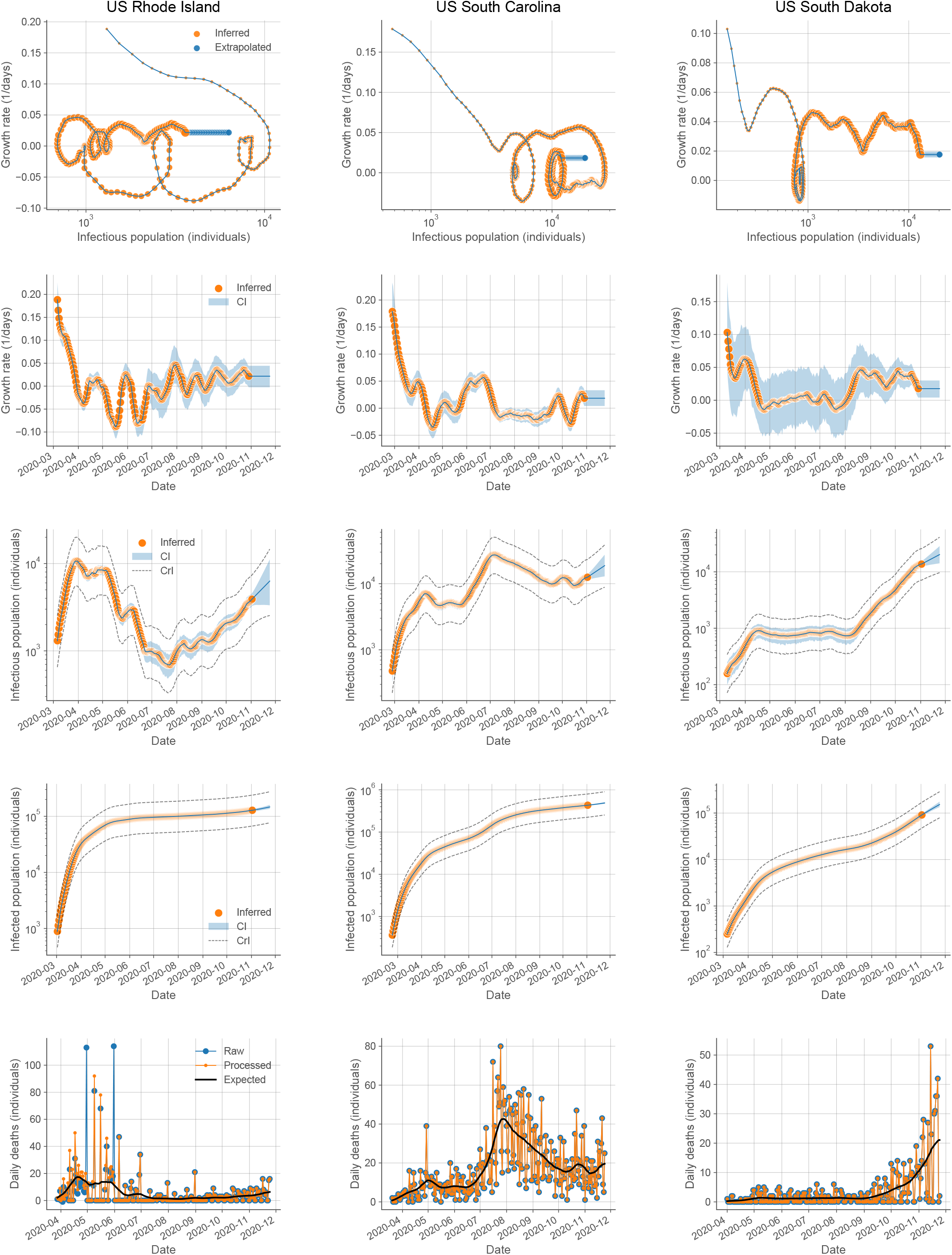
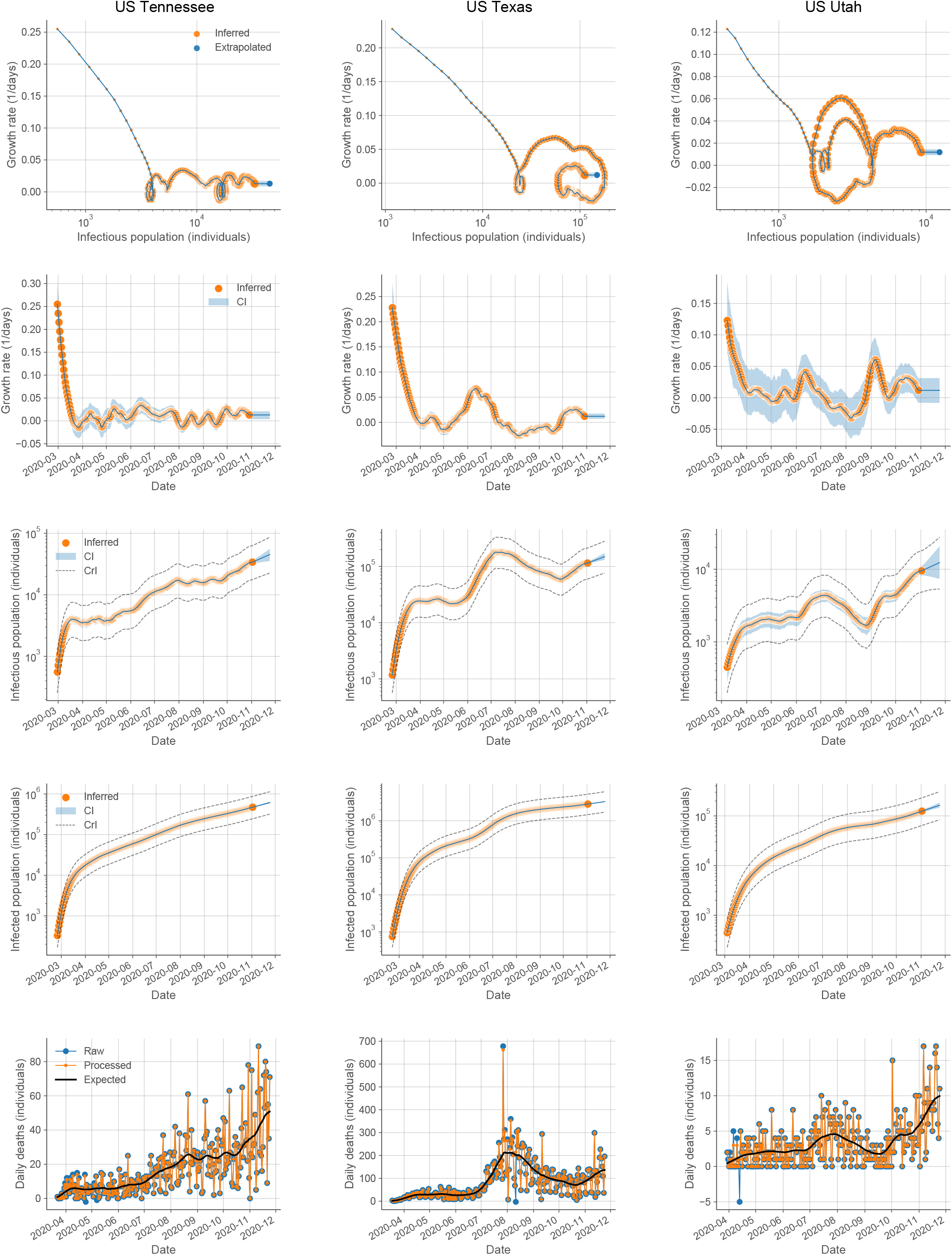
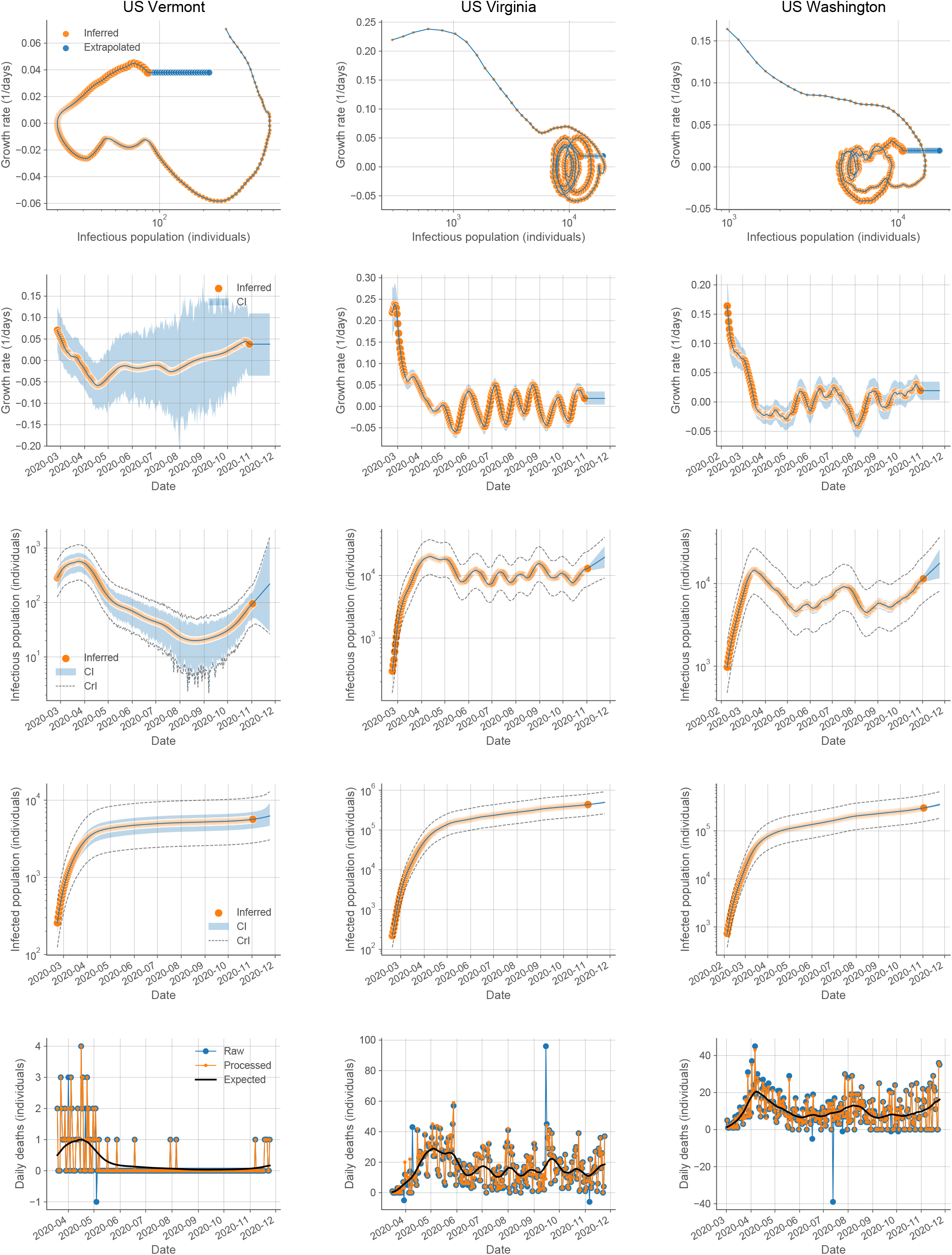
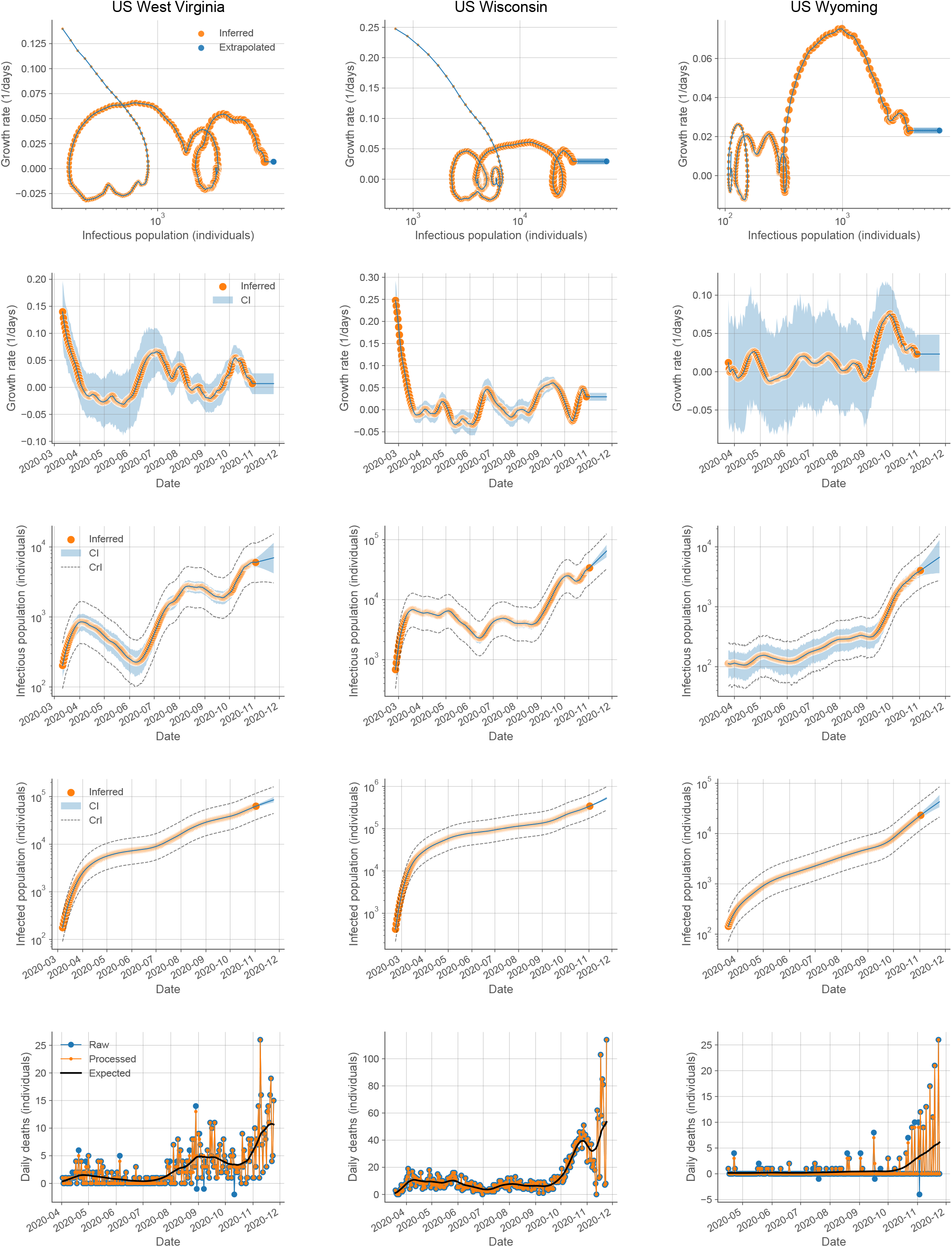
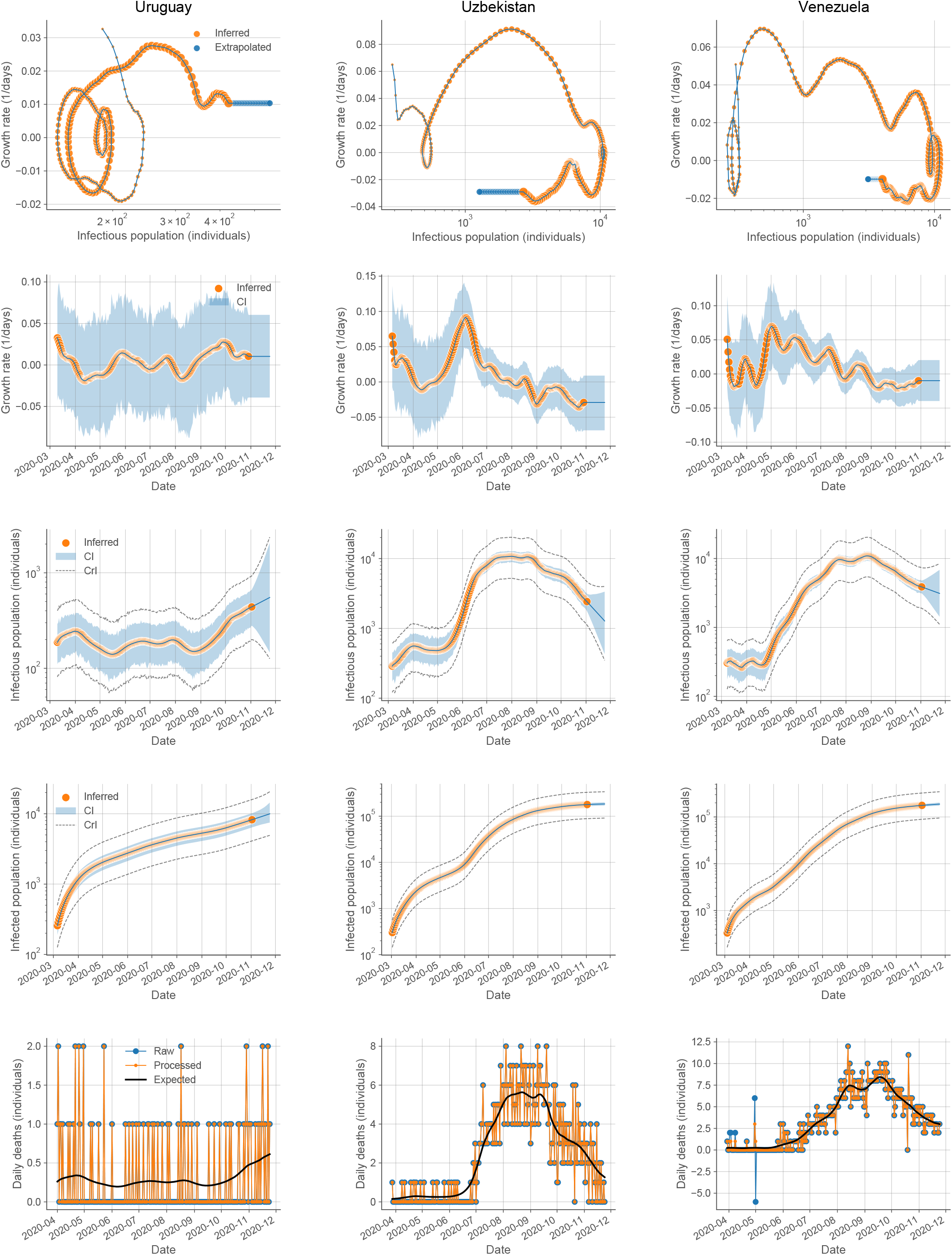
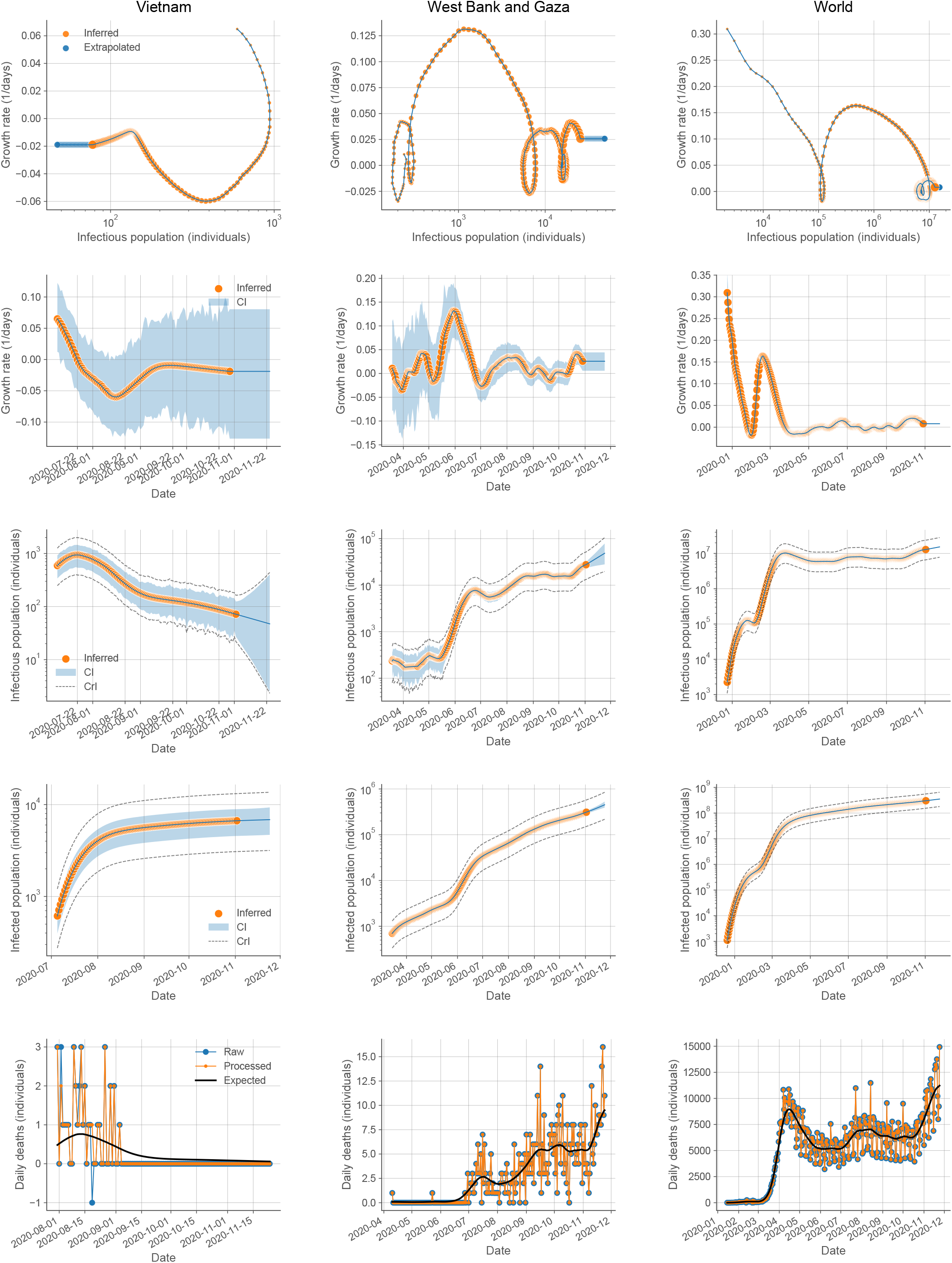
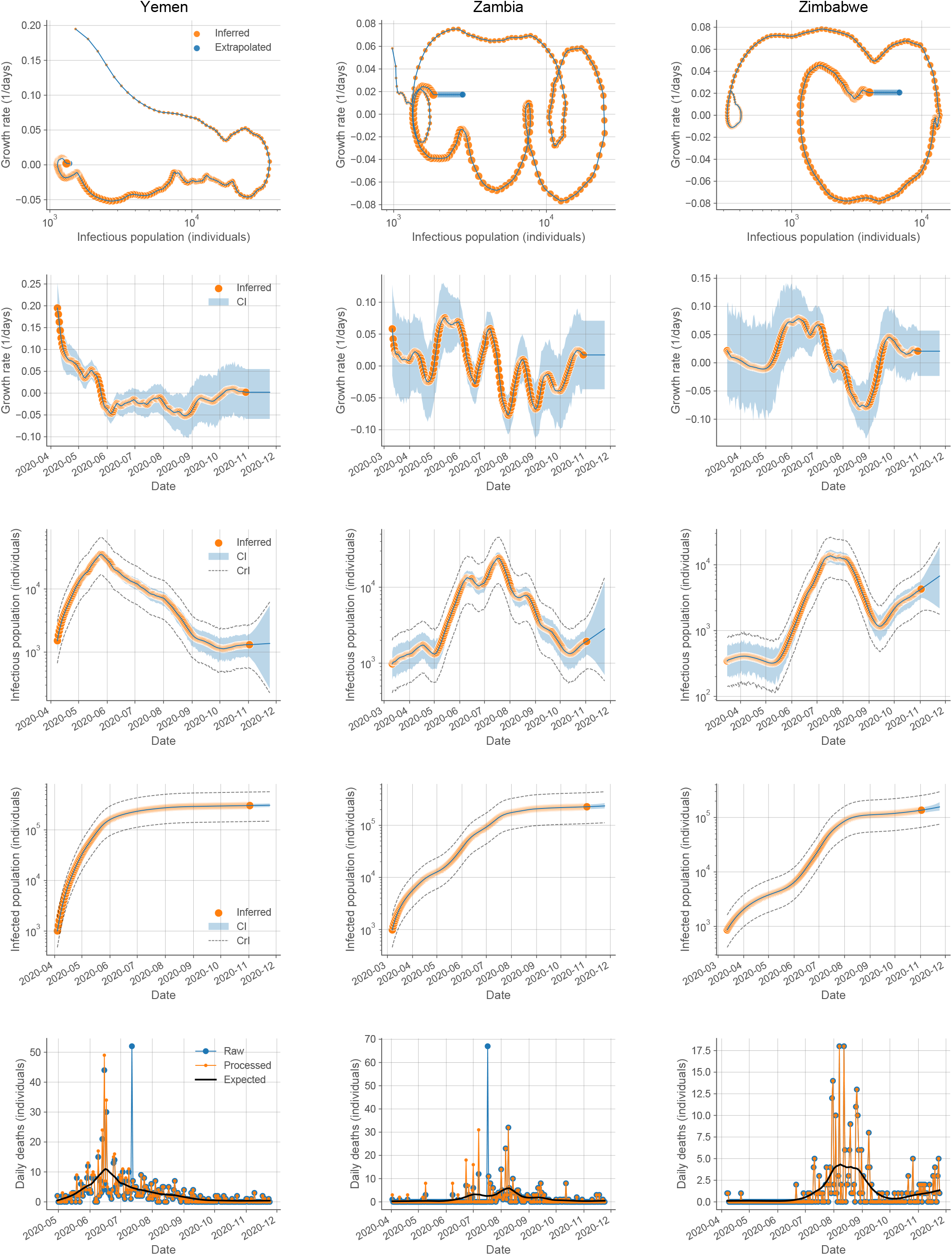
Characterization of the temporal evolution of the COVID-19 outbreak for each of the countries in the World and states and territories in the US. The figure includes 207 locations. The data for each location is shown in a column of 5 panels with its name on the top. The 1st panel from the top shows the trajectory in the growth rate-infectious population space. Each day is indicated by a symbol increasing in size with time. The largest symbol corresponds to the last day of the estimation (November 2, 2020). The blue line at the end indicates the extrapolation to the current time (November 24, 2020) assuming for the growth rate its last estimated value. The 2nd panel from the top shows the temporal evolution of the growth rate (orange circles) and its extrapolation (blue line without circles). The shaded blue region indicates the 95% confidence intervals (CI). The 3rd panel from the top shows the temporal evolution of the infectious population (orange circles) and its extrapolation (blue line without circles). The shaded blue region indicates the 95% CI assuming a certain *IFR*. The dashed lines indicate the overall 95% credibility intervals (CrI) taking into account the uncertainty in the estimates of the *IFR*. The 4th panel from the top shows the temporal evolution of the infected population (orange circles) and its extrapolation (blue line without circles). The shaded blue region indicates the 95% CI assuming a certain *IFR*. The dashed lines indicate the overall 95% CrI taking into account the uncertainty in the estimates of the *IFR*. The 5th panel from the top shows the raw reported daily deaths (blue circles), the processed daily deaths to mitigate reporting artifacts (orange circles), and the expected deaths (black curve). Countries are arranged in alphabetical order. Locations in the US are arranged in alphabetical order after the data for the “United States” as a country. The prefix “US “has been added to the name of the locations in the US. The analyses for the locations “World” and “United States” have been computed with their overall deaths and demographics in contrast to Fig. 3 in the main text, which considered the cumulative contributions of countries and states. Death counts for Spain between April 1 and November 4, 2020 were obtained from “https://www.mscbs.gob.es“[31] because of missing updates in JHU CSSE COVID-19 data [1]. Death counts for China before January 22, not present in JHU CSSE COVID-19 data [1], were obtained from the European Centre for Disease Prevention and Control [40]. The following locations were not included in the analysis because of lack of demographic information in the United Nations or US Census data: Andorra, Diamond Princess, Dominica, Holy See, Kosovo, Liechtenstein, Marshall Islands, Monaco, MS Zaandam, Saint Kitts and Nevis, San Marino, US Diamond Princess, US Grand Princess, US Northern Mariana Islands, and US Virgin Islands. The following locations were not included in the analysis because they did not have at least 30 reported COVID-19 deaths: Antigua and Barbuda, Barbados, Bhutan, Brunei, Burundi, Cambodia, Comoros, Eritrea, Fiji, Grenada, Iceland, Laos, Mauritius, Mongolia, New Zealand, Papua New Guinea, Saint Lucia, Saint Vincent and the Grenadines, Sao Tome and Principe, Seychelles, Singapore, Solomon Islands, Taiwan, Tanzania, Timor-Leste, US American Samoa, Vanuatu, and Western Sahara.

## Notes

### Competing Interest Statement

The authors have declared no competing interest.

### Funding Statement

J.M.G.V. acknowledges support from Ministerio de Ciencia e Innovacion under grant PGC2018-101282-B-I00 (MCI/AEI/FEDER, UE). L.S. acknowledges support from the University of California, Davis.

## REFERENCES

1. JHU CSSE COVID-19 Data. 2020; Available from: https://github.com/CSSEGISandData/COVID-19.

2. Li, Y., et al. The temporal association of introducing and lifting non-pharmaceutical interventions with the time-varying reproduction number (R) of SARS-CoV-2: a modelling study across 131 countries. The Lancet Infectious Diseases, 2020. DOI: 10.1016/s1473-3099(20)30785-4.

3. Ma, J., Estimating epidemic exponential growth rate and basic reproduction number. Infect Dis Model, 2020. 5: p. 129–141.

4. Wu, J.T., et al., Estimating clinical severity of COVID-19 from the transmission dynamics in Wuhan, China. Nat Med, 2020. 26(4): p. 506–510.

5. Verity, R., et al., Estimates of the severity of coronavirus disease 2019: a model-based analysis. Lancet Infect Dis, 2020. 20(6): p. 669–677.

6. Rothe, C., et al., Transmission of 2019-nCoV Infection from an Asymptomatic Contact in Germany. N Engl J Med, 2020. 382(10): p. 970–971.

7. Böhmer, M.M., et al., Investigation of a COVID-19 outbreak in Germany resulting from a single travel-associated primary case: a case series. The Lancet Infectious Diseases, 2020. 20(8): p. 920–928.

8. Li, R., et al., Substantial undocumented infection facilitates the rapid dissemination of novel coronavirus (SARS-CoV-2). Science, 2020. 368(6490): p. 489–493.

9. Flaxman, S., et al., Estimating the effects of non-pharmaceutical interventions on COVID-19 in Europe. Nature, 2020. 584(7820): p. 257–261.

10. Grassly, N.C. and C. Fraser, Mathematical models of infectious disease transmission. Nat Rev Microbiol, 2008. 6(6): p. 477–87.

11. Hethcote, H.W., The mathematics of infectious diseases. SIAM review, 2000. 42(4): p. 599–653.

12. Bi, Q., et al., Epidemiology and transmission of COVID-19 in 391 cases and 1286 of their close contacts in Shenzhen, China: a retrospective cohort study. The Lancet Infectious Diseases, 2020. 20(8): p. 911–919.

13. Backer, J.A., D. Klinkenberg, and J. Wallinga Incubation period of 2019 novel coronavirus (2019-nCoV) infections among travellers from Wuhan, China, 20-28 January 2020. Euro Surveill, 2020. 25, DOI: 10.2807/1560-7917.ES.2020.25.5.2000062.

14. Long, Q.-X., et al., Clinical and immunological assessment of asymptomatic SARS-CoV-2 infections. Nature Medicine, 2020. 26(8): p. 1200–1204.

15. Wolfel, R., et al., Virological assessment of hospitalized patients with COVID-2019. Nature, 2020. 581(7809): p. 465–469.

16. Zhou, F., et al., Clinical course and risk factors for mortality of adult inpatients with COVID-19 in Wuhan, China: a retrospective cohort study. Lancet, 2020. 395(10229): p. 1054–1062.

17. UN World Population Prospects: Total Population - Both Sexes. 2020; Available from: https://population.un.org/wpp/Download/Standard/Population.

18. US Census: ACS Demographic and Housing Estimates 2018. 2019; Available from: https://data.census.gov/cedsci.

19. Anand, S., et al. Prevalence of SARS-CoV-2 antibodies in a large nationwide sample of patients on dialysis in the USA: a cross-sectional study. Lancet, 2020. DOI: 10.1016/S0140-6736(20)32009-2.

20. CDC COVID Data Tracker: Nationwide Commercial Laboratory Seroprevalence Survey. 2020; Available from: https://covid.cdc.gov/covid-data-tracker/#national-lab.

21. Woolf, S.H., et al., Excess Deaths From COVID-19 and Other Causes, March-July 2020. JAMA, 2020. 324(15): p. 1562–1564.

22. Hale, T., et al., Oxford COVID-19 Government Response Tracker, Blavatnik School of Government. 2020.

23. Levin, A.T., et al. Assessing the Age Specificity of Infection Fatality Rates for COVID-19: Systematic Review, Meta-Analysis, and Public Policy Implications. medRxiv, 2020. DOI: 10.1101/2020.07.23.20160895.

24. Pollan, M., et al., Prevalence of SARS-CoV-2 in Spain (ENE-COVID): a nationwide, population-based seroepidemiological study. Lancet, 2020. 396(10250): p. 535–544.

25. Erikstrup, C., et al. Estimation of SARS-CoV-2 infection fatality rate by real-time antibody screening of blood donors. Clin Infect Dis, 2020. DOI: 10.1093/cid/ciaa849.

26. Ali, S.T., et al., Serial interval of SARS-CoV-2 was shortened over time by nonpharmaceutical interventions. Science, 2020. 369(6507): p. 1106–1109.

27. Herzog, S., et al. Seroprevalence of IgG antibodies against SARS coronavirus 2 in Belgium: a serial prospective cross-sectional nationwide study of residual samples. medRxiv, 2020. 2020.06.08.20125179 DOI: 10.1101/2020.06.08.20125179.

28. Walker, A.S., et al. Viral load in community SARS-CoV-2 cases varies widely and temporally. medRxiv, 2020. DOI: 10.1101/2020.10.25.20219048.

29. Vilar, J.M.G. and J.M. Rubi, Determinants of population responses to environmental fluctuations. Sci Rep, 2018. 8(1): p. 887.

30. Saad-Roy, C.M., et al., Immune life history, vaccination, and the dynamics of SARS-CoV-2 over the next 5 years. Science, 2020. 370(6518): p. 811–818.

31. Ministerio de Sanidad. Fallecidos COVID-19. 2020; Available from: https://www.mscbs.gob.es/profesionales/saludPublica/ccayes/alertasActual/nCov-China/documentos/Fallecidos_COVID19.xlsx.

32. Hastie, T., R. Tibshirani, and J. Friedman, The Elements of Statistical Learning: Data Mining, Inference, and Prediction. 2013: Springer New York.

33. Salje, H., et al., Estimating the burden of SARS-CoV-2 in France. Science, 2020. 369(6500): p. 208–211.

34. Hallal, P., et al. Remarkable variability in SARS-CoV-2 antibodies across Brazilian regions: nationwide serological household survey in 27 states. medRxiv, 2020. 2020.05.30.20117531 DOI: 10.1101/2020.05.30.20117531.

35. Bogogiannidou, Z., et al. Repeated leftover serosurvey of SARS-CoV-2 IgG antibodies, Greece, March and April 2020. Eurosurveillance, 2020. 25, DOI: 10.2807/1560-7917.Es.2020.25.31.2001369.

36. Gallian, P., et al., Lower prevalence of antibodies neutralizing SARS-CoV-2 in group O French blood donors. Antiviral Research, 2020. 181: p. 104880.

37. Menachemi, N., et al., Population Point Prevalence of SARS-CoV-2 Infection Based on a Statewide Random Sample — Indiana, April 25–29, 2020. MMWR. Morbidity and Mortality Weekly Report, 2020. 69(29): p. 960–964.

38. Tunheim, G., et al., Seroprevalence of SARS-CoV-2 in the Norwegian population measured in residual sera collected in April/May 2020 and August 2019. 2020, Norwegian Institute of Public Health, ISBN (digital): 978-82-8406-109-2.

39. Sutton, M., P. Cieslak, and M. Linder, Notes from the Field: Seroprevalence Estimates of SARS-CoV-2 Infection in Convenience Sample - Oregon, May 11-June 15, 2020. MMWR Morb Mortal Wkly Rep, 2020. 69(32): p. 1100–1101.

40. European Centre for Disease Prevention and Control. Daily update of new reported cases of COVID-19 by country worldwide. 2020; Available from: https://www.ecdc.europa.eu/sites/default/files/documents/COVID-19-geographic-disbtribution-worldwide.xlsx.

